# A Data Science Pipeline Applied to Australia’s 2022 COVID-19 Omicron Waves

**DOI:** 10.1101/2023.12.04.23299260

**Authors:** James M Trauer, Angus E Hughes, David S Shipman, Michael T Meehan, Alec S Henderson, Emma S McBryde, Romain Ragonnet

## Abstract

The field of software engineering is advancing at astonishing speed, with packages now available to support many stages of data science pipelines. These packages can support infectious disease modelling to be more robust, efficient and transparent, which has been particularly important during the COVID-19 pandemic. We developed a package for the construction of infectious disease models, integrated this with several open-source libraries and applied this pipeline to multiple data sources that provided insights into Australia’s 2022 COVID-19 epidemic. We aimed to identify the key processes relevant to COVID-19 transmission dynamics and thereby develop a model that could quantify relevant epidemiological parameters. Extending the base model to include mobility effects slightly improved model fit to data, but including the effect of 2022 vaccination programs on transmission did not. Our simulations suggested that one in every two to six COVID-19 episodes were detected, subsequently emerging Omicron subvariants escaped 30 to 60% of recently acquired natural immunity and that natural immunity lasted only one to eight months. We documented our analyses algorithmically and present our methods in conjunction with interactive online notebooks.

## Introduction

Throughout the pandemic, epidemiological modelling has been used to influence some of the most significant and intrusive public health policy decisions in history, providing analyses to justify a range of programs that extended from lockdowns to vaccination.^1,2^ However, this policy impact brings with it a responsibility for modellers to ensure results are accurate, transparent and effectively communicated not only to policy makers, but also the public who are impacted by such decisions.^2^

While libraries are available to support many components of the data science pipeline, the application of software engineering principles to epidemiological modelling remains limited.^3^ The rapid growth of data science as a field, and the corresponding investment in platform development provides constant opportunities to expand the range of packages that can be integrated with such models, provided the model code itself is developed with this in mind. In particular, a software package whose single responsibility is model construction can separate this concern from the multiple other stages in the formulation of a modelling-based analysis.

Australia’s 2022 epidemic provides an important case study for understanding the epidemiological characteristics of COVID-19, because of the distinct epidemic waves, negligible prevalence of natural immunity from past infection,^4-6^ stable vaccination coverage and multiple high-quality data streams. Australia pursued an elimination approach through the first two years of the pandemic, achieving one of the lowest COVID-19-related mortality rates in the world.^7^ Shortly after achieving very high coverage of wild-type vaccination, most of the country emerged abruptly from this elimination phase in 2022, relaxing most restrictions on population mobility as the Omicron variant rapidly replaced the preceding Delta SARS-CoV-2 variant of concern.^8^ Through 2022, national data are available that include a daily time-series for cases and deaths, serial survey-derived testing behaviour, population mobility, vaccination coverage and seroprevalence. Of particular value, the serial serosurveys demonstrate a rapid rise in nucleocapsid antibodies to more than 65% seroprevalence by late August 2022.^9^ The combination of these data sources provides the opportunity to improve our understanding of COVID-19 epidemiology in the Omicron era, such as quantification of the case detection rate and characteristics of population immunity.

We developed a suite of software tools to support evidence-based policies, which was applied to several countries of the Asia-Pacific Region in collaboration with the World Health Organization and other regional public health agencies.^10-13^ Our platform is based around a library for the construction of compartmental models of infectious disease transmission and is now integrated with publicly available libraries for numerical computing, optimisation, Bayesian inference, data visualisation and scientific documentation.^14-17^ The combined platform constitutes an end-to-end pipeline for infectious disease modelling, which we used to derive insights into COVID-19 epidemiology through its application to Australia’s three-wave 2022 epidemic.

## Results

We released a suite of open-source packages to support infectious disease modelling and used these packages to represent the key epidemiological processes relevant to Australia’s 2022 COVID-19 epidemic (Figure 1). At the heart of our pipeline, we developed the *summer* Python package to support easy and reliable construction of infectious disease models through an epidemiologically intuitive application programming interface. *summer*’s backend is integrated with Google’s *jax* library for high-performance numerical computing. We validated *summer* against a popular textbook of infectious diseases modelling,^18^ demonstrating that it could recover the behaviours of a wide range of infectious diseases models through a series of [jupyter notebooks](https://github.com/monash-emu/summer-vynnycky-white-validation). Next, through a series of interactive Google Colab-hosted [notebooks](https://github.com/monash-emu/summer-textbook), we released a textbook of infectious disease modelling that systematically demonstrates core infectious disease modelling principles using *summer*. Last, we released *estival*, a wrapper for the calibration and optimisation of *summer*-based models, which supports the integration of these models with current state-of-the-art calibration, optimisation and interpretation platforms, including the *PyMC* package for Bayesian inference^14^ and Facebook’s *nevergrad* library for gradient-free optimisation.^15^ *ArviZ* was used for Bayesian diagnostics,^16^ with interactive visuals produced using *plotly*.^17^

**Figure 1.**
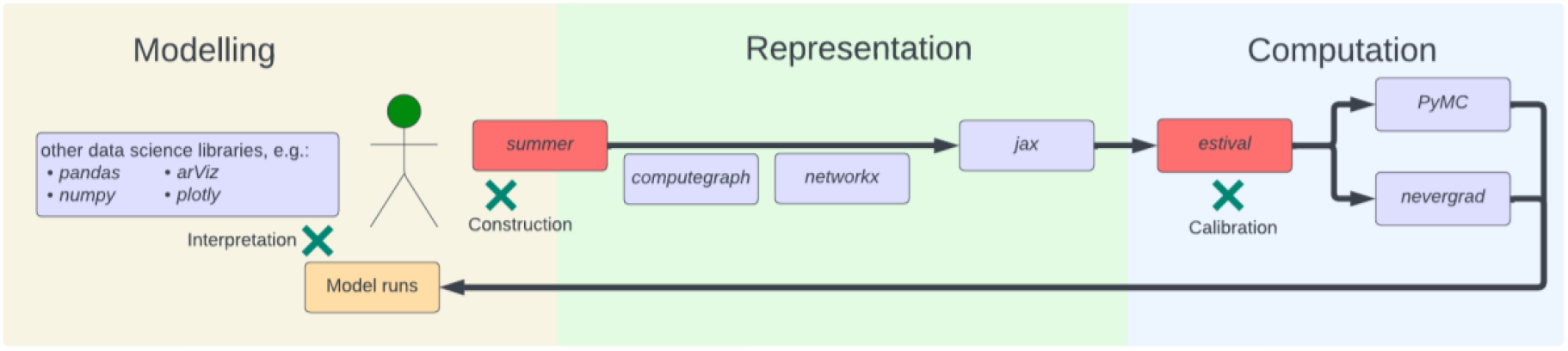
Computational structure of our modelling pipeline. Red-coloured boxes represent packages developed by our team, mauve-coloured boxes represent publicly available packages, green crosses represent points for user interaction.

The analysis is provided as an installable Python package that incorporates interactive Google Colab Jupyter notebooks for the inspection of model features and interrogation of outputs. Through this approach, we fit a complex model of COVID-19 dynamics (1984 compartments under the base case, 2976 compartments under the vaccination extension) to three key epidemiological indicators (cases, deaths and nucleocapsid antibody seroprevalence a marker of ever being infected).

### Candidate model comparison

We considered four candidate models with and without extended structure for mobility and vaccination for their ability to capture the broad epidemic profile of Australia’s 2022 Omicron waves (i.e. mobility extension, vaccination extension, both extensions, and neither). All four models were able to capture the broad epidemic profile we targeted, with each achieving a good fit to the time-series of deaths. The model configurations with additional structure for scaling contact rates with mobility data achieved a somewhat better fit to the seroprevalence targets than the two configurations without this extension (median seroprevalence likelihood contribution 0.6 versus 0.9, Figure 2). Inclusion of additional model structure for time-varying vaccination-related immunity to infection resulted in a slightly poorer fit to the time-series for cases (median cases likelihood contribution –12.7 versus –12.2 to –12.4 for the other analyses). We therefore selected the mobility extension model as the primary model analysis for consideration in the following sections. Additional approaches to model construction, calibration and interpretation of results can easily be explored via the interactive notebooks available on the project homepage.

**Figure 2.**
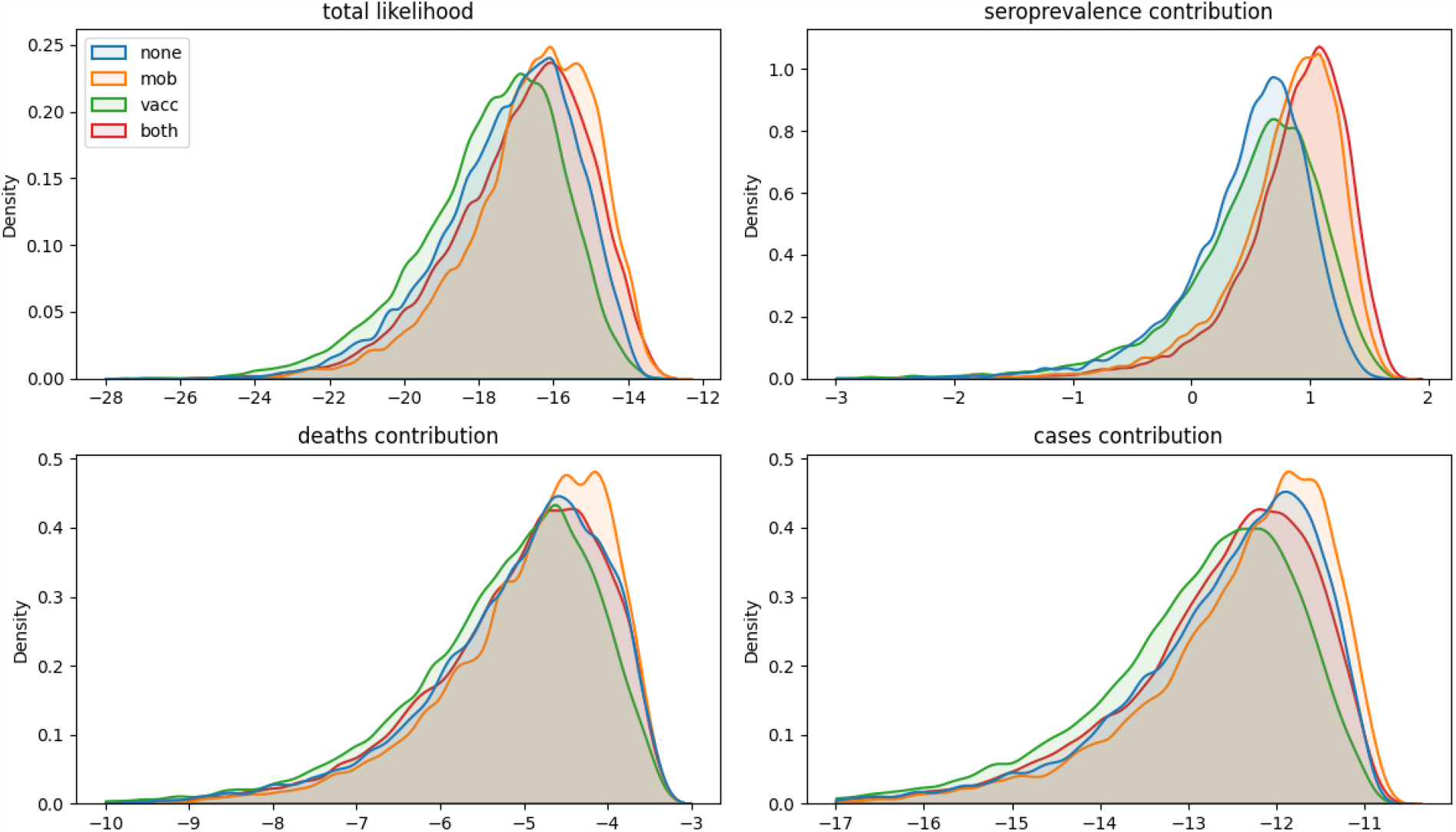
Likelihood comparison. Comparison of the kernel density distribution of the final likelihood from calibration algorithm, with the contributions to the final likelihood of the three targets from which it was constructed.

### Calibration results

Leveraging Google’s *jax* package and calibration algorithms from *PyMC*, epidemiological models of 1984 to 2976 compartments (depending on application of the vaccination extension) completed 60,000 iterations per chain within 4-8 hours on 8-core 3rd Generation Intel Xeon machines clocked at 2.9 to 3.5 GHz. For the primary (mobility extension) analysis, each core completed 3.62 model iterations per second. Metrics of the calibration algorithm for the primary analysis are presented in the Supplemental Material. The algorithm achieved highly satisfying chain convergence, with the Rhat statistic for all parameters below 1.05 and all effective sample sizes above 150 (Supplemental Figure 4).

Figure 3 shows model fit for each of the target epidemiological indicators. Calibration fit was better for deaths than for case notifications, which is reflected in the markedly lower likelihood contributions for the cases calibration target (median –12.2) than for the deaths targets (median – 4.7) (Figure 2). This difference was particularly noticeable during the first (BA.1) wave of 2022, at a time when notifications may have been a more variable epidemic indicator as Australia struggled to scale testing capacity up to match demand (Figure 3).^19^ Under the constraint that BA.1 and BA.5 had the same modelled severity,^20^ accepted model runs often under-estimated the peak number of deaths for the third (BA.5) wave of 2022. Our results typically showed a higher seroprevalence than estimated from the serosurvey target values for its first round, but lower for the third. The relative contribution of each variant and each infection process (i.e. de novo infection, early reinfection due to immune escape and late reinfection due to waned immunity) is presented in Figure 4.

**Figure 3.**
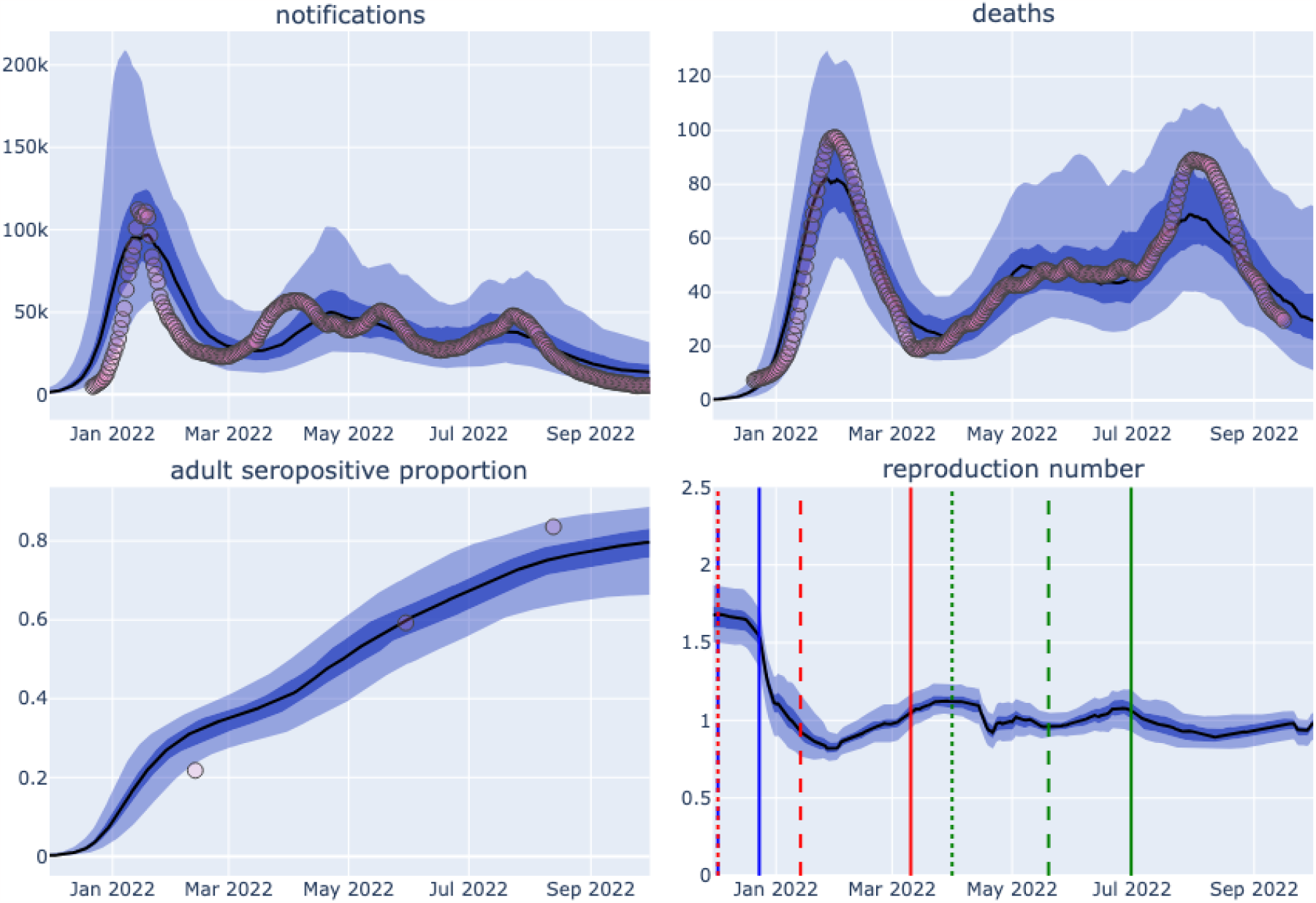
Primary analysis output credible intervals. Model median estimate (black line), 2.5 to 97.5 centile credible interval (light blue shading), and 25 to 75 centile credible interval (dark blue shading), with comparison against epidemiological targets (red circles). Panel for each epidemiological output as indicated. Sampled runs from same calibration also presented as interactive online figures for [cases](https://monash-emu.github.io/outputs/aust-covid/notifications_ma_3dspaghetti.html), [deaths](https://monash-emu.github.io/outputs/aust-covid/deaths_ma_3dspaghetti.html), [seroprevalence](https://monash-emu.github.io/outputs/aust-covid/adult_seropos_prop_3dspaghetti.html) and [reproduction number](https://monash-emu.github.io/outputs/aust-covid/reproduction_number_3dspaghetti.html). Key dates for each variant are shown as vertical bars on lower right panel: blue, BA.1; red, BA.2; green, BA.5; dotted, first detection; dashed, >1% of isolates; solid, >50% of isolates. Proportion of isolates and dates based on reported Pango lineage variant designated proportions for Australia on Cov-Spectrum.^21^

**Figure 4.**
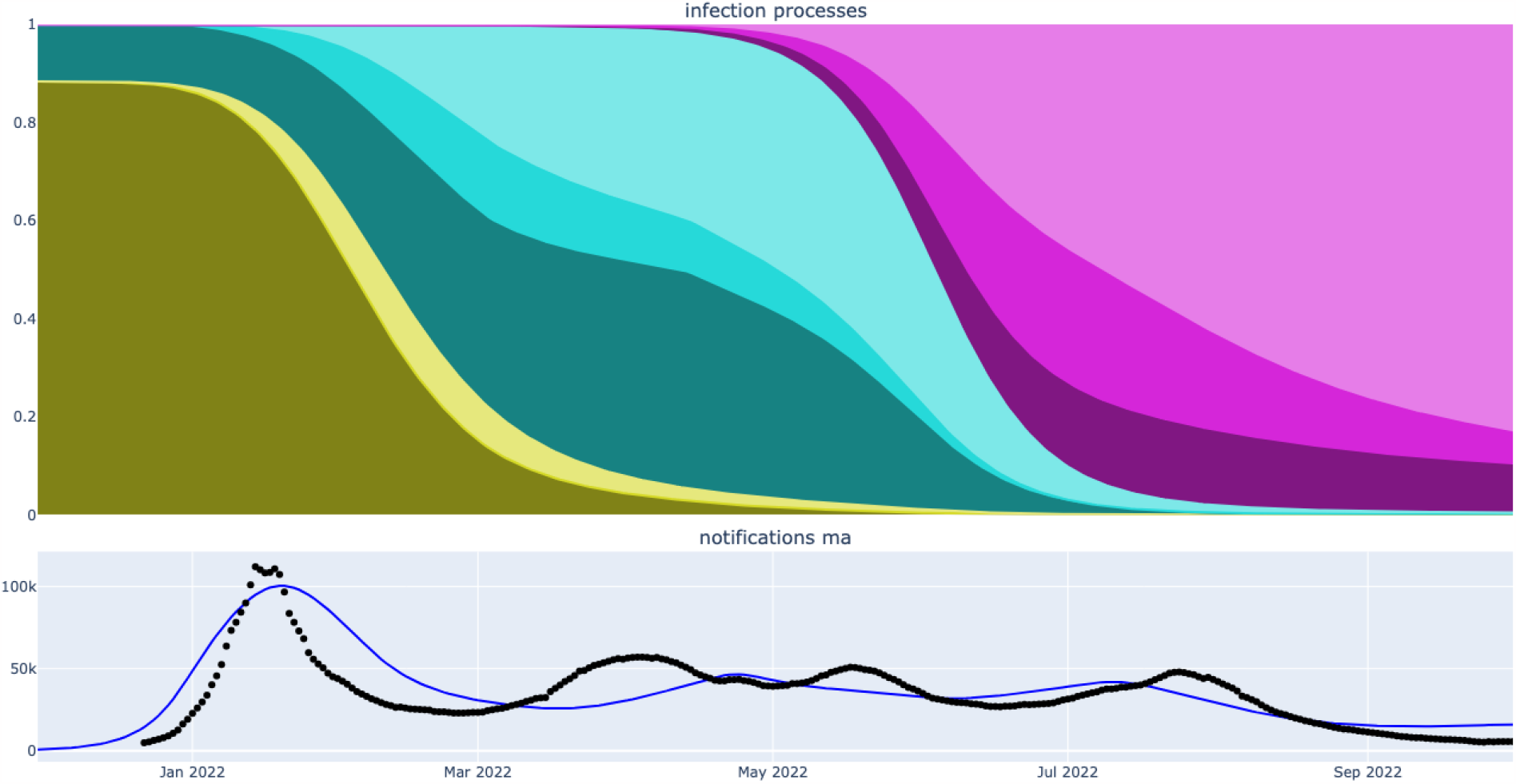
Contribution of various infection processes through the course of the simulated epidemic under the maximum posterior parameter set from the primary (mobility extension) analysis. Colour shows infection with BA.1 (greens), BA.2 (blues) and BA.5 (purples). Shading depth shows infection process, with initial infection (dark), early reinfection (intermediate darkness), late reinfection (light). (Note early reinfection with BA.1 does not occur to a significant extent.)

### Parameter inference

The short prior estimates for the durations for the latent and infectious periods^22,23^ were not substantially influenced by the process of fitting to target data, while the period of partial immunity following infection was estimated to be markedly shorter than our prior belief (Figure 5). Our uninformative prior estimate for the proportion of cases detected was also substantially influenced by the fitting process, suggesting that the highest case detection early in the BA.1 epidemic wave was around 17 to 50%. The infection fatality rate had to be inflated around two- to three-fold from the baseline estimates taken from a population-wide epidemiological study in Denmark.^24^ The extent of immune escape of both BA.2 and BA.5 against previous infection with other sub-variant strains was moderately greater than anticipated in our prior distributions,^25-30^ and was centred around a value of 50% for both subvariants (i.e. past infection only protected half as much against early reinfection with novel subvariants as compared to against early reinfection with the previously infecting subvariant). The relative reduction in severity of BA.2 compared to BA.1 and BA.5 was consistent with past evidence,^20,31-33^ while little additional information was obtained for the time to WA fully mixing with the rest of the country or the parameters pertaining to the convolution processes for notifications and deaths. The seeding time parameters resulted in epidemic profiles that were consistent with reports of national genomic data (see Supplemental Figure 25).^34^ On examination of the bivariate distributions of combinations of two parameters, the contact rate parameter showed expected inverse interactions with the extent of population immunity and the infectious duration parameters (Figure 6). The association of shorter duration of post-infection immunity and a lower case detection proportion can be attributed to both these processes being associated with larger modelled epidemics. The expected association between short duration of post-infection immunity and the extent of BA.2 and BA.5 escape was observed, but was modest.

**Figure 5.**
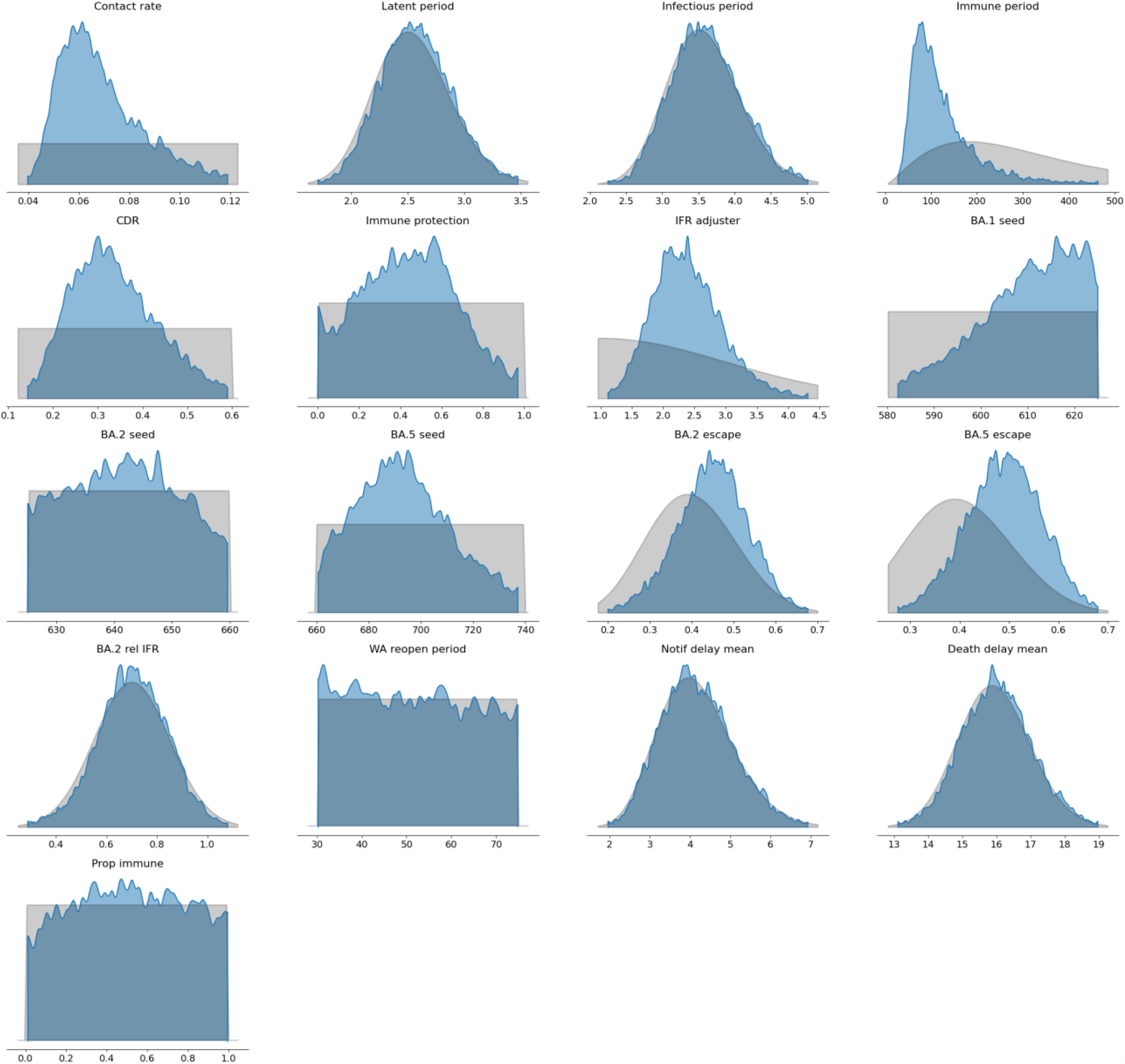
Posterior densities and prior distributions. Inferred parameter posterior densities (blue areas) compared against corresponding calibration algorithm prior distributions (grey areas).

**Figure 6.**
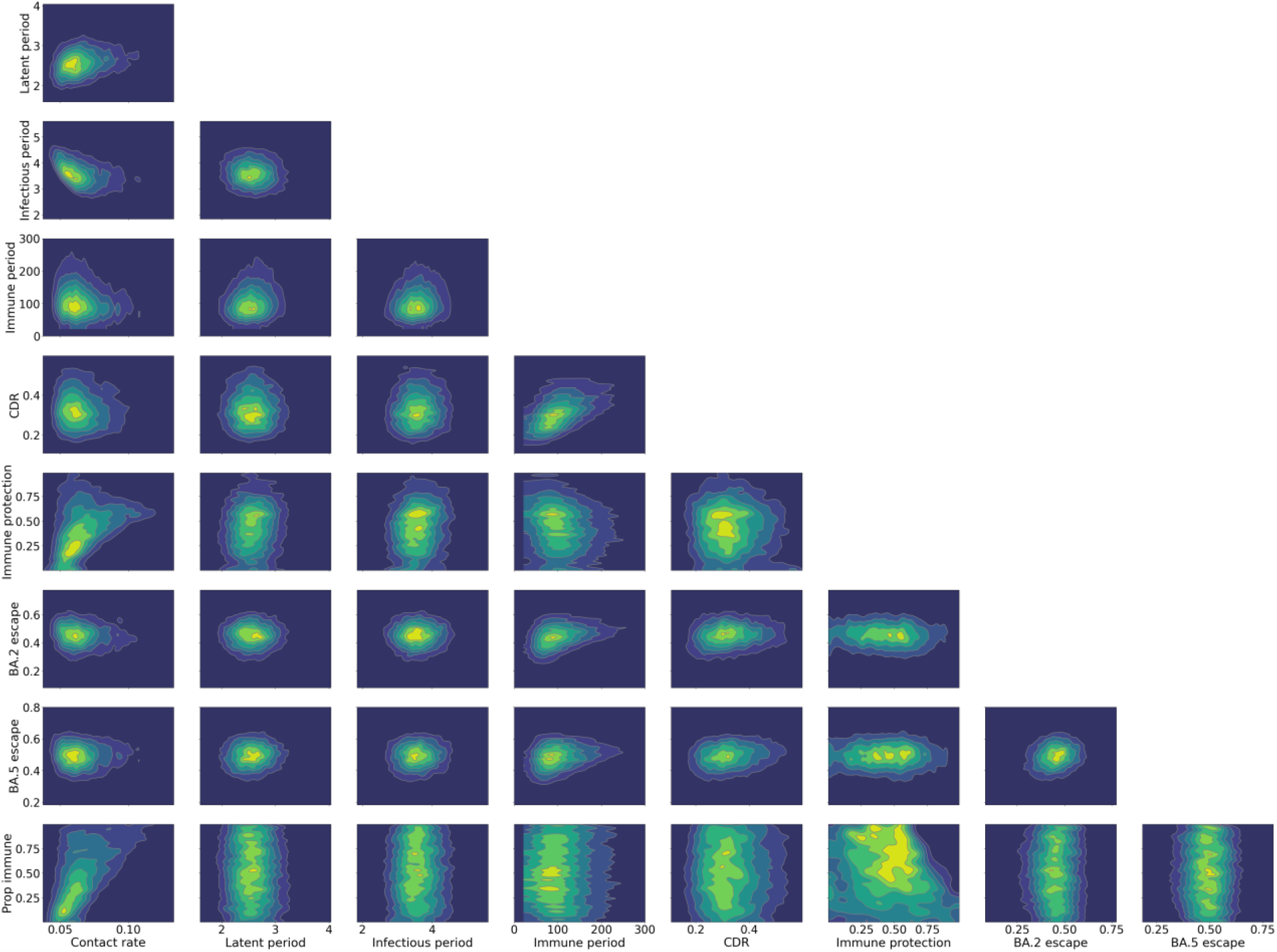
Bivariate distributions of selected parameter combinations for accepted parameter sets from selected (mobility extension) model calibration. Three-way interactive parameter combination plots are available at our interactive outputs page.

## Discussion

Our analysis demonstrates the feasibility of integrating epidemiological modelling with advanced tools in data science, including open-source tools produced by user communities and Big Tech. Our results supported a major role for Omicron subvariants in driving the three major epidemic waves observed in Australia over the course of 2022. With each subvariant epidemic following closely on from the preceding wave, most accepted model configurations required a combination of both high levels of immune escape, as well as a short period of natural immunity. In addition to improving efficiency and expanding the functional possibilities from our platform, our methodological approach reduces the volume of code that the modeller must produce, thereby enhancing transparency and minimising opportunities for errors.

Of our candidate models, all were able to find epidemiologically plausible parameter space and were generally associated with similar results in relation to parameter inference. The coherence of our epidemiological model with multiple data sources simultaneously permitted our analysis to capture a range of model trajectories and associated parameter sets that could accurately represent Australia’s 2022 COVID-19 epidemics. We achieved a closer fit to the time-series of deaths than we achieved for cases, which may be attributable to the higher quality or greater consistency of the mortality data during a period when testing recommendations and test availability changed markedly. By contrast to our expectations in late 2021 when the Omicron variant first emerged in Southern Africa,^35^ the first (BA.1) epidemic wave of 2022 was likely not Australia’s largest, with the subsequent waves (largely attributable to BA.2 and BA.5) also associated with substantial attack rates. The vaccination extension did not improve calibration metrics, which may be attributable to the complex evolving profile of vaccination-induced population immunity, or the limited indirect protection conferred by vaccination.^36^ By contrast, population mobility appeared to improve model fit slightly, possibly by allowing for a smaller initial BA.1 wave through capturing the “shadow lockdown” in summer 2021/2022.^37,38^

Parameter posterior estimates suggested a greater level of immune escape and a shorter duration of immunity than our prior beliefs from the literature,^39-41^ which can be attributed to the rapid succession of each subvariant’s wave as the preceding wave had only recently begun to decline. Our inflation factor for the infection fatality rate suggested considerably greater severity than observed in a population with a longer history of COVID-19 epidemics,^24^ which may relate to lower natural immunity to COVID-19 along with a larger vulnerable population after two years of lower circulation of influenza and other viruses.^42^

The availability of seroprevalence estimates for nucleocapsid antibodies that increased markedly from close to zero to nearly 80% within the simulation window we considered markedly increases our confidence that our analysis captures the overall epidemic size.^9^ Although we did not consider antibody waning, the greatest delay from infection to antibody measurement relevant to our analysis would have been infection early in the BA.1 wave (January 2022) followed by antibody measurement in August 2022 (for persons not infected in the intervening waves), whereas these antibodies are known to be well maintained over ten months.^43^ Moreover, given the apparently large size of Australia’s first BA.1 wave, we found it surprising that adult seroprevalence only reached 20.7% after the first wave, whereas the subsequent estimates implied around 50% of persons not previously infected were exposed within the subsequent windows between survey rounds (February to June and June to August). There are several possible causes of bias in such estimates, including selection bias of blood donors by comparison to the general population,^44^ which has not been quantified in the Australian context to our knowledge. Rather than attempt to adjust for such biases, we acknowledged the bidirectional uncertainty in these data through the likelihood calculation for the seroprevalence calibration target.

More generally, while adjusting our approach to allow for a smaller first wave would have improved fit to the first seroprevalence estimate (e.g. through a higher initial case detection ratio), a lower absolute epidemic peak would also have led to a flatter epidemic peak. By contrast, the first (BA.1 wave) notifications peak was very sharp in shape. Although the shape of this wave was likely modified by changing reporting requirements and poor national availability of rapid antigen tests, this implies a large epidemic. The mobility extension models we considered improved fit to the seroprevalence estimates, implying that the “shadow” (public-led) lockdown over the 2021-2022 summer was likely part of the explanation for this wave being smaller than expected.^37,38^ We considered that accepting model runs that modestly over-estimated the first seroprevalence estimate but under-estimated the latter two estimates was the optimal balance, as was the case with our chosen primary analysis for parameter inference.

By contrast to mobility, allowing for the additional vaccination programs rolled out during 2022 to modify transmission after Australia had reached close to complete coverage with the two-dose primary courses did not improve model fit. The greatest modelled effect of these programs would be the roll-out of the third dose program for persons aged 16 and above, which reached its greatest rate in February and March 2022 (with all vaccine-related effects lagged to 14 days later). During this period, the modelled effective reproduction number had begun to increase following a nadir, by contrast to its marked decline during January driven by natural immunity following the major BA.1 wave. Therefore, the timing of this program did not help to explain the epidemic profile over a period when the overall change in population immunity was difficult to determine because of the simultaneous implementation of several vaccination programs when immunity from past vaccination was waning.

Our final model structure aimed to balance parsimony against complexity and avoid over-fitting to the available data. Our analysis points to several features that may improve model fit. In addition to the considerations around seroprevalence targets and epidemic size discussed above, we did not allow for a direct effect of vaccination on severe outcomes, for the BA.5 subvariant to be more severe than both BA.1 and BA.2, or for population heterogeneity in spatial structure or immunity beyond our two-category approach. Such configurations and others of interest to the reader can be explored through our interactive notebooks.

## Conclusions

Australia’s 2022 COVID-19 epidemic was characterised by overlapping waves that were driven by Omicron subvariants that exhibited substantial immune-escape properties, with rapidly waning immunity to past infection also likely contributing. The multiple data sources and clear swings in the epidemic profile and reproduction number allowed our beliefs pertaining to several epidemiological parameters to be substantially updated, suggesting a peak case detection rate of 17 to 50% and higher infection fatality rates than observed in a setting with a greater experience with COVID-19.

Our pipeline for infectious disease modelling supports greater understanding of these results through interactive notebooks and online visuals and could constitute a new paradigm for infectious disease modelling.

## Methods

We constructed four candidate models with common underlying characteristics to represent COVID-19 dynamics during the course of 2022 in Australia. Epidemiological details are presented in the repository for this analysis and described in detail in our Supplemental Material, which is algorithmically generated from the code used in model construction to ensure accuracy of documentation. In brief, we built an SEIRS model with chained serial latent and infectious compartments and reinfection from the waned (second S) compartment. To this we added age structure in five-year bands from 0-4 years to 75 years and above. Age stratification determined the initial population distribution, and an age-specific mixing matrix was adapted to the Australian population structure from United Kingdom survey data was applied to capture heterogeneous mixing between age groups.^45^ The model was further stratified into Western Australia (WA) and the other major jurisdictions of Australia to acknowledge the negligible community transmission in WA prior to the re-opening of internal borders to the state. Further stratification was applied to replicate all model compartments into two populations with differing levels of immunity to infection and reinfection, with no transition between these two classes permitted in the base model configuration (without vaccination extension). Three subvariant strains were introduced into the base model through the course of the simulation to represent the BA.1, BA.2 and BA.5 subvariants of Omicron, with incomplete cross-immunity to subsequent strains during the early recovered stage conferred through infection with earlier strains.

The unextended model described in the preceding paragraph was elaborated in two respects. First, the mixing matrix that remains fixed over modelled time in the unextended model was allowed to vary over time, with the location-specific contribution to each cell of the matrix scaled according to metrics sourced from Google’s Community Mobility Reports.^46^

Second, the model was extended to allow that the historical profile of vaccination through 2022 could have influenced rates of infection. Under this alternative analysis, all the model’s initial population was assigned to the non-immune category, with population then transitioning to the partially immune class as new vaccination programs (booster and paediatric) were rolled out through 2022. Vaccine-derived immunity was then allowed to wane, with vaccinated persons returning to a third immunity stratum with the same susceptibility to infection as those who had never received vaccination under these programs.

From these two extensions to the base model, we created four alternative analytical approaches: no additional structure (“none”), mobility extension only (“mob”), vaccination extension only (“vacc”), and both mobility and vaccination extensions (“both”).

Last, we calibrated each of the four candidate models described in the preceding paragraph to publicly available data for three empirical indicators of the COVID-19 epidemic through 2022: the seven-day moving average of national daily time-series for case notifications, the seven-day moving average of national daily time-series for deaths and the results of a nationally representative adult blood donor seroprevalence survey at three key time points in 2022. Equivalent modelled quantities to notifications and deaths were estimated through a convolution approach that allowed a gamma-distributed delay from onset of symptoms (taken as the time of transition from the first to the second serial infectious compartment) to notification or death. The proportion of the population no longer in the original susceptible compartment was further compared to seroprevalence estimates of SARS-CoV-2 exposure, which were adjusted for nucleocapsid test sensitivity and lagged forward by 14 days. Model calibration was then achieved independently for each of the four candidate models using the *PyMC* implementation of the differential evolution Metropolis algorithm “DEMetropolis(Z)”.^14^ All important epidemiological parameter inputs were included in the calibration algorithm, creating a 17-dimensional parameter space for exploration. The approach with only mobility implemented was selected as the primary analysis for parameter inference, largely because of its superior fit to seroprevalence estimates.

## Data Availability

Every aspect of this analysis from input data, through methodological techniques, to model output estimates are publicly available through our GitHub repository.

https://github.com/monash-emu/aust-covid

https://monash-emu.github.io/outputs/aust-covid/

## Author contributions

James Trauer undertook the analysis and drafted the manuscript. Angus Hughes synthesised evidence for parameter inputs and provided other epidemiological advice. David Shipman developed the software pipeline. Michael Meehan and Emma McBryde provided advice on the mathematical basis of the analysis. Romain Ragonnet and Alec Henderson provided advice on calibration algorithms. Romain Ragonnet supervised the project.

