## Supplementary Material for "A Data Science Pipeline Applied to Australia’s 2022 COVID-19 Omicron Waves"

### Supplementary material to analysis of Australia's 2022 COVID-19 epidemic

#### Contents

**1 Approach to analyses** **2**

**2 Base compartmental structure** **2**

**3 Population** **3**

**4 Stratification** **3**

**5 Reinfection** **4**

**6 Mixing** **5**

**7 Mobility extension** **6**

### 1 Approach to analyses

The following document describes the methods used in our analyses of the 2022 SARS-CoV-2 epidemic in Australia. We constructed 4 alternative dynamic transmission models based around the same core features. These are named 'none, mob, vacc, both' and were all based on the features described in Sections 2, 3, 4, 5, 6. Two of the models (mob, both) incorporated additional structure to capture time-varying mobility 7, while two incorporated additional structure for time-varying vaccination effects vacc, both 8, such that these additional features are applied factorially to the base model structure (therefore including one model with neither extension).

Each of the four alternative modelling approaches were then calibrated to the same target data for the 2022 Australian COVID-19 epidemic (see Section 11). The calibration algorithms were also harmonised to the greatest extent possible (see Section 12), although the two analysis approaches that included structure for vaccination required a different parameter to be substituted for the parameters used in the analyses not incorporating this structure (as described below). These four approaches were then compared with regards to their fit to the target data, with the 'mob' analysis (with structure for mobility but not for vaccination) found to achieve the highest likelihood. As a result, this approach to analysis was selected as the primary analysis (see Section 13)). This approach was used for the further analyses, including parameter inference (e.g. Section 14).

### 2 Base compartmental structure

This section describes the model features that were common to the model used for all four analyses. We used the summer framework to construct a compartmental model of COVID-19 dynamics. The base model consists of 11 states, representing sequential epidemiological conditions with regards SARS-CoV-2 infection and COVID-19 disease (susceptible, recovered, waned, latent_0, latent_1, latent_2, latent_3, infectious_0, infectious_1, infectious_2, infectious_3). Each of the sequentially numbered infectious compartments contribute equally to the force of infection.

The infection process moves people from the susceptible compartment to the latent_0 compartment (i.e. the first latent compartment), under the assumption of frequency-dependent transmission. Following infection, infected persons proceed to transition through a series of 4 latent compartments. These are chained in sequence, with infected persons transitioning sequentially from compartment 0 through to compartment 3. To achieve the same mean sojourn time in the composite latent stage, the rate of transition between successive latent compartments and of exiting the the last latent compartment are multiplied by the number of serial compartments (i.e. 4). As persons exit the final latent compartment, they enter the first infectious compartment. As for the latent compartments, the infectious compartments are also chained in series, with a total of 4 again chained together in sequence. As for the latent compartments, each transition rate is multiplied by 4. As persons exit the final infectious compartment, they enter the recovered compartment. An Erlang-distributed infectious and latent duration is consistent with epidemiological evidence that the serial interval [19] and generation time [21] are often well represented by a gamma distribution, with multiple past modelling studies choosing a shape parameter of four or five having been previously used to fit this distribution [7, 8]. A waned compartment is included in the model to represent persons who no longer have natural immunity from past SARS-CoV-2 infection. As these persons lose their infection-induced immunity, they transition from the recovered compartment to the waned compartment at a rate equal to the reciprocal of the natural immunity period parameter (See Table 2).

### 3 Population

Each simulation is run from 1$^{st}$ of July 2021 to 1$^{st}$ of October 2022. For estimates of the Australian population, data were downloaded from the Australian Bureau of Statistics website on 1$^{st}$ of March 2023 [40] (sheet 31010do002_202206.xlsx). Minor jurisdictions other than Australia's eight major state and territories (i.e. Christmas island, the Cocos Islands, Norfolk Island and Jervis Bay Territory) are excluded from these data. These much smaller jurisdictions likely contribute little to overall COVID-19 epidemiology and are also unlikely to mix homogeneously with the larger states/territories. The populations of states other than Western Australia (WA) were summed to obtain the population of the second 'other states' spatial patch of the model. The population estimates for all 5-year age brackets from 75-79 upwards were summed to obtain the 75 and above age group estimates (Figure 1). The simulation starts with 25.683 million fully susceptible persons, with the infection process triggered through subsequent strain seeding as described below.

### 4 Stratification

#### 4.1 Age

We stratified all compartments of the base model described above (Section 2) into sequential age brackets comprising 5-year bands from age 0 to 4 through to age 70 to 74, with a final age band to represent those aged 75 and above. These age brackets were chosen to match those used by the POLYMOD survey [30] and so fit with the mixing approach implemented (Section 6). The population distribution by modelled age group was obtained from the Australian Bureau of Statistics data introduced previously (Section 3, Figure 1). Ageing between sequential bands was not permitted given the short time window of the simulation.

#### 4.2 Omicron Sub-variants

We stratified all compartments other than susceptible according to strain (recovered, waned, latent_0, latent_1, latent_2, latent_3, infectious_0, infectious_1, infectious_2, infectious_3), replicating all of these compartments to represent the various major Omicron sub-variants relevant to Australia's 2022 epidemic, namely: BA.1, BA.2, BA.5. This was implemented using the summer library's 'StrainStratification' class.

In Australia, three sequential but overlapping epidemic waves were observed, which were attributable to the subvariants: BA.1, BA.2, BA.5. Each strain (including the starting BA.1 strain) was seeded through a triangular step function that introduces new infectious persons into the latent_0 compartment over a fixed seeding duration defined by a single variable Seed time parameter (i.e. applied to all subvariants) at a peak rate defined by a single Seed rate parameter (also applied to all subvariants). The time of first emergence of each strain into the system was defined by a separate emergence time parameter for each modelled subvariant strain. This emergence time was varied across credible intervals, with model outputs then compared against available Australian sequencing data (see Figure 25).

##### 4.3 Spatial

All model compartments previously described were further stratified into strata to represent Western Australia (WA) and 'other' to represent the remaining major jurisdictions of Australia. This approach was adopted to remove WA's contribution to community transmission prior to WA reopening its borders to the rest of the country on 2022-03-03 00:00:00. To achieve this effect, transmission in WA was initially set to zero, and subsquently scaled up to being equal to that of the other jurisdictions of Australia over a period that governed by a calibrated parameter ('WA reopen period').

##### 4.4 Heterogeneous susceptibility

All (multiply stratified) compartments introduced above were further stratified into 2 strata with differing levels of susceptibility to infection in the two analyses without extension for vaccination. For these two analyses, a calibrated parameter was used to represent the proportion of the population with immunological protection against infection. A second calibrated parameter (Immune protection) was then used to quantify the relative reduction in the rate of infection and reinfection for those in the stratum with reduced susceptibility. This approach was adopted because some population heterogeneity in susceptibility may have been introduced through a proportion of the population having greater protection through vaccination, boosting or other intrinsic individual variation that would not otherwise be captured under the assumptions inherent in our compartmental model. By contrast to the vaccination extension approach, no flows between these strata were applied, such that the proportion of the population in each of the two strata remains fixed over time.

#### 5 Reinfection

We modelled reinfection from both the recovered and waned compartments, which we term 'early' and 'late' reinfection respectively. In the case of early reinfection, this is only possible for persons who have recovered from an earlier circulating sub-variant. That is, early BA.2 reinfection is possible for persons previously infected with BA.1, and early BA.5 reinfection is possible for persons previously infected with BA.1 or BA.2, while other reinfection processes are not permitted. The parameter governing the degree of immune escape is determined by the infecting variant was estimated separately for BA.2 and BA.5. Therefore, the rate of reinfection is equal for BA.5

***Figure 1:*** **Stacked Australian population sizes implemented in the model.** Western Australia (blue bars), aggregate of remaining major jurisdictions of Australia (red bars).

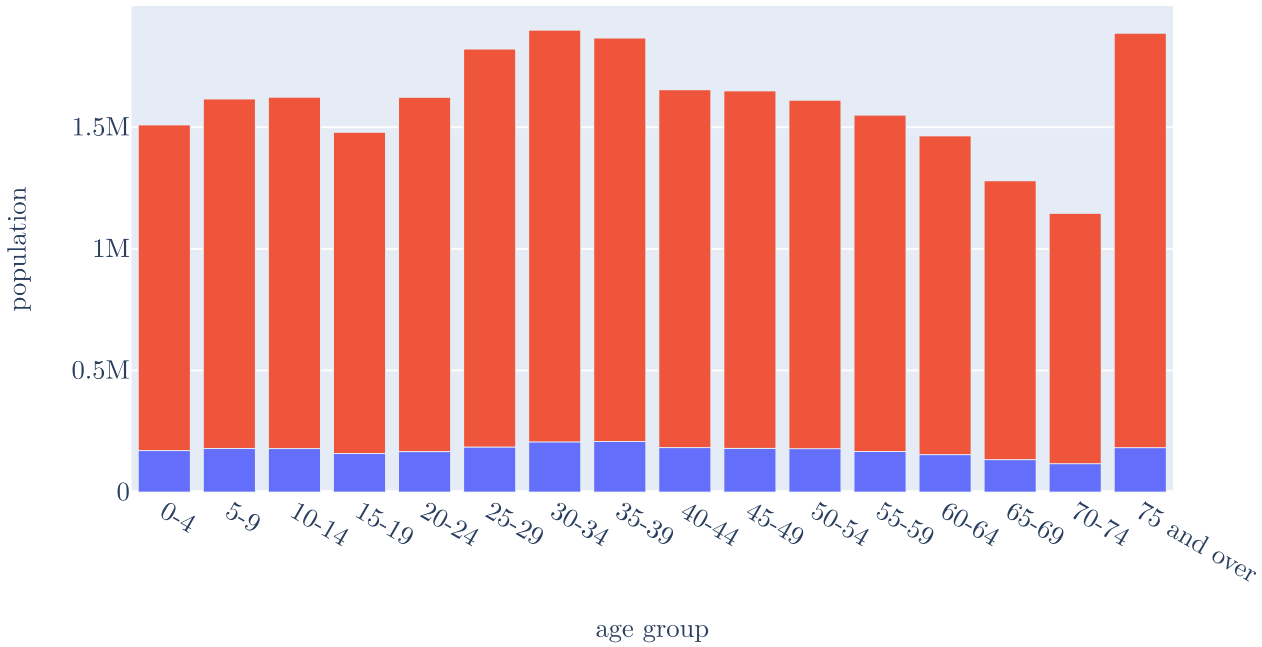

reinfecting those recovered from past BA.1 infection as for those recently recovered from past BA.2 infection.

For late reinfection, all natural immunity is lost for persons in the waned compartment, such that the rate of reinfection for these persons is the same as the rate of infection for fully susceptible persons. As for the process of first infection, all reinfection processes transition individuals to the latent compartment corresponding to the infecting strain.

### 6 Mixing

Raw, location-specific social contact matrices from the POLYMOD study for Great Britain (Figure 3) were adjusted to account for the differences in the age distribution betweem the Australian population distribution in 2022 (Figure 1) and the population of Great Britain in 2000 (Figure 2). The raw matrices were adjusted by taking the dot product of the location-specific unadjusted matrices and the diagonal matrix containing the vector of the ratios between the proportion of the British and Australian populations within each age bracket as its diagonal elements. In analyses without contact scaling for mobility, the resulting adjusted matrices summed over location (Figure 4) were implemented as fixed rates of contact between each possible pair of age groups.

***Figure 2:*** **United Kingdom population sizes used in matrix weighting.**

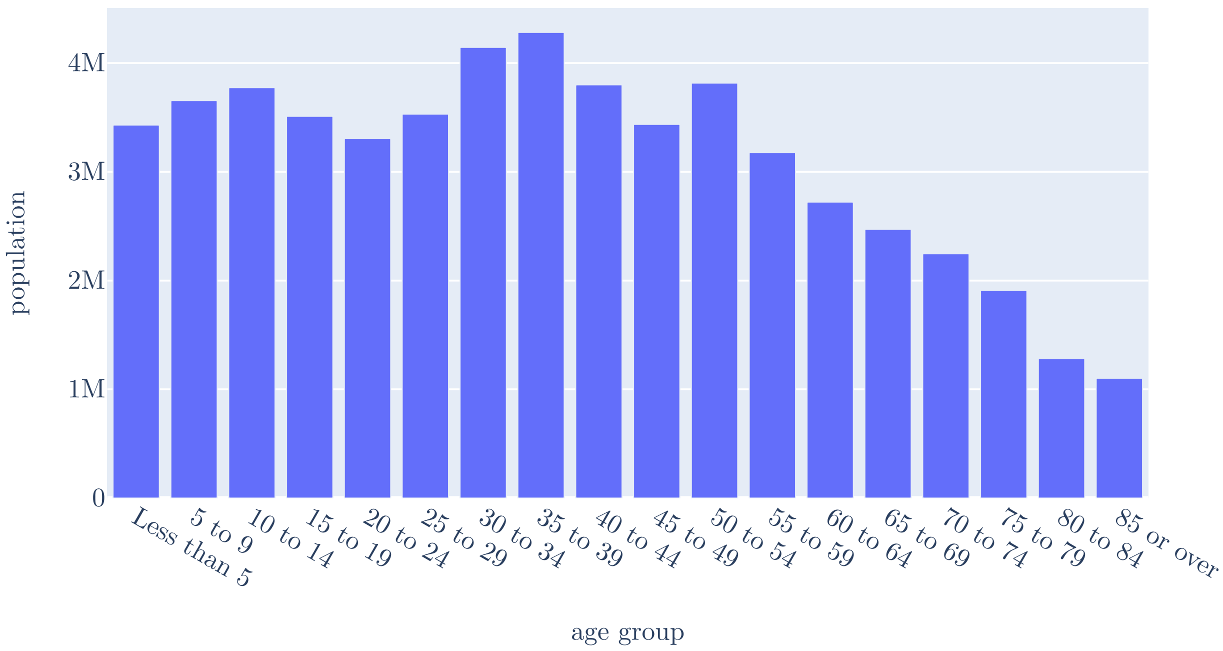

To align with the methodology of the POLYMOD study [30] we sourced the 2001 UK census population for those living in the UK at the time of the census from the Eurostat database.

### 7 Mobility extension

#### 7.1 Application

The two scaling functions developed in the previous section were used to adjust rates of contact over time in the two time-varying matrix locations. These were summed with the two static locations to obtain the final matrix. Examples of the final effect of the matrix scaling function on the dynamic mixing matrices are presented in Figure 5.

#### 7.2 Data processing

We undertook an alternative analysis in which estimates of population mobility were used to scale transmission rates.

Raw estimates of Australian population mobility were obtained from Google with 2021 and 2022 data concatenated together (Figure 6). Values for Western Australia were extracted separately

***Figure 3:*** **Raw contact rates obtained from POLYMOD surveys for the United Kingdom.** Number of contacts per day by respondent age group (row), contact age group (column) and location (panel).

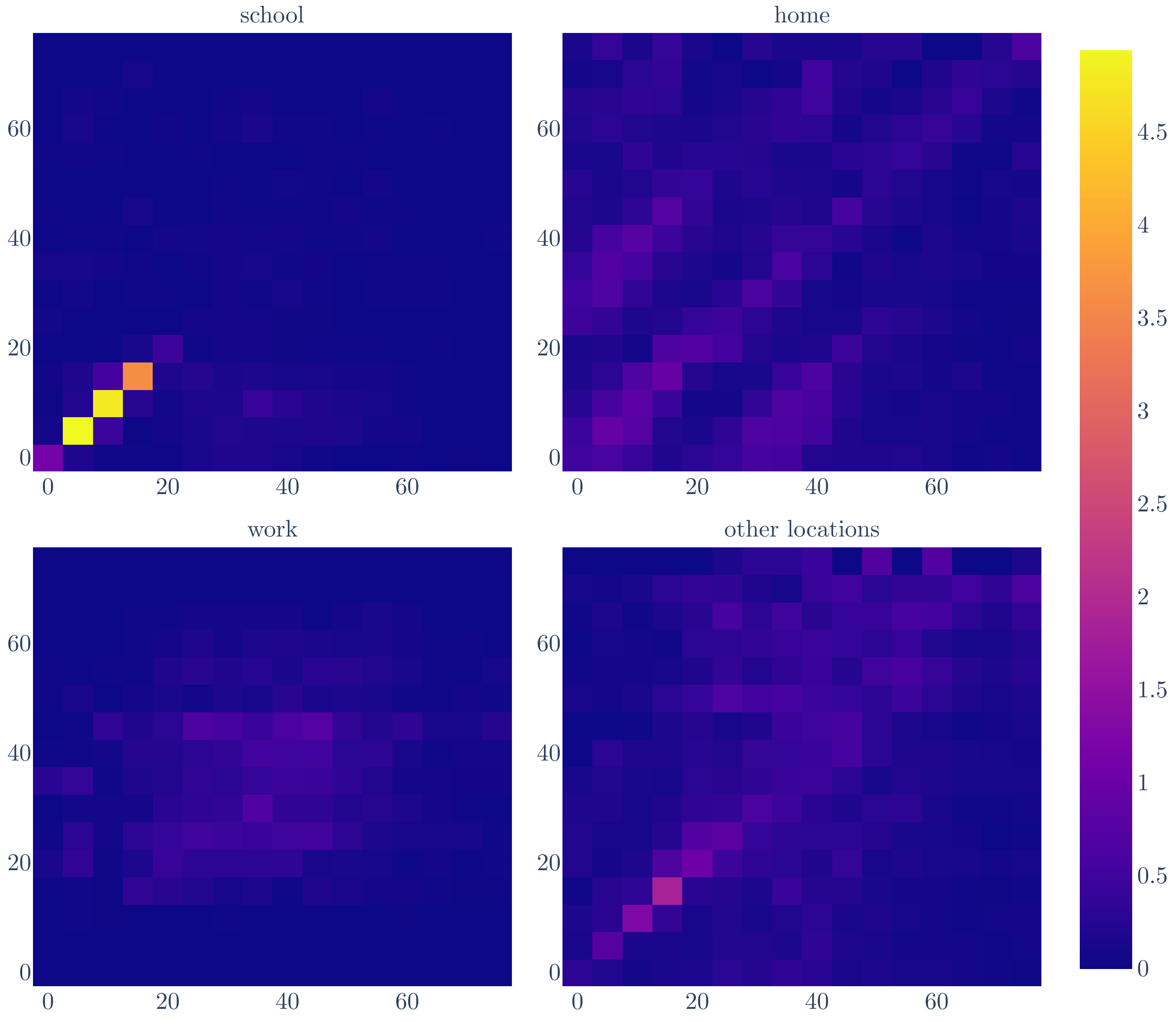

***Figure 4:*** **Contact rates after adjustment to the Australian population distribution.** Number of contacts per day by respondent age group (row), contact age group (column) and location (panel).

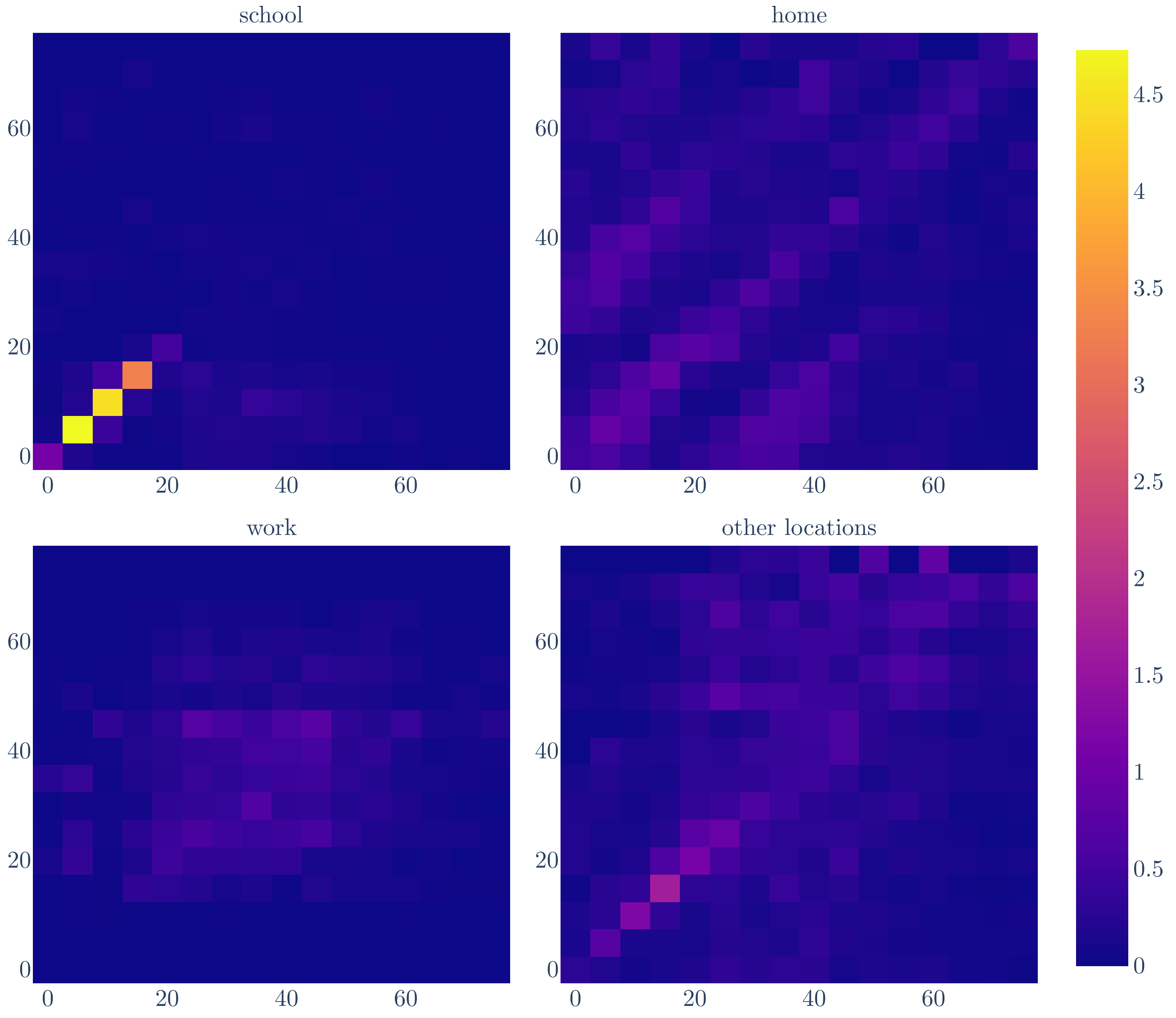

***Figure 5:*** **Dynamic mixing matrices.**

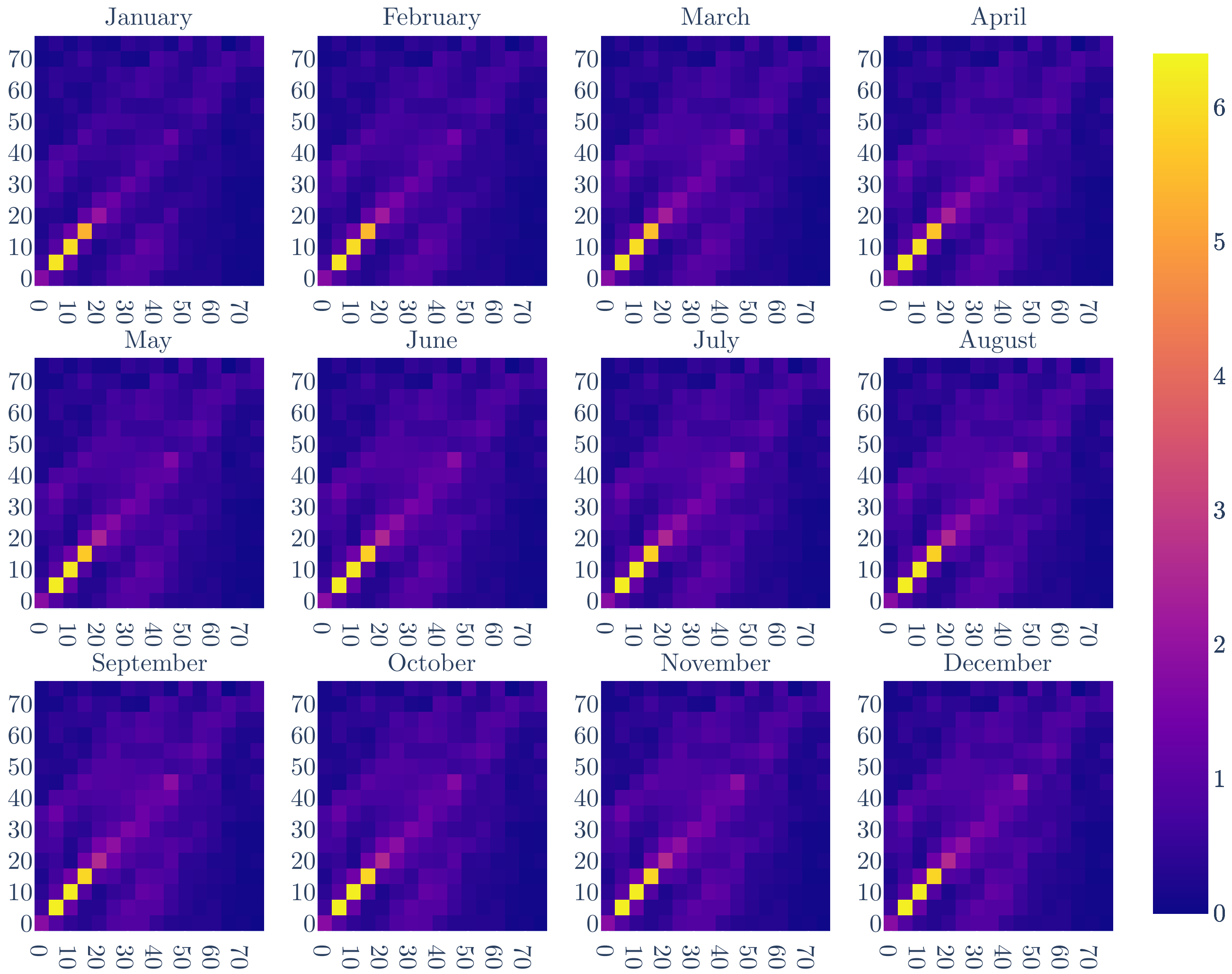

| | Other locations | Work |
|---|---|---|
| Retail and recreation | 0.34 | 0.0 |
| Grocery and pharmacy | 0.33 | 0.0 |
| Parks | 0.0 | 0.0 |
| Transit stations | 0.33 | 0.0 |
| Workplaces | 0.0 | 1.0 |
| Residential | 0.0 | 0.0 |

***Table 1:*** **Mobility mapping..**

***Figure 6:*** **Raw state-level mobility obtained from Google.** Locations: Retail and recreation (green), grocery and pharmacy (purple), parks (orange), transit stations (yellow), workplaces (blue), and residential (red).

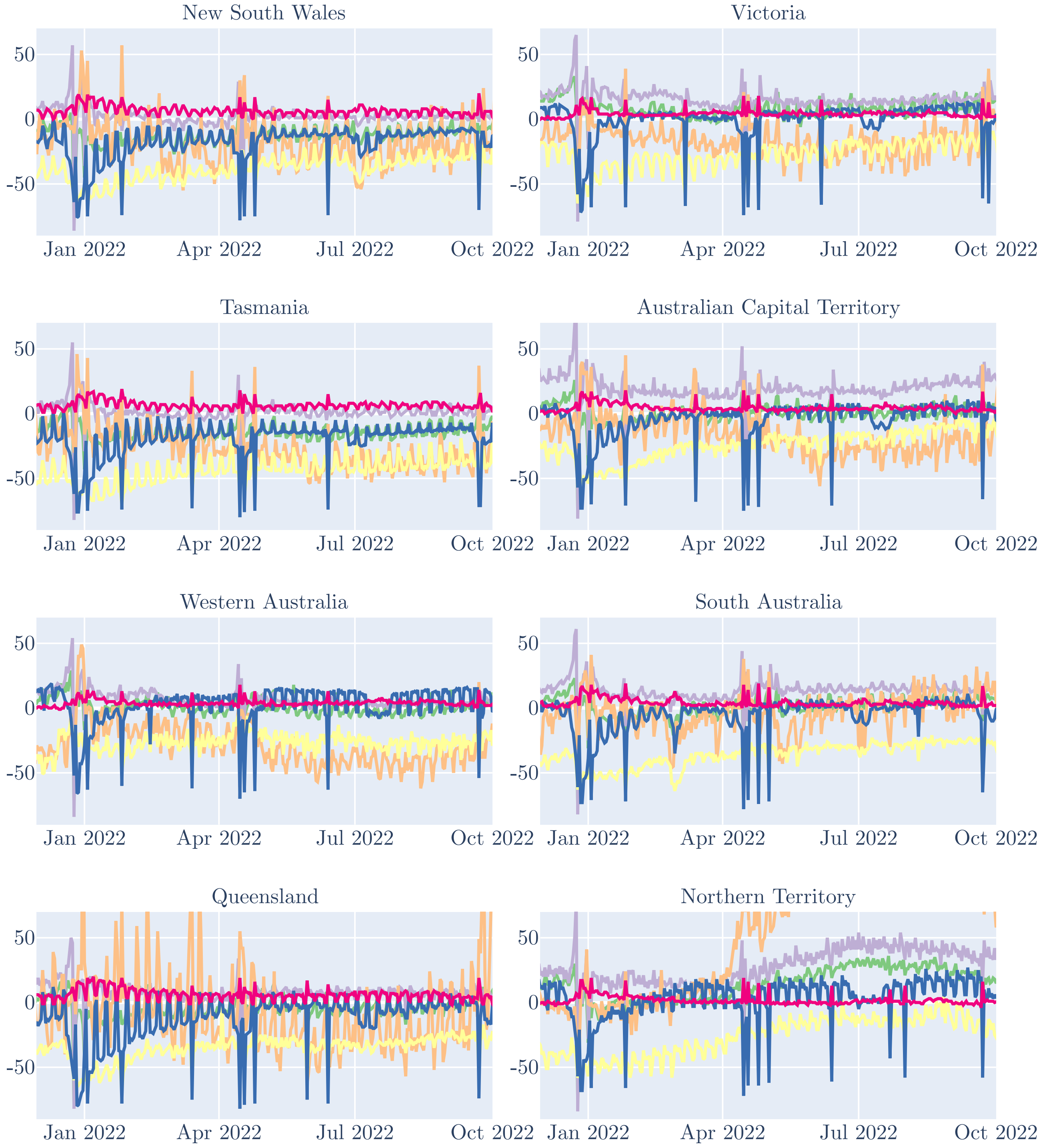

***Figure 7:*** **Processed mobility for model.** Work mobility (yellow line), smoothed work mobility (blue line), squared smoothed work mobility (red line), averaged other locations mobility (green line), smoothed averaged other locations mobility (purple), and squared smoothed averaged other locations mobility (orange).

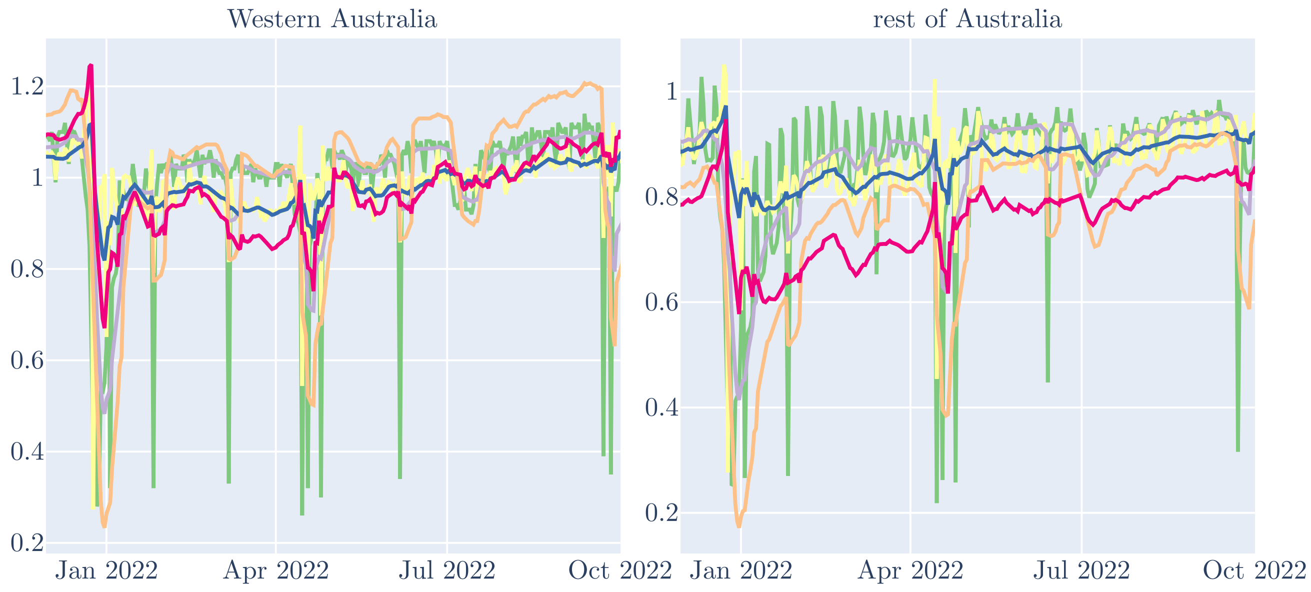

from the pooled data, while the data for the remaining states were linked to the same population size data as used to set the compartment sizes for the model (Figure 1). These population values were then used as weights to calculate weighted national averages for population mobility by each Google 'location' (retail and recreation, grocery and pharmacy, parks, transit stations, workplaces, residential).

Values were then converted from the reported percentage change from baseline to the proportional change relative to baseline, to obtain contact scaling factors. Next, we used the mapping algorithm displayed in Table 1 to map from Google's reported 'locations' to the contact locations of the model's mixing matrix. Next, we took the 7-day moving average to smooth the often abrupt shifts in mobility, including with weekend and public holidays. Last, we squared the relative variations in mobility to account for the effect of reductions in visits to specific locations for both the infector and the infectee of the modelled social contacts. These sequentially processed functions of time are illustrated in Figure 7.

### 8 Vaccination extension

#### 8.1 Rationale

Although Australia's population was relatively unexposed to SARS-CoV-2 infection and so had little natural immunity at the start of our simulation period, the population had extensive vaccination-derived immunity across eligible age groups. This is illustrated in Figure 8, which shows that most age groups had reached very high coverage with a second-dose of vaccine by early 2022. As such, we considered that the continuing roll-out of second doses were very unlikely to have substantially modified the epidemic, particularly given the questionable impact of such vaccination programs on onward transmission.

We therefore considered programs that were rolled out over the course of 2022 for their potential impact on transmission. As shown in Figure 9, we considered that the most likely programs to have had an effect on transmission through 2022 were the fourth dose 'winter booster' program, and the primary course (completing second doses) program for children aged 5 to 11 years. The heterogeneous immunity stratification of the base model was utilised to consider the impact of these programs on community transmission, as described in the following section.

#### 8.2 Application

Using the various reporting streams from which the vaccination data were derived (illustrated as different colours in Figure 9), we calculated the total number of persons vaccinated. Next, we converted this to a proportion using the population denominators supplied by the Commonwealth in the same dataset. We then calculated the proportion of the population previously unvaccinated under each of these programs and divided by the time interval over which this occurred to calculate the rate at which these population groups received vaccination (Figure 10). These were then applied as unidirectional flows taht transitioned persons from the unvaccinated stratum to the vaccinated stratum. For this extended model configuration, a third stratum was added to the immunity stratification. The reported coverage and coverage lagged by 14 days are compared against the modelled population distribution across the three immunity strata in Figure 11.

***Figure 8:*** **Full vaccination coverage** Second ('full') dose roll-out by age group. Number of persons receiving second dose (upper panel) and proportion of population having received second dose (lower panel). Age groups coloured from cyan (12 to 15 years-old) to purple (95+ years-old).

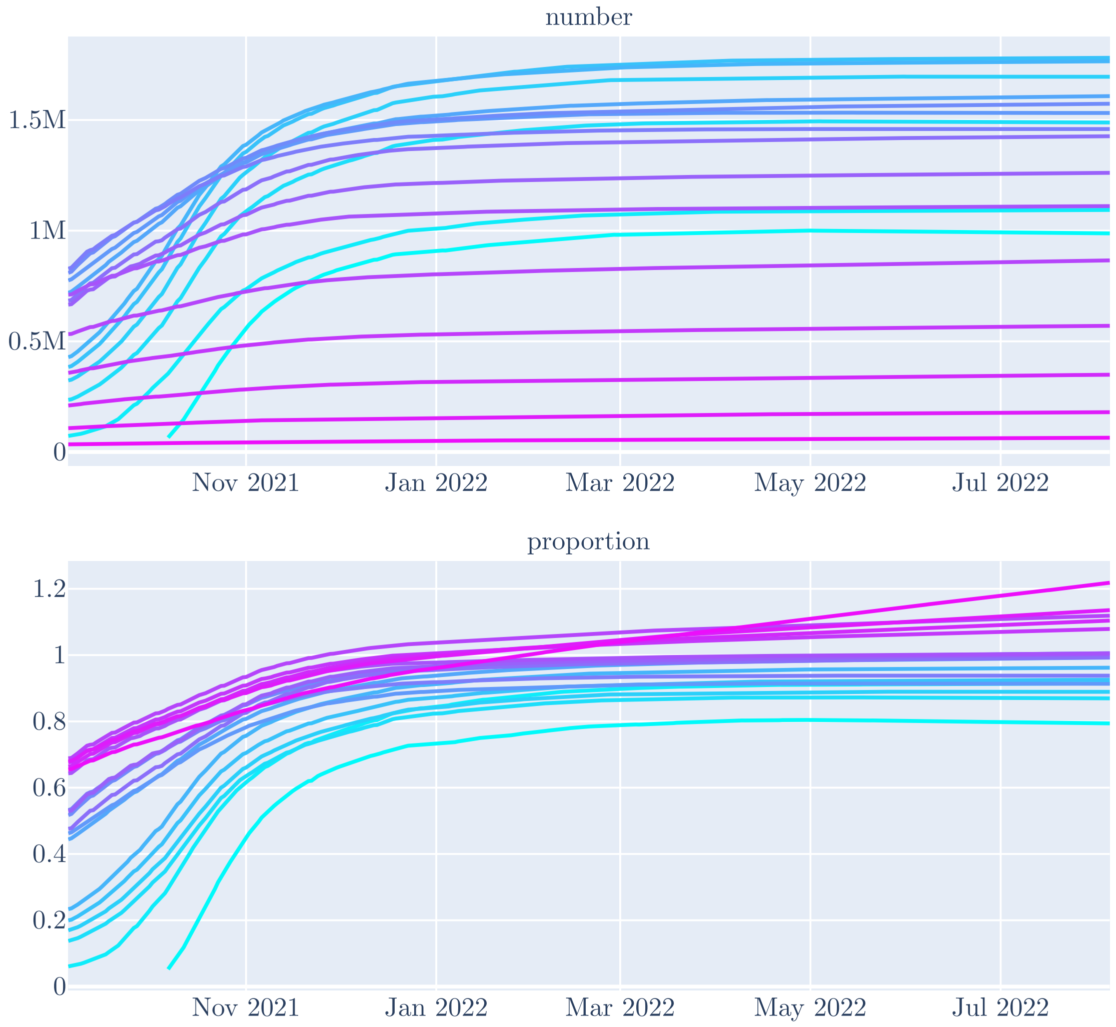

***Figure 9:*** **Vaccination coverage by subsequent programs.**

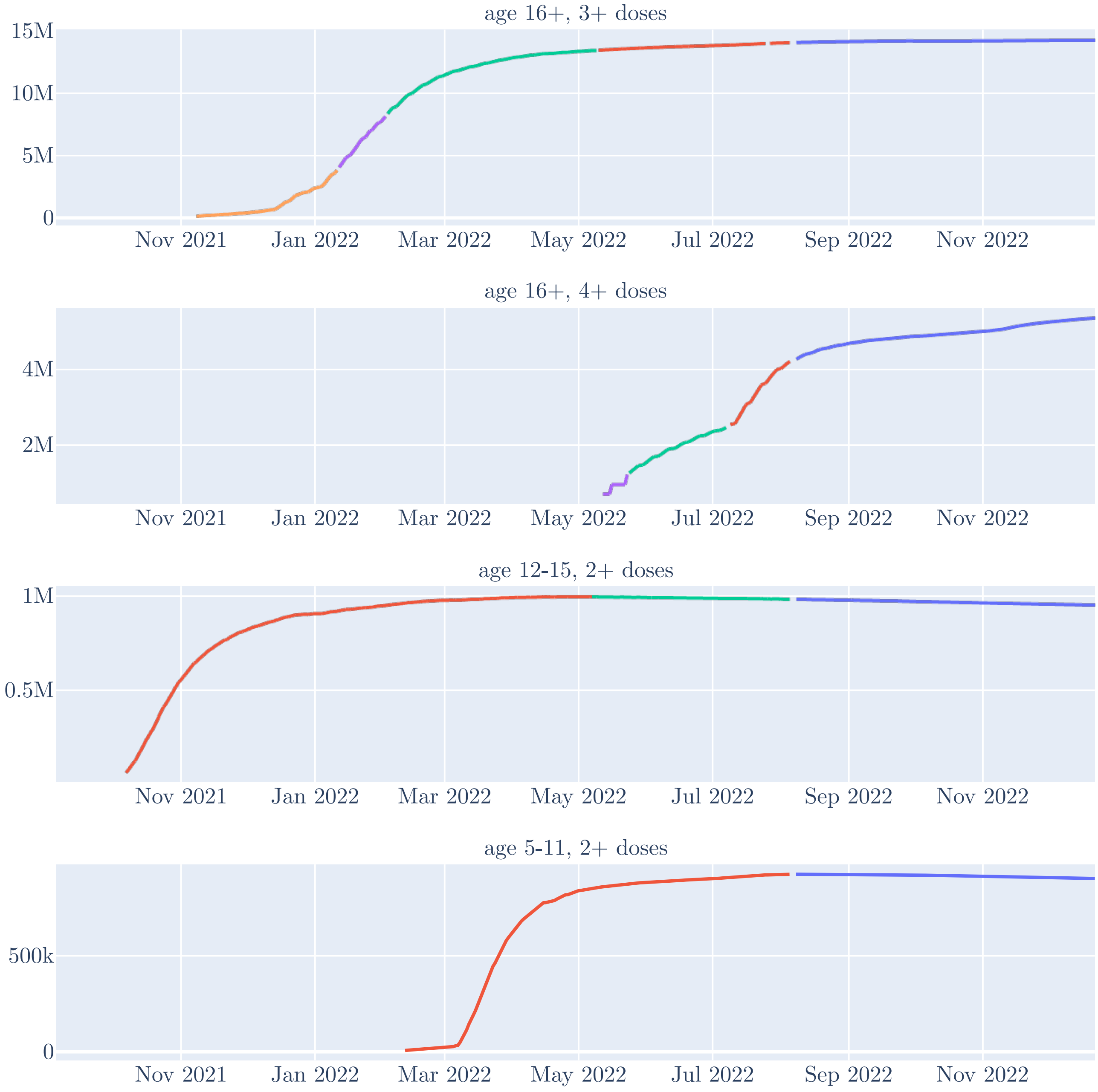

***Figure 10:*** **Vaccination implementation.** Booster program for persons aged 16 and above (red line), primary vaccination course for persons aged 5 to 11.

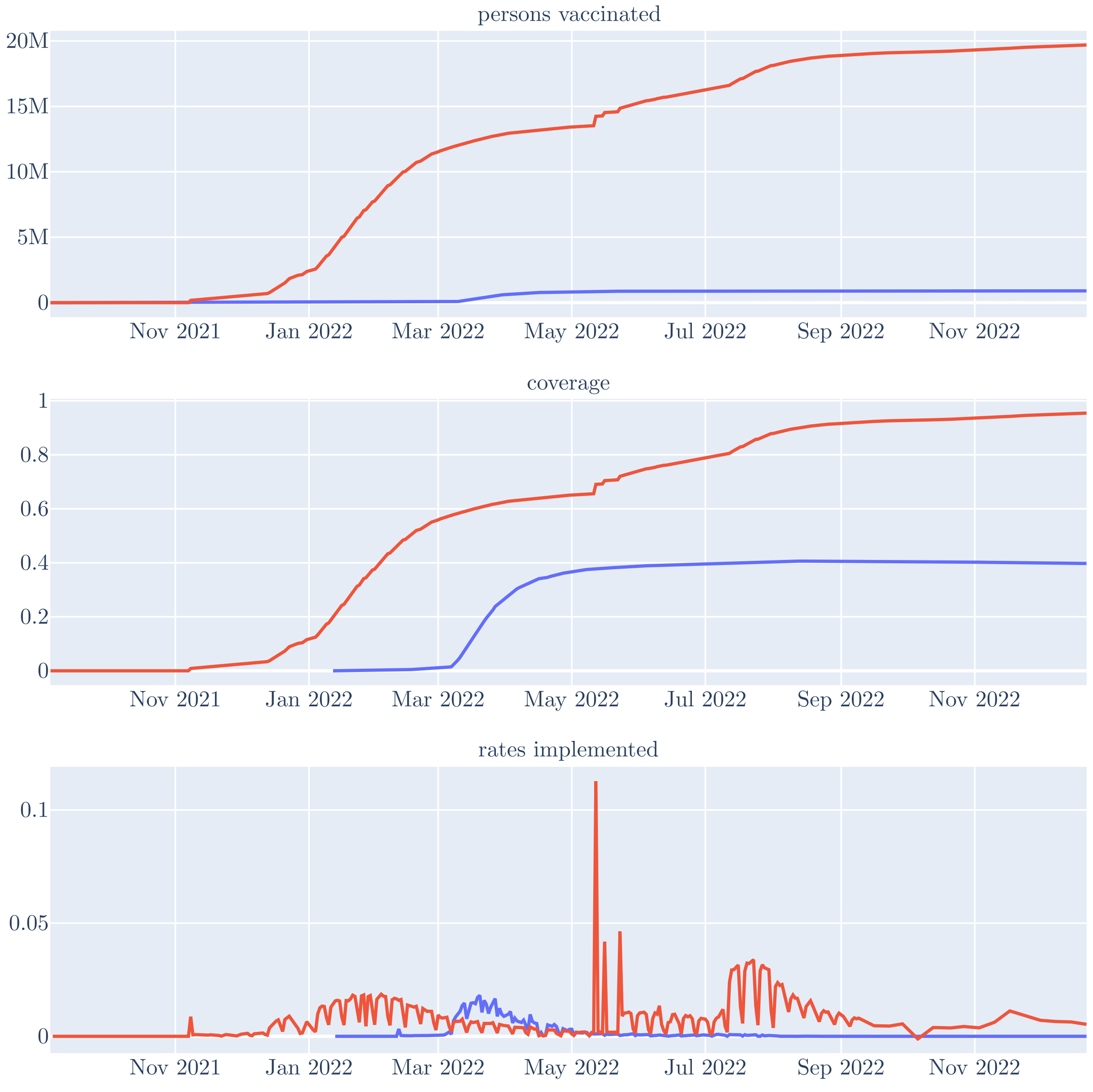

***Figure 11:*** **Comparison of reported to modelled vaccination status distribution.** Reported vaccination coverage by program (dashed black line), and lagged vaccination coverage (dotted black line). Coloured areas represent distribution of population by vaccination status under vaccination extension: not yet vaccinated under program (green), recently vaccinated under program (red), and vaccinated under program but protective effect lost (blue).

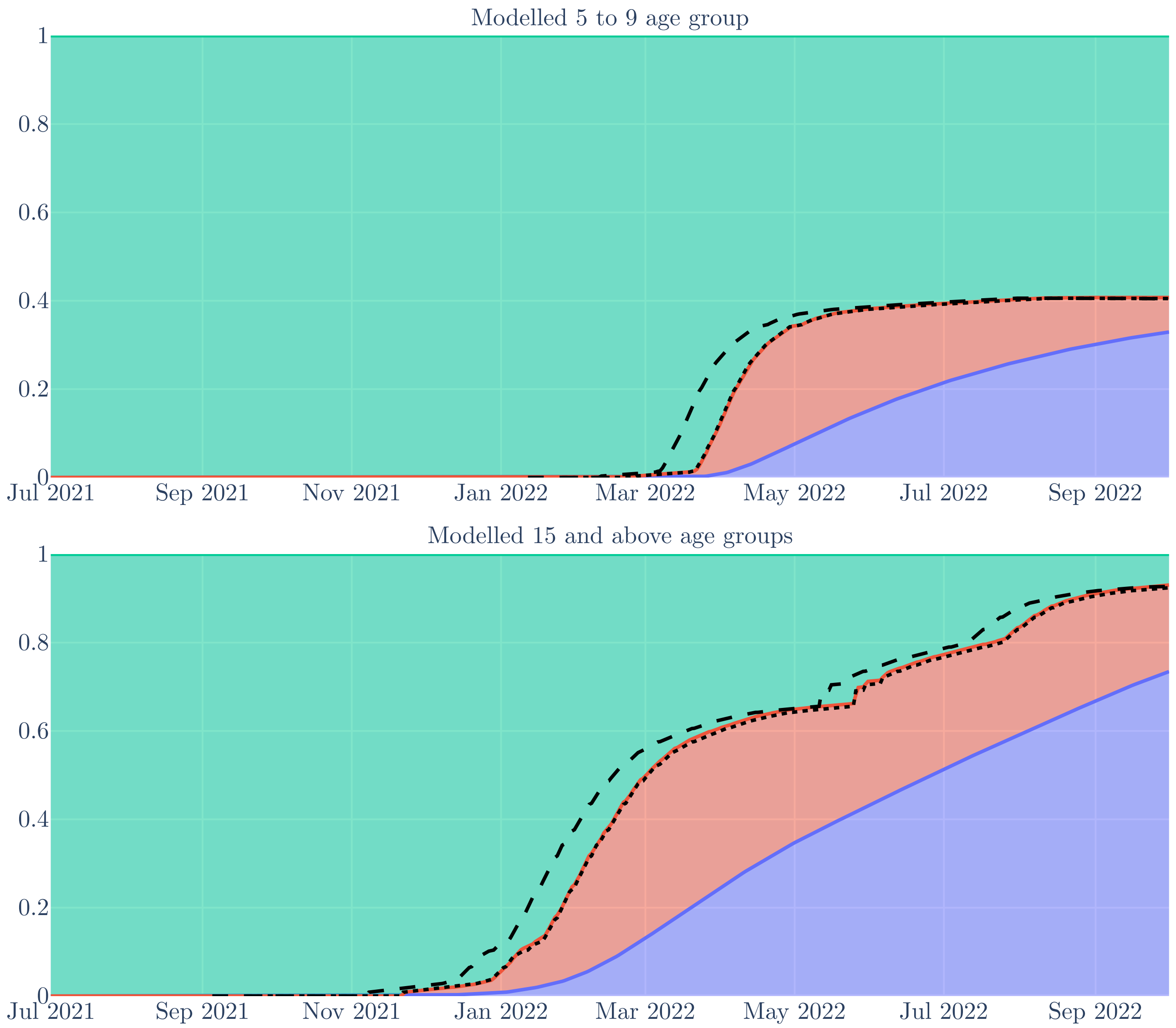

### 9 Outputs

Results from an example run are presented in Figure 14, Figure **??** and Figure 15, and model construction is described in the following sections.

#### 9.1 Notifications

Age group, strain-specific, and overall incidence of SARS-CoV-2 (including modelled episodes that would never have been detected) were tracked. This incidence was not used explicitly in the calibration process, but tracking this process was necessary for the calculation of several other model outputs, as described below. The point at which new cases contribute to incidence is taken as the time of symptom onset in those infected persons who do develop symptoms. To account for the observation that infectiousness is often present for a short period prior to the onset of symptoms, we estimated incidence as the transition from the first to second chained sequential infectious compartment.

The extent of community testing following symptomatic infection is likely to have declined over the course of 2022. To understand these trends, we first considered data from the Australian Bureau of Statistics Household Impacts of COVID-19 surveys, which were undertaken periodically throughout 2022 with standardised questions at each round (downloded on the $12^{\text{th}}$ of June 2023). These surveys reported on several indicators, including the proportion of households reporting a household member with symptoms of cold, flu or COVID-19, and the proportion of households reporting a household member that has had a COVID-19 test. We considered that the ratio of the proportion of households reporting having undertaken COVID-19 tests to the proportion of households with a symptomatic member provided the best available indicator of the decline in testing over this period (Figure 12). We define the case detection rate (CDR) as the proportion of all incident SARS-CoV-2 infection episodes (including asymptomatic and undetected episodes) that were captured through the surveillance data we used in calibration.

In calibration, we varied the starting case detection rate at the time of the first survey through plausible ranges, which declined thereafter according to the survey estimates described. The relationship between CDR and our calculated ratio of testing in households to symptomatic persons in households was defined by an exponential function to ensure that the CDR remained in the domain [0, 1], dropping to zero when household testing reached zero and approaching one were household testing to approach very high levels. Specifically, the case detection rate when the ratio is equal to $r$ with starting CDR of $s$ is given by $s = 1 - e^{-p \times r}$. The value of $p$ is calculated to ensure that $s$ is equal to the intended CDR when $r$ is at its starting value. This approach led to an estimated fall in the case detection ratio by a factor of two over the first half of 2022. This is consistent with our intuition and with an epidemiological modelling analysis from New Zealand over a similar time period that integrated surveillance and waste water data [46].

#### 9.2 Deaths

Calculation of the COVID-19-specific deaths followed an analogous approach to that described for notifications, except that there is no assumption of partial observation and age-specific infection fatality rates are used (as described in Section 10.1). For each age group, we first multiplied the age-specific incidence by the infection fatality rate for that group, and then adjusted this rate according to the relative infectiousness of the BA.2 subvariant in the case of this strain (with the 'ba2_rel_ifr' parameter). Next, we convolved this rate with a gamma distribution for the delay from

***Figure 12:*** **Construction of CDR function.** Raw survey values from Household Impacts of COVID-19 surveys (upper panel), with proportion of households reporting symptoms (blue line), proportion of households positive for COVID-19 (green line) and proportion of hoseholds reporting testing for COVID-19 (red line). Ratio of testing to reporting symptoms (blue line, lower panel).

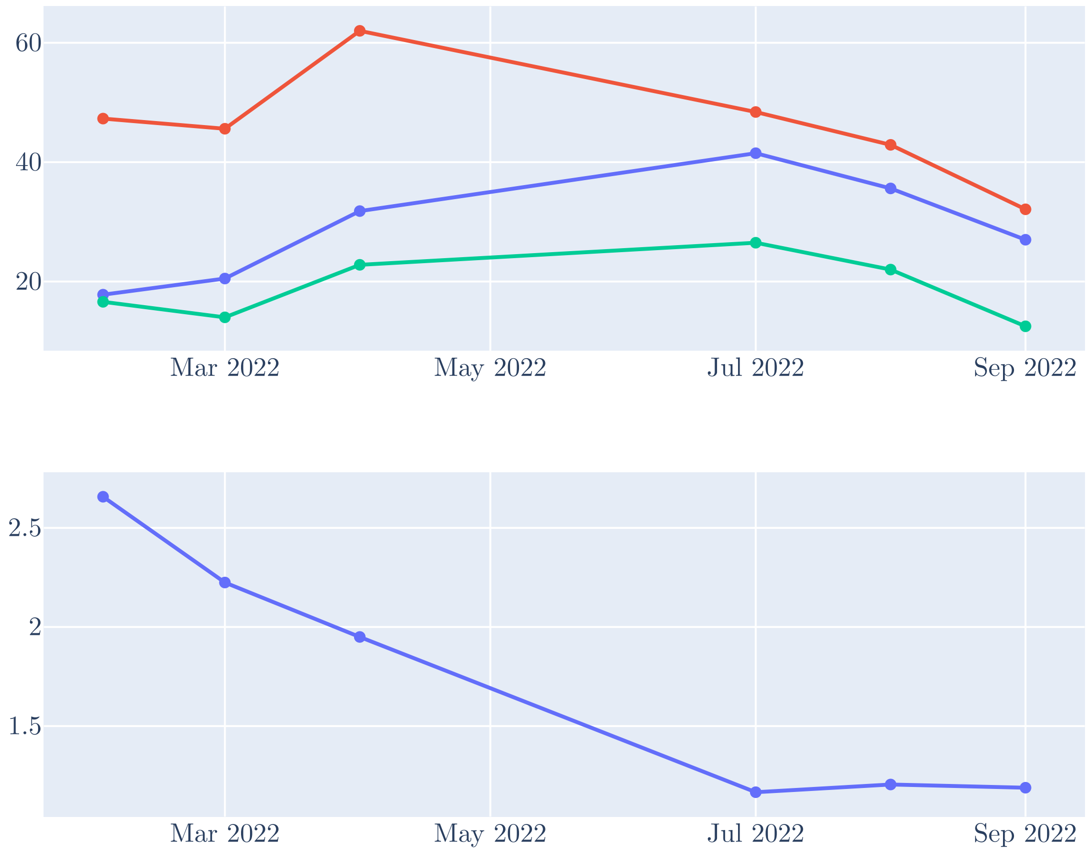

***Figure 13:*** **Examples of the modelled effect of various starting CDR proportion parameters.** Modelled case detection ratio over time for 12 randomly selected parameter draws from calibration algorithm.

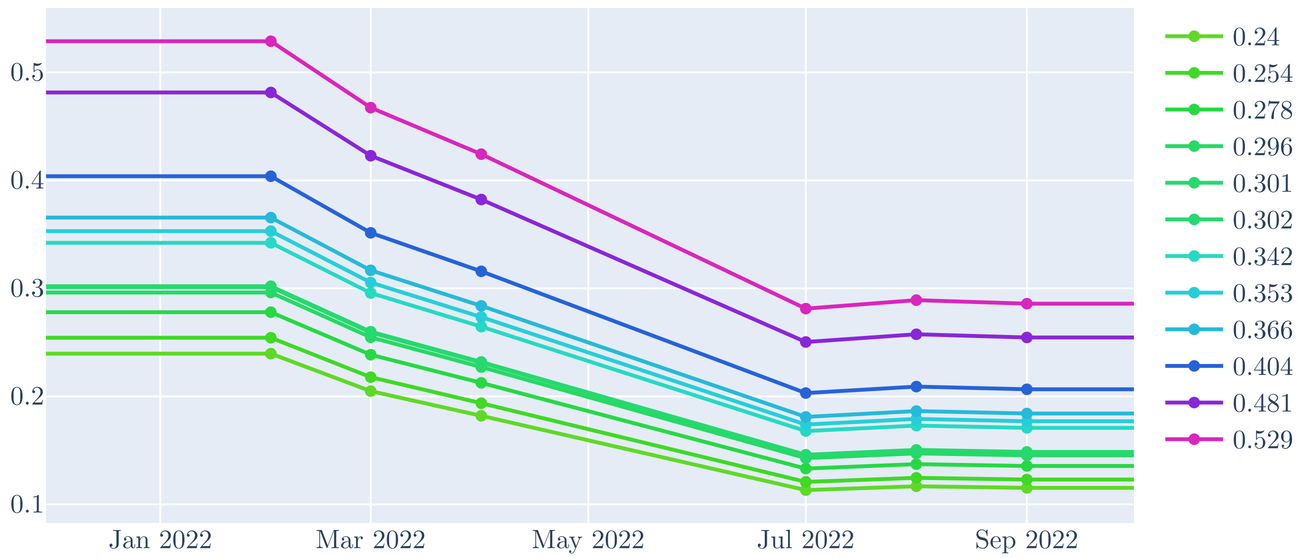

***Figure 14:*** **Outputs from single model run**

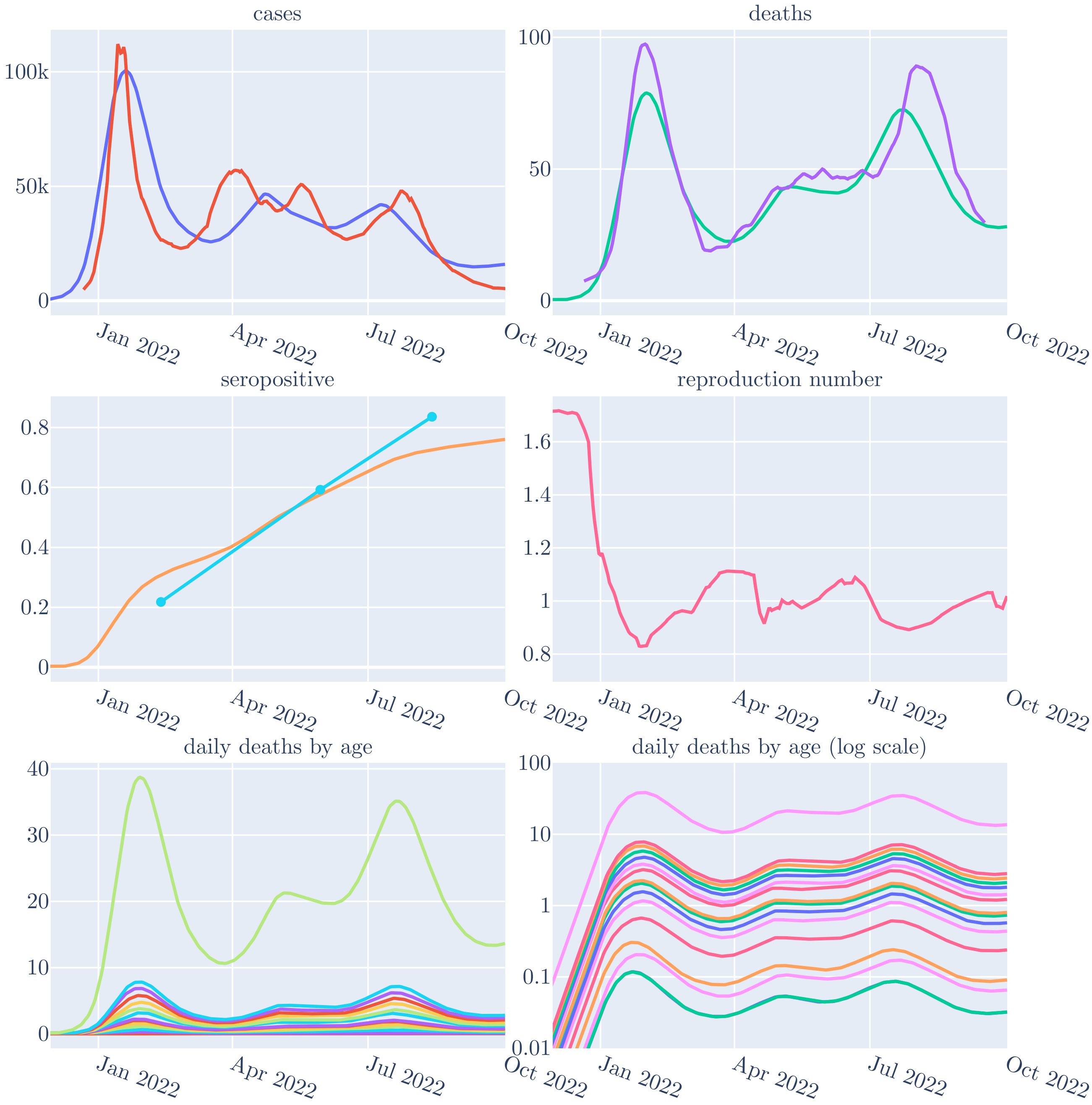

***Figure 15:*** **Simulated infection processes for maximum likelihood results** Colour shows infection with BA.1 (greens), BA.2 (blues) and BA.5 (purples). Shading depth shows infection process, with initial infection (dark), early reinfection (intermediate darkness), late reinfection (light).Note early reinfection with BA.1 does not occur to a significant extent.

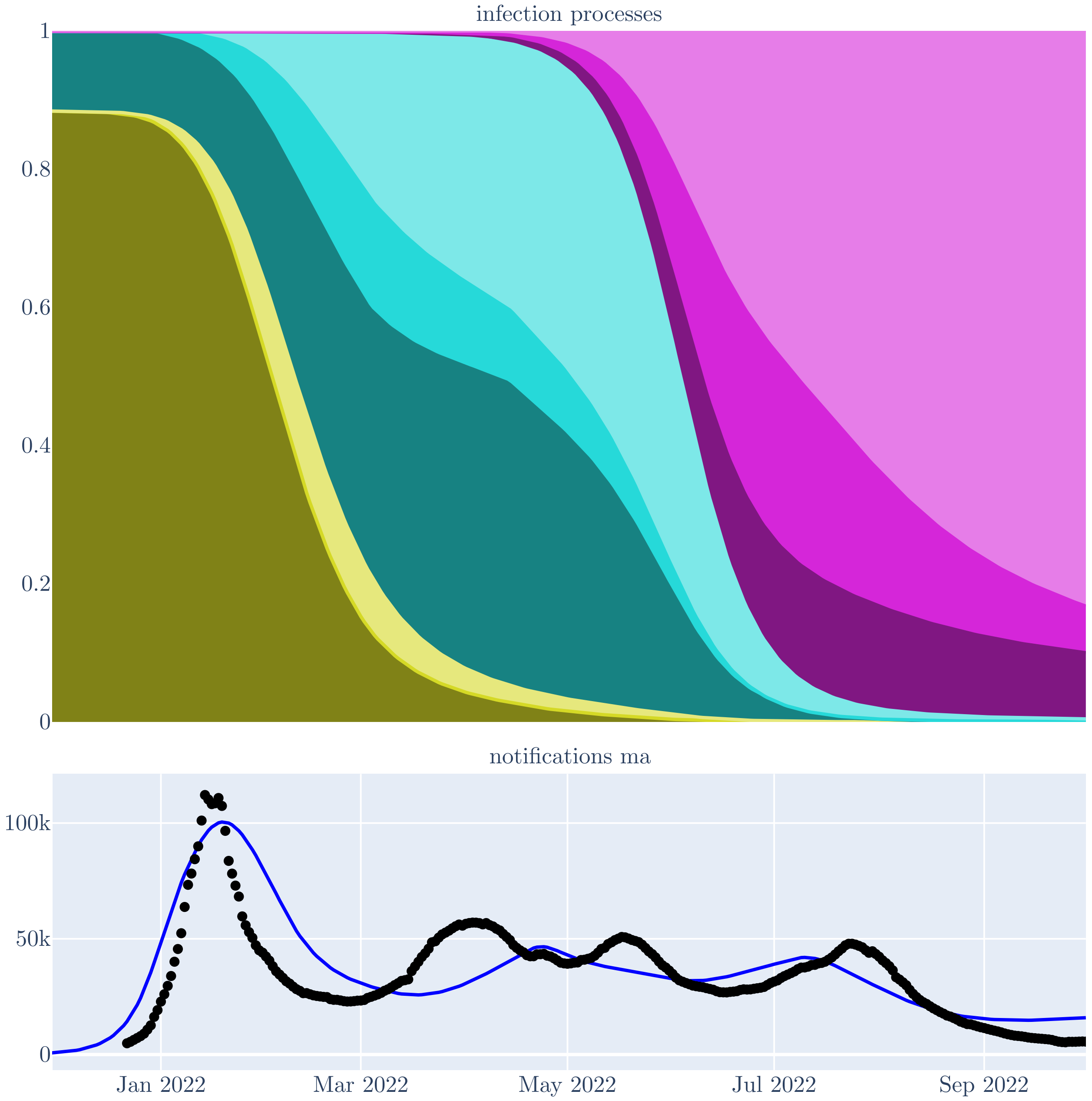

symptom onset to death to obtain the daily rate of deaths for each age group, and lastly summed over age groups.

##### 9.3 Seroprevalence

The proportion of the overall population in any compartment other than the susceptible compartment among those aged 15 years and above was used to estimate the adult 'seropositive' proportion, in order to align with reported seroprevalence estiamtes.

##### 9.4 Sub-variants

Proportional prevalence of each Omicron sub-variant was tracked as the proportion of the population currently in any of the infectious compartments that is infected with the modelled strain of interest (noting that simultaneous infection with multiple strains is not permitted).

##### 9.5 Reproduction Number

The time-varying effective reproduction number was calculated as the rate of all infections (including both first infection and reinfection) divided by the prevalence of infectious persons (i.e. in the infectious compartments) multiplied by the duration of the infectious period. This quantity was tracked for illustrative purposes, but did not contribute to any modelled processes or to likelihood calculation.

#### 10 Parameters

All epidemiologial parameters, including those used in the calibration algorithm are presented in Table 2. In addition to these epidemiological parameters, the dispersion parameter for the negative binomial distribution used when calculating the case time-series and death time-series contributions to the likelihood were included in our calibration algorithm (See Section 11.4). The approach to estimating the age-specific infection fatality rate for each modelled age group is described in 10.1. All epidemiologically significant model parameters were included as priors in our calibration algorithm.Calibration priors are identified in the parameters table and illustrated in detail in Section 12.1.

| | Value | Units | Evidence |
|---|---|---|---|
| Effective contact rate | Calibrated | Contacts per person per day | Calibrated within plausible range |

| | | | |
|---|---|---|---|
| Latent period | Calibrated | Days | The shorter serial interval for Omicron compared to previous variants would also be consistent with a shorter latent period [19, 9, 21]. Transmission pairs were identified through contact tracing from an outbreak of BA.1.1 in Hangzhou (China) in early 2022, through which the latent period was estimated at 3.1 days $95\% CI: 2.8 - 3.5$ [52]. The analysis used censoring to account for uncertainty in the time of infection and infectiousness onset from their data on time of contact with the assumed index case and the time of first positive PCR result. |
| Infectious period | Calibrated | Days | Multiple studies have found a shorter serial interval or generation time for Omicron compared to previous variants [19, 9, 21, 52], with values of five to seven days or less implying short latent and infectious periods. Taking duration of culturable virus as the best proxy for the infectious period, a systematic review and meta-analysis focused on the Omicron variant and reported the time from symptom onset or first positive PCR result to the day after the last positive viral culture to be 5.16 days $95\% CI: 4.18 - 6.14$ [51]. We take this as an upper limit of the effective infectious duration because this quantity may be modified as symptomatic persons modify their behaviour to reduce contacts whilst infectious [52]. |

| | | | |
|---|---|---|---|
| Natural immunity period | Calibrated | Days | Early reinfection with SARS-CoV-2 is known to occur,[31] and antibodies against the virus decline significantly in the months following infection.[18, 6] However, estimating the duration of natural immunity following infection with SARS-CoV-2 is challenging because reinfection due to the development of immune-escape properties in circulating viruses occurs contemporaneously with waning of immunity. An analysis of non-SARS-CoV-2 human coronaviruses that fitted a transmission model to positive laboratory test results estimated that immunity against infection waned over 45 weeks.[25] Another analysis of non-SARS-CoV-2 human coronaviruses used antibody dynamics to conclude that reinfection typically occurred 12 months or more following past infection.[10] However, an analysis of antibody dynamics for a range of human-infecting coronaviruses found that the duration of protection following SARS-CoV-2 infection was less than half of that of several other human-infecting coronaviruses, with reinfections likely occurring from as early as three months post-infection.[43] |
| Case detection rate | Calibrated | | Calibrated across an uniform/uninformative but plausible range. |
| Proportion with increased immunity | 0.4 | | Allowed to vary through full range of possible values (zero to one) based on observation in model development that the sharpness of the BA.1 peak was more difficult to capture in the absence of heterogeneity in susceptibility. |

| | | | |
|---|---|---|---|
| Post-vaccination period partial immunity | Calibrated | Days | Neutralising antibody titres have been identified as key correlates of vaccine-derived immunological protection against infection with SARS-CoV-2.[14, 13, 35] However, neutralising antibodies and vaccine efficacy against infection with the wild-type virus were found to wane significantly over the first 250 days following vaccination.[24] More recently, whilst an initial booster with a third monovalent wild-type mRNA vaccine dose produced a considerable increase in neutralising antibody titres against Omicron, the neutralising antibody response waned significantly by 3-4 months.[15, 49] Observational studies have similarly demonstrated that, following a third monovalent booster, vaccine effectiveness against infection with Omicron waned significantly over 3-4 months.[33] A prospective cohort study of healthcare workers in Israel demonstrated similar neutralising antibody titre and waning profiles following third and fourth doses.[3] Similar to a third booster dose, observational data have suggested that a fourth dose provides about 3-4 months of protection against infection before waning.[2] To accommodate these lines of evidence, we chose a prior distribution that broadly spanned conditions under which the bulk of vaccine-derived immunity against infection was lost within 3-4 months (also noting that the modelled vaccination program is lagged by 14 days to allow for the development of immunity). |
| Immunity infection protection | Calibrated | | Allowed to vary through full range of possible values (zero to one) based on observation in model development that the sharpness of the BA.1 peak was more difficult to capture in the absence of heterogeneity in susceptibility. |
| Adjuster applied to all age-specific IFRs | Calibrated | | Adjustment applied to all age-specific base IFR estimates, see 10.1. |
| BA.1 seed start time | Calibrated | | Calibrated within range that produces model outputs consistent with national sequencing data. |
| BA.2 seed start time | Calibrated | | Calibrated within range that produces model outputs consistent with national sequencing data. |
| BA.5 seed start time | Calibrated | | Calibrated within range that produces model outputs consistent with national sequencing data. |

| | | |
|---|---|---|
| BA.2 escape BA.1 immunity | Calibrated | BA.2 appears to be a significant immune-escape variant to previous strains of SARS-CoV-2, including past BA.1 infection. A single past BA.1 infection induces markedly lower titres of neutralising antibodies against BA.2 than does a past BA.2 infection.[37] A test-negative case-control study of health care workers in Quebec found a 72% (95% CI 65-78) reduction in risk of SARS-CoV-2 infection for those previously infected with BA.1 in unvaccinated participants during a period of predominant BA.2 transmission, [4] although indication for testing differed markedly between cases and controls. Another test-negative case-control study conducted in adolescents in the UK during the BA.1 and BA.2 periods found that previous Omicron infection conferred 59.3% (95% CI 46.7-69.0) protection against Omicron reinfection in the unvaccinated.[34] Although this figure does not distinguish between sub-variants, this analysis is likely to include many BA.2 reinfections following previous BA.1 infection. A third estimate from the highly vaccinated population of Qatar found very high levels of protection 94.2% (95% CI 89.2-96.9) against BA.2 reinfection from past BA.1 infection, although follow-up was relatively short.[5] For all of these studies, distinguishing between waning with immunity with time and escape properties of the virus are extremely hard to disentangle. We note that in addition to Australia several other settings have observed BA.2 waves that have followed closely on the heels of a declining BA.1 epidemic, including the UK, France and the Netherlands.[11] This could suggest significant immune escape properties of BA.2 against BA.1 immunity or a markedly greater reproduction number. Given the extremely infectious nature of all Omicron strains, the former seems more epidemiologically plausible. |

| | | |
|---|---|---|
| BA.5 escape BA.1 or BA.2 immunity | Calibrated | BA.5 again shows significant immune-escape to previous SARS-CoV-2 variants, including BA.1 and BA.2, with lower levels of neutralising antibodies generated against BA.5 from past BA.1 infection than for BA.2 infection.[37] The loss of neutralisation activity against BA.5 following a BA.1 infection appears greater in unvaccinated than vaccinated persons.[23] Significant immune evasion of BA.5 against prior Omicron subvariant infection is suggested by the UK's ONS infection survey, which found that 54% of BA.5 reinfections had their first infection during the BA.1 or BA.2 waves.[41] Observational studies have estimated the real-world protection against BA.5 reinfection conferred by prior BA.1 or BA.2 infection. Portugal was one of the first countries to be affected by the BA.5 subvariant, only a few months after a major mixed BA.1 and BA.2 epidemic. Infection with BA.1 or BA.2 was estimated to confer 75.3% protection against BA.5 infection, which was somewhat greater than for previous variants (e.g. Delta 61.3% protection).[27] The authors note a perception of very low protection of past BA.1/BA.2 infection against BA.5, but attribute this to moderate protection in a setting with a high attack rate from the recent BA.1/BA.2 wave. A national test-negative case-control study in Qatar similarly found that prior infection with BA.1 or BA.2 was 68.7% effective in preventing reinfection in the unvaccinated.[1] A retrospective cohort study from Singapore found no protection against BA.4/5 infection was conferred from prior BA.1/2 infection, although vaccination status differed markedly from the previously infected and infection-naive groups.[42] Although not directly applicable to this parameter, these findings are consistent with substantial immune escape. A limitation of these three studies [27, 1, 42] is that they leverage passive surveillance data and so may underestimate the true number of infections, particularly asymptomatic or pauci-symptomatic infections and hence underestimate the extent of immune escape. Prospective household contact transmission studies with an active surveillance strategy may better estimate the effect of prior infection,[29] but were unavailable for the variants considered here. |

| | | |
|---|---|---|
| Relative IFR for BA.2 compared to BA.1 and BA.5 | Calibrated | Two national-level studies estimated the severity of BA.2 relative to BA.1 using the time period of the epidemic combined with PCR S-gene target failure status to identify cases of each sub-variant. In the United Kingdom, the hazard ratio for death was estimated at 0.80 (95%CI 0.71-0.90) for BA.2 compared to BA.1.[47] Although a large South African study did not estimate the relative risk of death, the adjusted odds ratio for a composite severe disease outcome was comparable to this estimate (0.78, 95%CI 0.63-0.97) [50]. A nationwide study from Denmark [17] leveraged high coverage with PCR testing and whole-genome sequencing to estimate the odds of hospitalisation with BA.5 relative to BA.2, estimating an adjusted odds ratio of 1.69 (95%CI 1.22-2.33). An analysis from Scotland was consistent with a comparably increased adjusted hazard of hospitalisation (1.21, 95%CI 1.03, 1.43) and death (1.55, 95%CI 0.78-3.08), although with a broad uncertainty ranges.[36] Although these estimates comparing BA.5 to BA.2 may suggest a greater difference in severity than between BA.1 and BA.2, the South African analysis found no difference in severity between BA.1 and BA.5.[50] |

| | | | |
|---|---|---|---|
| Time to WA fully mixing with rest of country | Calibrated | Days | Although the Western Australia (WA) Government re-opened borders from 3rd March 2022, some restrictions initially remained in place after this date, including the requirement for a rapid antigen test on arrival into the state.[48] Further, domestic air travel into the WA remained below pre-Covid levels until around mid-year 2022.[20] More importantly, the modelled assumption of homogeneous mixing within population age groups means that it would be unrealistic that the population of WA would be subject to the same force of infection as other Australian jurisdictions immediately from the date of re-opening. For this reason, we scale up the rate of infection in WA to reach the same level as the remainder of the country over a period of time that is varied through calibration.Although the Western Australia (WA) Government re-opened borders from 3rd March 2022, some restrictions initially remained in place after this date, including the requirement for a rapid antigen test on arrival into the state.[48] Further, domestic air travel into the WA remained below pre-Covid levels until around mid-year 2022 and was relatively lower than other Australian states until April-May 2022.[20] More importantly, the modelled assumption of homogeneous mixing within population age groups means that it would be unrealistic to assume that the population of WA would be subject to the same force of infection as other Australian jurisdictions immediately from the date of re-opening. For this reason, we scale up the rate of infection in WA to reach the same level as the remainder of the country over a period of time that is consistent with a mid-year epidemic. |
| New variant seeding duration | 10.0 | Days | Arbitrarily set to a low value to introduce new sub-variant. |
| New variant seeding rate | 1.0 | Persons per day | Arbitrarily set to a low value to introduce new sub-variant. |

| | | | |
|---|---|---|---|
| Gamma distribution mean, infection to notification delay | Calibrated | Days | By contrast to the time from symptom onset to death, which we consider to be more a biological phenomenon intrinsic to the pathogen, we searched for locally applicable estimates to inform the delay from symptom onset to case notification. A published analysis estimating the transmission potential in Australia from February 2020 to February 2021[16] provided publicly accessible data on the number of days from symptom onset to notification in Victoria and New South Wales. From these data we calculated a mean delay from symptom onset to notification of 4.17 days. |
| Gamma distribution shape, infection to notification delay | 2.0 | | Estimated from plotting distribution of notification data.[16] |
| Gamma distribution mean, infection to death delay | Calibrated | Days | A cohort of patients presenting to two central Wuhan hospitals early in the pandemic estimated a mode of 14 days for time from symptom onset to death,[38] while a study of case presentations outside of the Wuhan epicentre estimated a mean of 20.2 days (95% CI 7.4-39.9) when right truncation was accounted for.[26] These two studies were combined with a second Wuhan cohort [39] (likely overlapping with the first Wuhan study[38]) in a meta-analysis[22] that estimated the mean time to death at 15.93 days (95% CI 13.07-18.79). An analysis of Victorian data from 2020 estimated the mean time from diagnosis to death at 18.1 days (95% CI 16.9-19.3), which varied little by age.[28] Although fewer estimates were found for the Omicron period, and despite the clearly lower fatality rate for Omicron than for Delta, a study of adults in the UK from December 2021 found the same mean delay from testing positive to death for Omicron BA.1 as for Delta (18 days).[45] |
| Gamma distribution shape, infection to death delay | 5.0 | | A shape parameter of five appeared to broadly reproduce the distribution of deaths presented in the first Wuhan cohort [38] and the upper centiles reported in the non-Wuhan Chinese presentations analysis.[26] |
| IFR, ages 0 to 4 | 6e-06 | | See Section 10.1 text. |
| IFR, ages 5 to 9 | 2e-06 | | See Section 10.1 text. |

| | | |
|---|---|---|
| IFR, ages 10 to 14 | 2e-06 | See Section 10.1 text. |
| IFR, ages 15 to 19 | 6e-06 | See Section 10.1 text. |
| IFR, ages 20 to 24 | 1.71e-05 | See Section 10.1 text. |
| IFR, ages 25 to 29 | 2.79e-05 | See Section 10.1 text. |
| IFR, ages 30 to 34 | 3.73e-05 | See Section 10.1 text. |
| IFR, ages 35 to 39 | 4.67e-05 | See Section 10.1 text. |
| IFR, ages 40 to 44 | 5.61e-05 | See Section 10.1 text. |
| IFR, ages 45 to 49 | 8.62e-05 | See Section 10.1 text. |
| IFR, ages 50 to 54 | 0.000121 | See Section 10.1 text. |
| IFR, ages 55 to 59 | 0.00016 | See Section 10.1 text. |
| IFR, ages 60 to 64 | 0.000205 | See Section 10.1 text. |
| IFR, ages 65 to 69 | 0.000258 | See Section 10.1 text. |
| IFR, ages 70 to 74 | 0.000383 | See Section 10.1 text. |
| IFR, ages 75 and above | 0.0014 | See Section 10.1 text. |

***Table 2:*** **Epidemiological parameters and evidence.**.

#### 10.1 Infection Fatality Rates

Age-specific infection fatality rates (IFRs) have previously been estimated by various groups in unvaccinated populations, including O'Driscoll and colleagues who estimated IFRs using data from 45 countries [32]. These IFRs pertained to the risk of death given infection for the wild-type strain of SARS-CoV-2 in unvaccinated populations, and so are unlikely to represent IFRs that would be applicable to the Australian population in 2022 because of vaccine-induced immunity and differences in severity in severity between the wild-type variant and Omicron subvariants simulated in this analysis. We therefore considered more recent studies, such as that of Erikstrup and colleagues to be better applicable to our local context, although also with limitations. Danish investigators used the increase in anti-nucleocapsid IgG seroprevalence in blood donors from January to April 2022 to estimate age-specific attack rates for the first Omicron wave in Denmark. [12] They then re-weighted these values to estimate the attack rate for the general population aged 17-72. Linking this estimate to COVID-19 deaths reported within 60 days of a positive PCR, they estimated the Omicron-specific IFR, which was then re-weighted to exclude people with comorbidities. Therefore, their final results used in our analysis represent an Omicron-specific IFR for a healthy vaccinated

***Figure 16:*** **Illustration of the calculation of the base age-specific infection-fatality rates applied in the model.** O'Driscoll and Erikstrup indicate the original data reported in the studies of interest. Subsequent traces indicate the further steps in estimating values for use in the model, with the last trace representing the parameter applied according to the lower value of each age bracket.

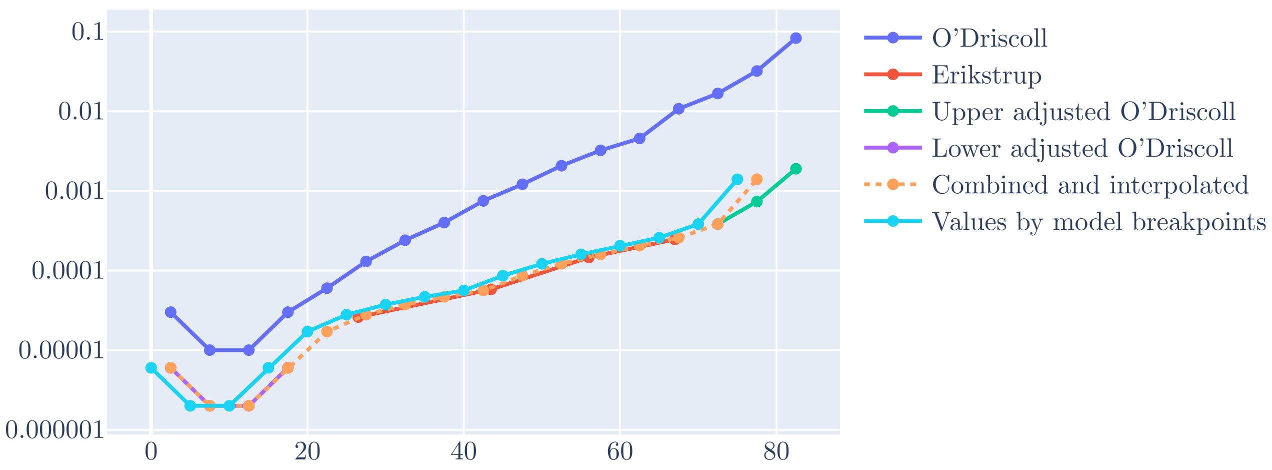

population aged 17 to 72 years.As expected, the estimates from Erikstrup are consideraly lower than those of O'Driscoll. However, there are also several potential differences between the Danish epidemic and that of Australia, most notably that community transmission had been established from much earlier in the pandemic in Denmark than in Australia, such that the Danish population would have markedly greater natural immunity, which could provide significant additional protection given the same vaccination status. Further, given that these estimates estimate attack rates from blood donors, the age ranges covered by this study extend from 17 years to 73 years of age, making it necessary to extrapolate from these estimates to the extremes of age. We approached this extrapolation by identifying broadly equivalent younger and older age groups from each study for use as baselines for the more extreme age groups. Specifically, we considered that the IFR estimate for the 17 to 36 years-old age group from Erikstrup could be compared to the 25 to 29 years-old age group form O'Driscoll, and that the 61 to 73 years-old age group from Erikstrup could be compared to the 65 to 69 years-old age group from O'Driscoll. We next calculated the ratio in the IFRs of these 'equivalent' age groups from each study, before then applying these ratios to the estimates from O'Driscoll for the age bands outside of the age range calculated by Erikstrup. (i.e. 0-4, 5-9, 10-14, 15-19, 70-74, 75-79 and 80+). Next, to obtain IFR estimates for each modelled 5-year band from 75-79 years-old we performed linear interpolation from the estimates available to the mid-point of each modelled age band. We now have estimates for each 5-year band from 0-4 to 75-79 and for 80+ years-old. To calculate the IFR parameter for the modelled 75+ age band, we took an average of the 75-79 and 80+ estimates, weighted using the proportion of the Australian population aged 75+ who are aged 75-79 and 80+.

### 11 Targets

Calibration targets were constructed as described throughout the following subsections and summarised in Figure 17.

#### 11.1 Notifications

Because official Australian Government notification data were unavailable for 2021, and the initial upslope of the epidemic occurred in the last months of this year, the calibration target for cases was constructed from the 'Our World in Data' (OWID) data for 2021 concatenated with the Australian Government data for 2022. That is, we preferentially used Australian Government data throughout most of our simulation period of interest, which is extracted from Australia's national surveillance reporting system. Note that daily case data for Australia were unavailable from the World Health Organization website (which draws from the Australian NNDSS), because reporting changed to weekly in 2021. Official COVID-19 data for Australian notifications through 2022 were obtained from The Department of Health on the 2$^{nd}$ of May 2023. Data that extended back to 2021 were obtained from OWID on the 16$^{th}$ of June 2023. The composite daily case data were then smoothed using a 7-day moving average. The notifications value for each date of the analysis were compared against the modelled estimate from a given parameter set using a negative binomial distribution. The dispersion parameter of this negative binomial distribution was calibrated from an uninformative prior distribution along with the epidemiological parameters through the calibration algorithm. The effect of the dispersion parameter on the comparison between modelled and empiric values is illustrated in Figure 18.

***Figure 17:*** **Calibration targets with raw data from which they were derived.**

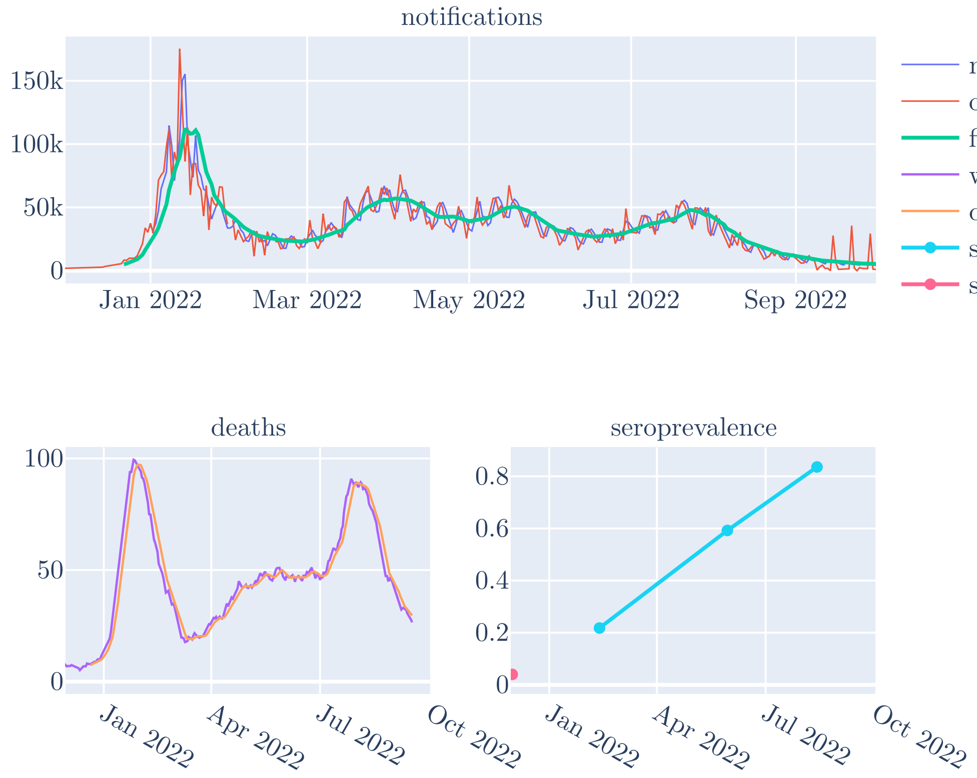

***Figure 18:*** **Dispersion examples.** Examples of the effect of values of the negative binomial distribution dispersion parameter, centiles of likelihood distribution. Actual targets used for likelihood calculation circles.

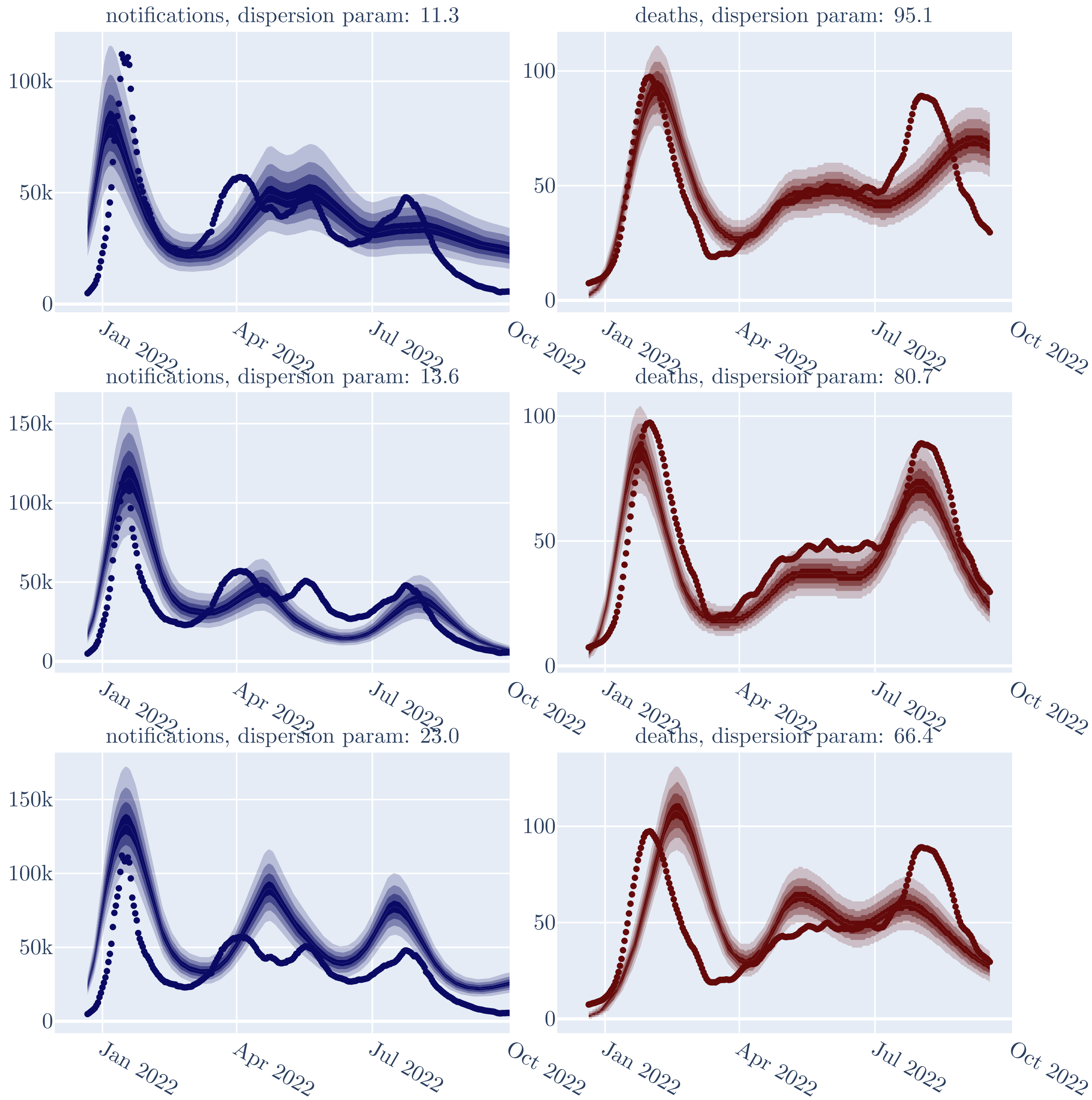

#### 11.2 Deaths

The daily time series of deaths for Australia was obtained from the World Health Organization's Coronavirus (COVID-19) Dashboard, downloaded on 18$^{th}$ of July 2023. These data were also smoothed using a 7-day moving average. As for case notifications, the comparison distribution used to obtain the likelihood of a given parameter set was negative binomial with calibrated dispersion parameter.

#### 11.3 Seroprevalence

In Australia, all programmatically available vaccines were directed at the SARS-CoV-2 spike protein, such that nucleocapsid-directed antibodies serve to indicate past immunological exposore to the virus. We obtained estimates of the seroprevalence of antibodies to nucleocapsid antigen from Australian blood donors from Kirby Institute serosurveillance reports. Data are available from the round 4 serosurvey, with information on assay sensitivity also reported. The raw values reported in the serosurvey were inflated by the reported assay sensitivity (i.e. 0.78), with no adjustment made for the (assumed very high) specificity of the assay. We lagged these empiric estimates by 14 days to account for the delay between infection and seroconversion. Although anti-nucleocapsid antibodies likely wane with time to some extent, most exposed persons maintain a response for ten months following infection, which is well within the time period of infection for which we used these estimates [44]. The proportion of the population seropositive was compared against the modelled proportion of the population ever infected using a binomial distribution.

We added a further recovered proportion target to avoid accepting runs with higher likelihood values in which the acceptable fit to data was a result of an implausibly high initial epidemic wave that occurred prior to the availability of target data (i.e. in late 2021 during the model run-in period). We term this the 'seroprevalence ceiling'. This was achieved by adding a large negative number to the likelihood estimate for any runs with a proportion ever infected greater than 4% on 1$^{st}$ of December 2021.

#### 11.4 Calibrated dispersion parameters

Figure 18 provides an illustration of the effect of specific values of the calibrated dispersion parameter used in the calibration algorithm to adjust the calculation of the contribution to the likelihood from the notifications and deaths time series.

### 12 Calibration methods

We calibrated our epidemiological model using the 'DE Metropolis(Z)' algorithm provided in the PyMC package for Bayesian inference.

Prior to running the algorithm, we ran a short optimisation using 2-point differential evolution provided by Facebook Research's nevergrad library. Initialisation of the optimisation algorithm was performed using Latin hypercube sampling to select initial parameter values that were broadly dispersed across the multi-dimensional parameter space. The deliberately short optimisation algorithm of 100 draws was used to move the parameter sets from these dispersed starting positions towards values with greater likelihood, but remained substantially dispersed from one-another.

We then ran the calibration algorithm from these starting points for 10,000 draws during the tuning phase, and for 50,000 draws during the calibration phase of the algorithm, discarding the first 25,000 draws as burn-in. An identical and independent algorithm was applied for each of the four analysis approaches.

##### 12.1 Priors

The priors used in any of the four analysis presented are described in this section, and displayed in Figure 19 and Table 3. In the case of the two alternative analyses incorporating time-varying (vaccine-induced) immunity, the 'Prop immune' parameter is not included in the priors implemented; whereas in the case of the two analyses not involving time-varying immunity, the 'Vacc immune period' parameter is omitted.

The priors used in any of the four analysis presented are described in this section, and displayed in Figure 19 and Table 3. In the case of the two alternative analyses incorporating time-varying (vaccine-induced) immunity, the 'Prop immune' parameter is not included in the priors implemented; whereas in the case of the two analyses not involving time-varying immunity, the 'Vacc immune period' parameter is omitted. The priors used in any of the four analysis presented are described in this section, and displayed in Figure 19 and Table 3. In the case of the two alternative analyses incorporating time-varying (vaccine-induced) immunity, the 'Prop immune' parameter is not included in the priors implemented; whereas in the case of the two analyses not involving time-varying immunity, the 'Vacc immune period' parameter is omitted. The priors used in any of the four analysis presented are described in this section, and displayed in Figure 19 and Table 3. In the case of the two alternative analyses incorporating time-varying (vaccine-induced) immunity, the 'Prop immune' parameter is not included in the priors implemented; whereas in the case of the two analyses not involving time-varying immunity, the 'Vacc immune period' parameter is omitted. The priors used in any of the four analysis presented are described in this section, and displayed in Figure 19 and Table 3. In the case of the two alternative analyses incorporating time-varying (vaccine-induced) immunity, the 'Prop immune' parameter is not included in the priors implemented; whereas in the case of the two analyses not involving time-varying immunity, the 'Vacc immune period' parameter is omitted.

#### 13 Analysis comparison

We compared our four candidate analyses according to their goodness of fit to the targets data (described under Section 11). The fit of all four of the models to the target data was considered adequate, but the likelihood of the 'mob' analysis was slightly higher than that of the other three approaches, with the inclusion of the mobility structure appearing to improve the calibration algorithm's fit to targets (See Figure 23). For this reason, the 'mob' analysis was considered as the primary analysis throughout the remaining sections. Figures 20, 21 and 22 illustrate the fit of each candidate model to the target data for the notification, death and seropositive proportion respectively.

#### 14 Calibration results

Results for the credible intervals around the main outputs compared against their targets are presented in Figure 26.

| | Distribution | Parameters | Support |
|---|---|---|---|
| Effective contact rate | Uniform distribution | loc: 0.02 scale: 0.13 | 0.02 to 0.15 |
| Latent period | Gamma distribution | shape: 56.645 scale: 0.045 | 0.0 to infinity |
| Infectious period | Gamma distribution | shape: 50.344 scale: 0.071 | 0.0 to infinity |
| Natural immunity period | Gamma distribution | shape: 2.511 scale: 119.114 | 0.0 to infinity |
| Case detection rate | Uniform distribution | loc: 0.1 scale: 0.5 | 0.1 to 0.6 |
| Immunity infection protection | Uniform distribution | loc: 0.0 scale: 1.0 | 0.0 to 1.0 |
| Adjuster applied to all age-specific IFRs | TruncNormal distribution | loc: 1.0 scale: 2.0 a: -0.4 b: inf | 0.2 to infinity |
| BA.1 seed start time | Uniform distribution | loc: 580.0 scale: 45.0 | 580.0 to 625.0 |
| BA.2 seed start time | Uniform distribution | loc: 625.0 scale: 35.0 | 625.0 to 660.0 |
| BA.5 seed start time | Uniform distribution | loc: 660.0 scale: 80.0 | 660.0 to 740.0 |
| BA.2 escape BA.1 immunity | Beta distribution | a: 8.687 b: 13.031 | 0.0 to 1.0 |
| BA.5 escape BA.1 or BA.2 immunity | Beta distribution | a: 8.687 b: 13.031 | 0.0 to 1.0 |
| Relative IFR for BA.2 compared to BA.1 and BA.5 | TruncNormal distribution | loc: 0.7 scale: 0.15 a: -3.333 b: inf | 0.2 to infinity |
| Time to WA fully mixing with rest of country | Uniform distribution | loc: 30.0 scale: 45.0 | 30.0 to 75.0 |
| Gamma distribution mean, infection to notification delay | Gamma distribution | shape: 19.51 scale: 0.214 | 0.0 to infinity |
| Gamma distribution mean, infection to death delay | Gamma distribution | shape: 226.255 scale: 0.07 | 0.0 to infinity |
| Post-vaccination period partial immunity | Gamma distribution | shape: 5.416 scale: 13.588 | 0.0 to infinity |
| Proportion with increased immunity | Uniform distribution | loc: 0.0 scale: 1.0 | 0.0 to 1.0 |

***Table 3:*** **Priors.**. Parameters implemented in calibration code are given. Note that the values for several of these were generated through algorithms that aimed to approximate epidemiological user-specified plausible ranges.

***Figure 19:*** **Priors.** Illustrations of prior distributions implemented in calibration algorithm.

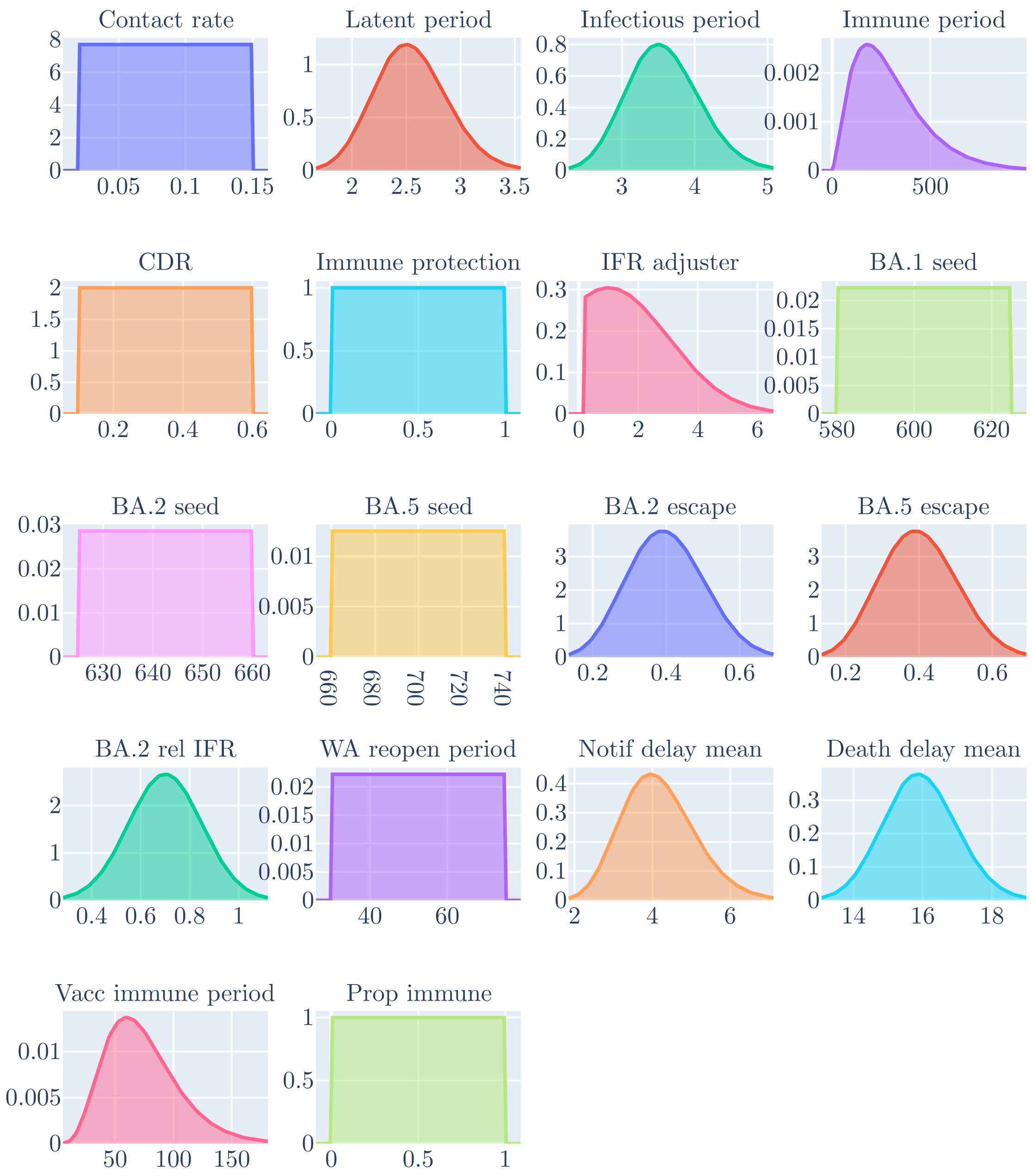

***Figure 20:*** **Cases credible intervals by analysis.** Case notifications with median estimate (black line), 2.5 to 97.5 centile credible interval (light blue shading), and 25 to 75 centile credible interval (dark blue shading), with comparison against epidemiological targets (red circles). Panel for each of the four candidate analyses.

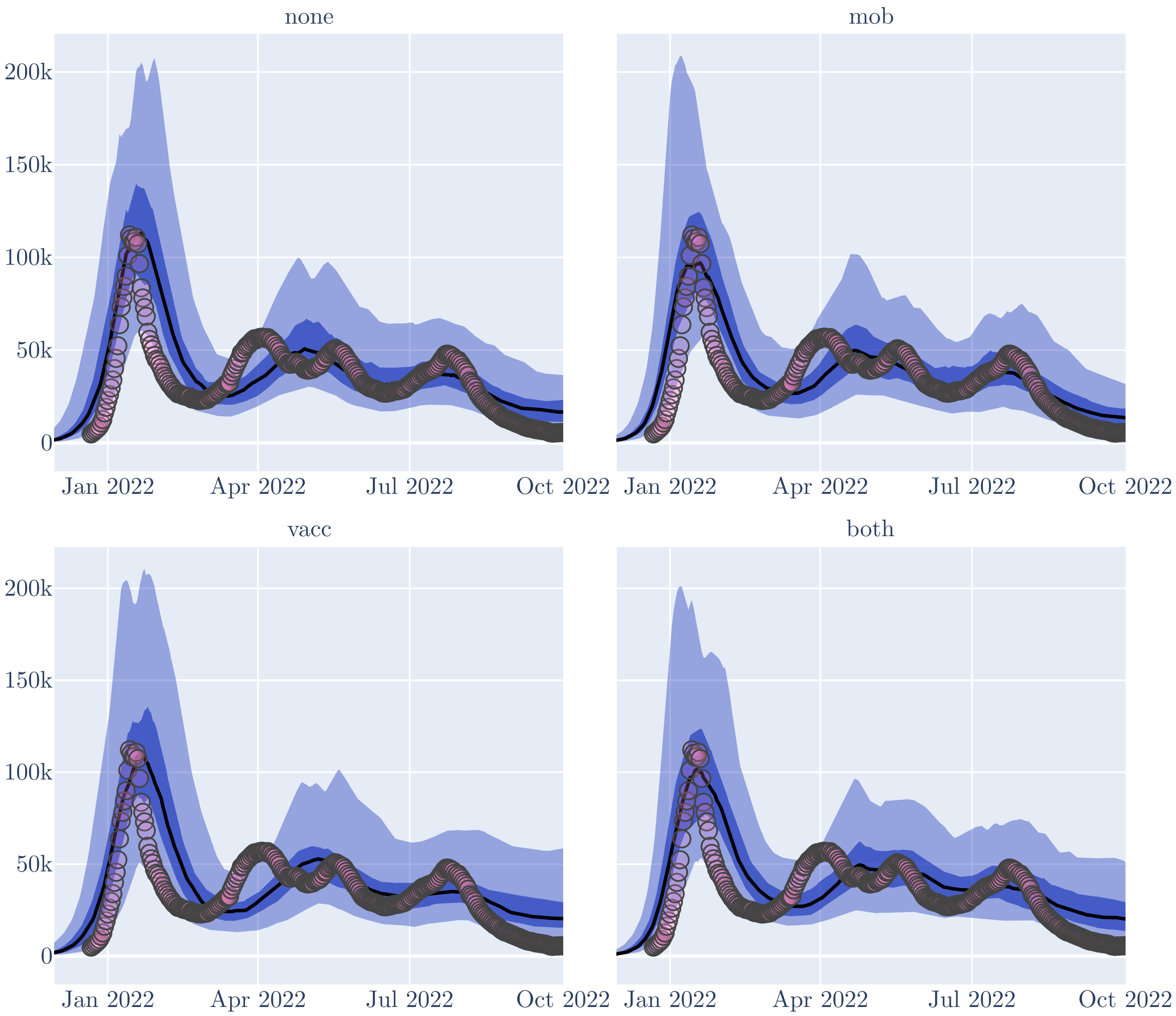

***Figure 21:*** **Deaths credible intervals by analysis.** Deaths with median estimate (black line), 2.5 to 97.5 centile credible interval (light blue shading), and 25 to 75 centile credible interval (dark blue shading), with comparison against epidemiological targets (red circles). Panel for each of the four candidate analyses.

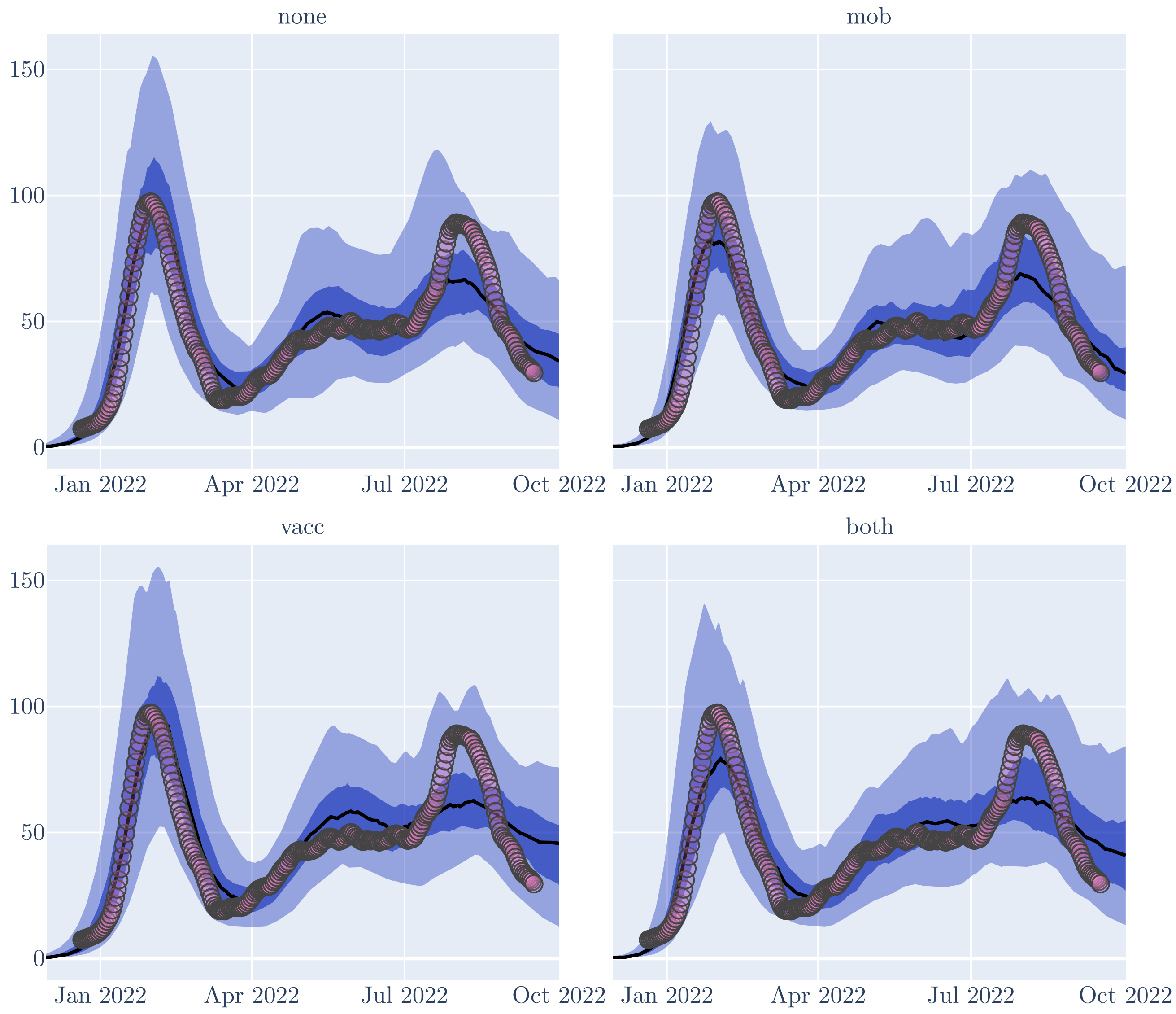

***Figure 22:*** **Seropositive proportion credible intervals by analysis.** Seropositive proportion with median estimate (black line), 2.5 to 97.5 centile credible interval (light blue shading), and 25 to 75 centile credible interval (dark blue shading), with comparison against epidemiological targets (red circles). Panel for each of the four candidate analyses.

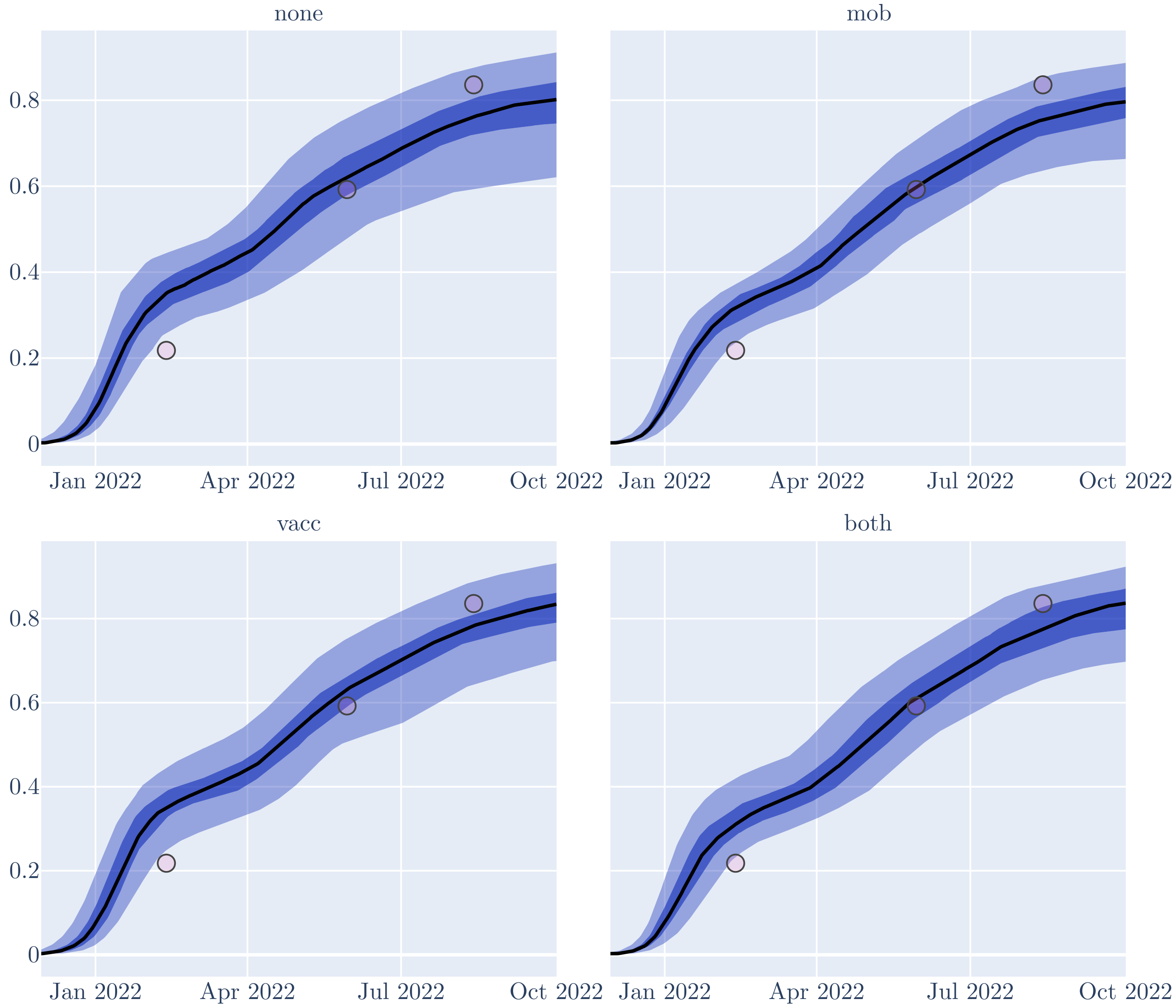

***Figure 23:*** **Likelihood comparison, kernel densities.** Comparison of the kernel density distribution of the final likelihood from calibration algorithm, with the contributions to the final likelihood of the three targets from which it was constructed.

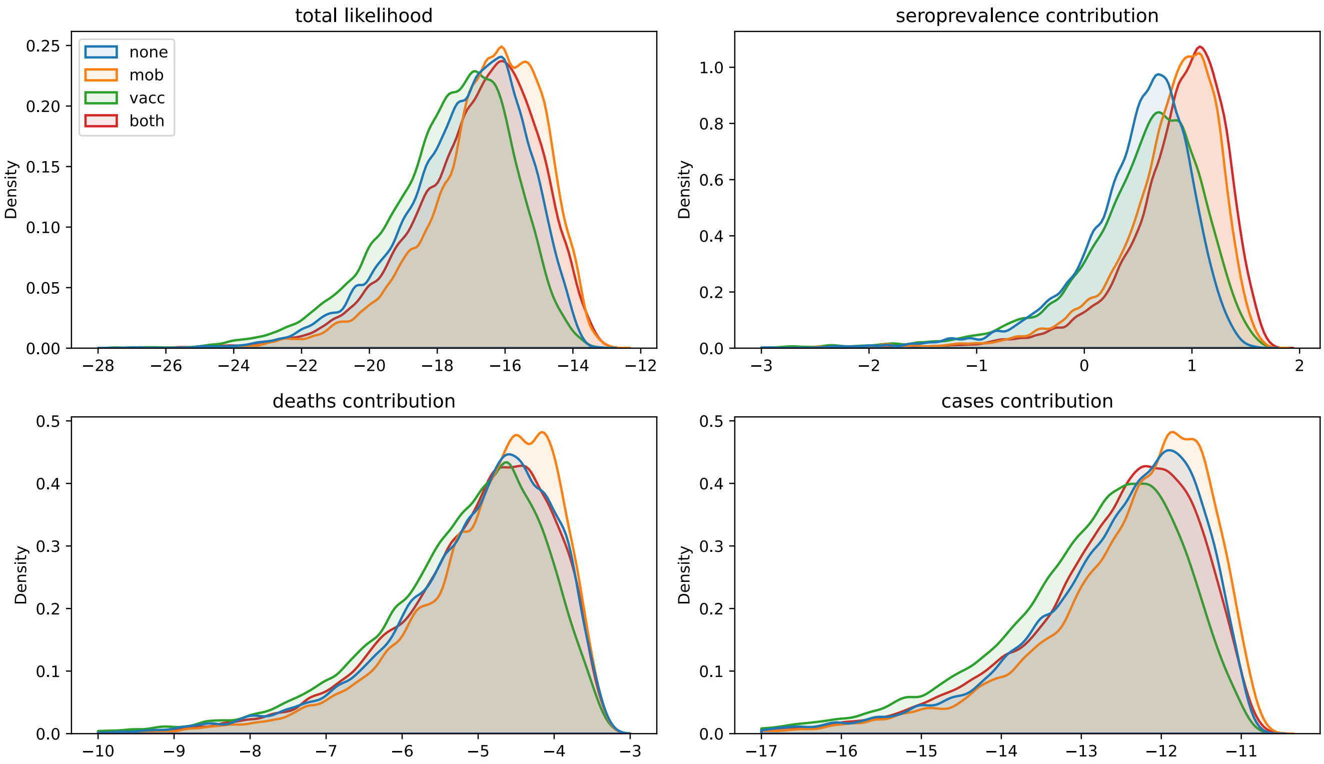

***Figure 24:*** **Key outputs for randomly sampled runs from calibration algorithm.**

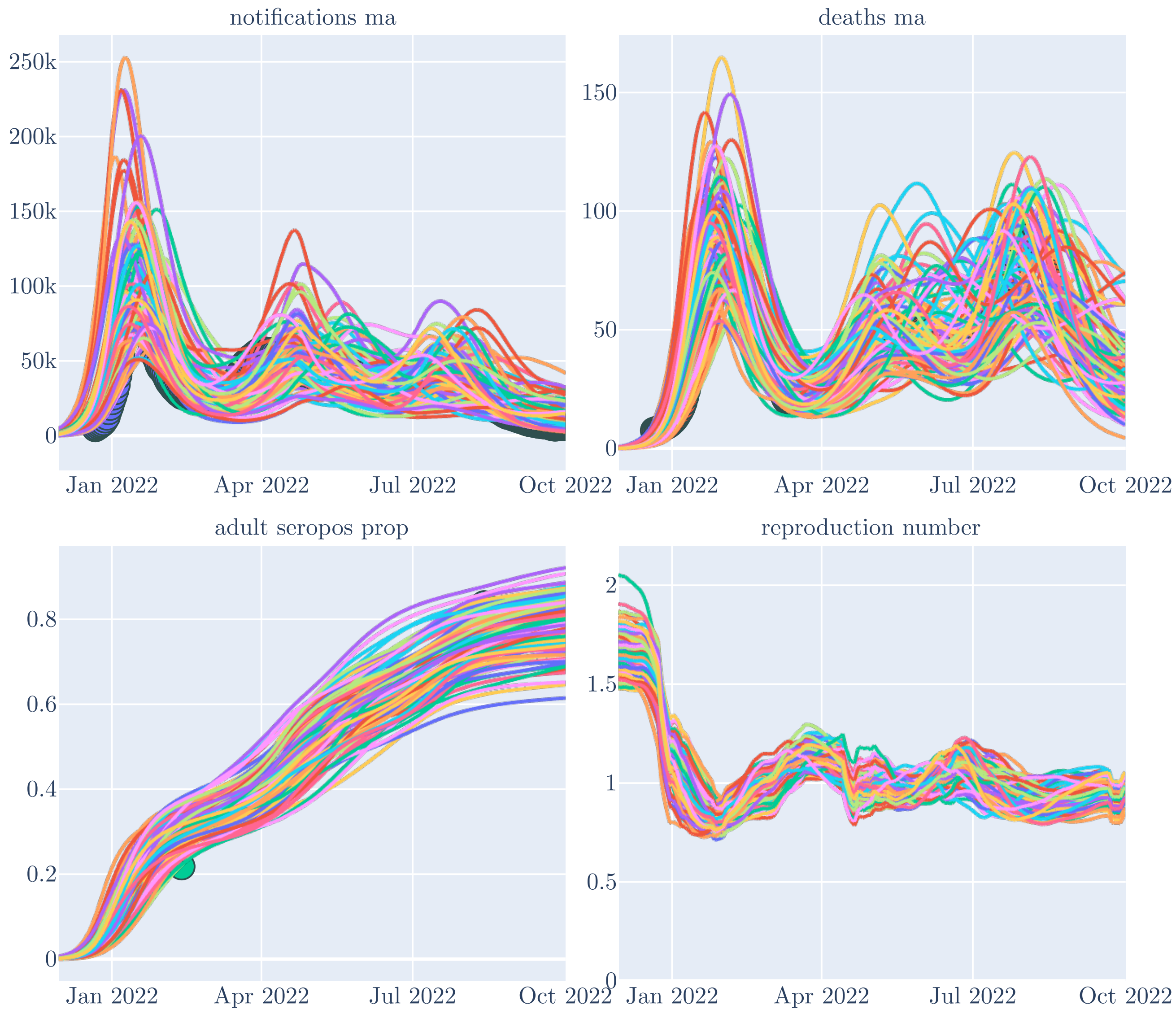

***Figure 25:*** **Proportional prevalence of modelled sub-variants.** Proportion of modelled cases attributable to each sub-variant over time. Solid curved lines, proportion of prevalence attributable to BA.1, Dashed curved lines, proportion of prevalence attributable to BA.1 or BA.2. Key dates for each variant are shown as vertical bars: blue, BA.1; red, BA.2; green, BA.5; dotted, first detection; dashed, 1%; solid, 50%.

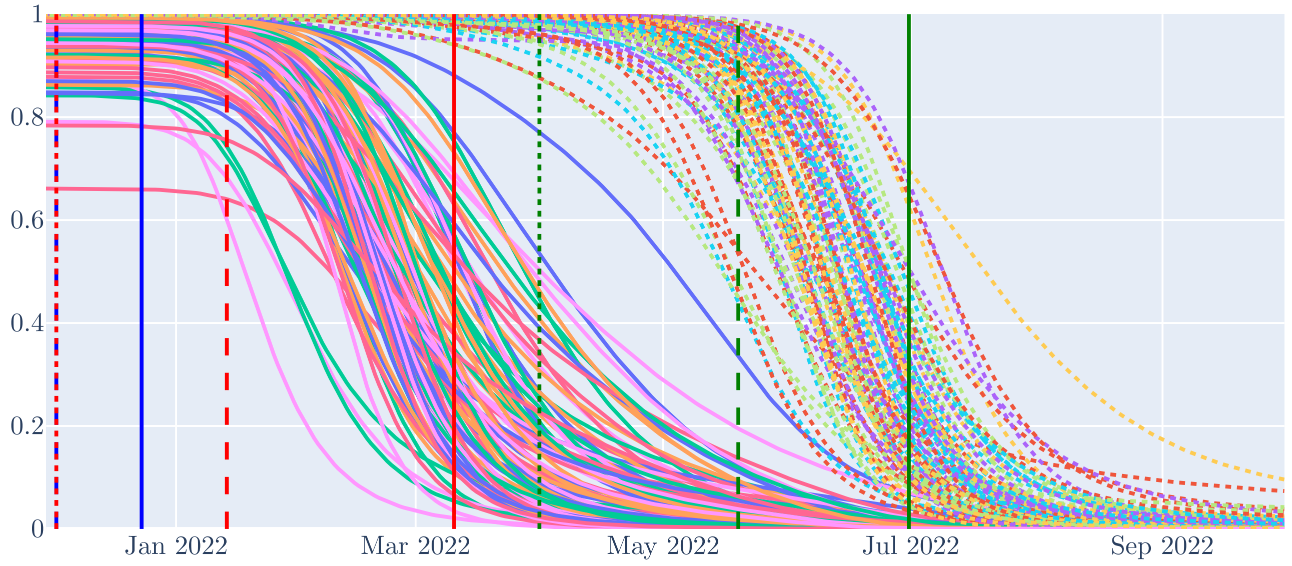

***Figure 26:*** **Primary analysis output credible intervals.** Primary analysis model median estimate (black line), 2.5 to 97.5 centile credible interval (light blue shading), and 25 to 75 centile credible interval (dark blue shading), with comparison against epidemiological targets (red circles). Panel for each epidemiological output.

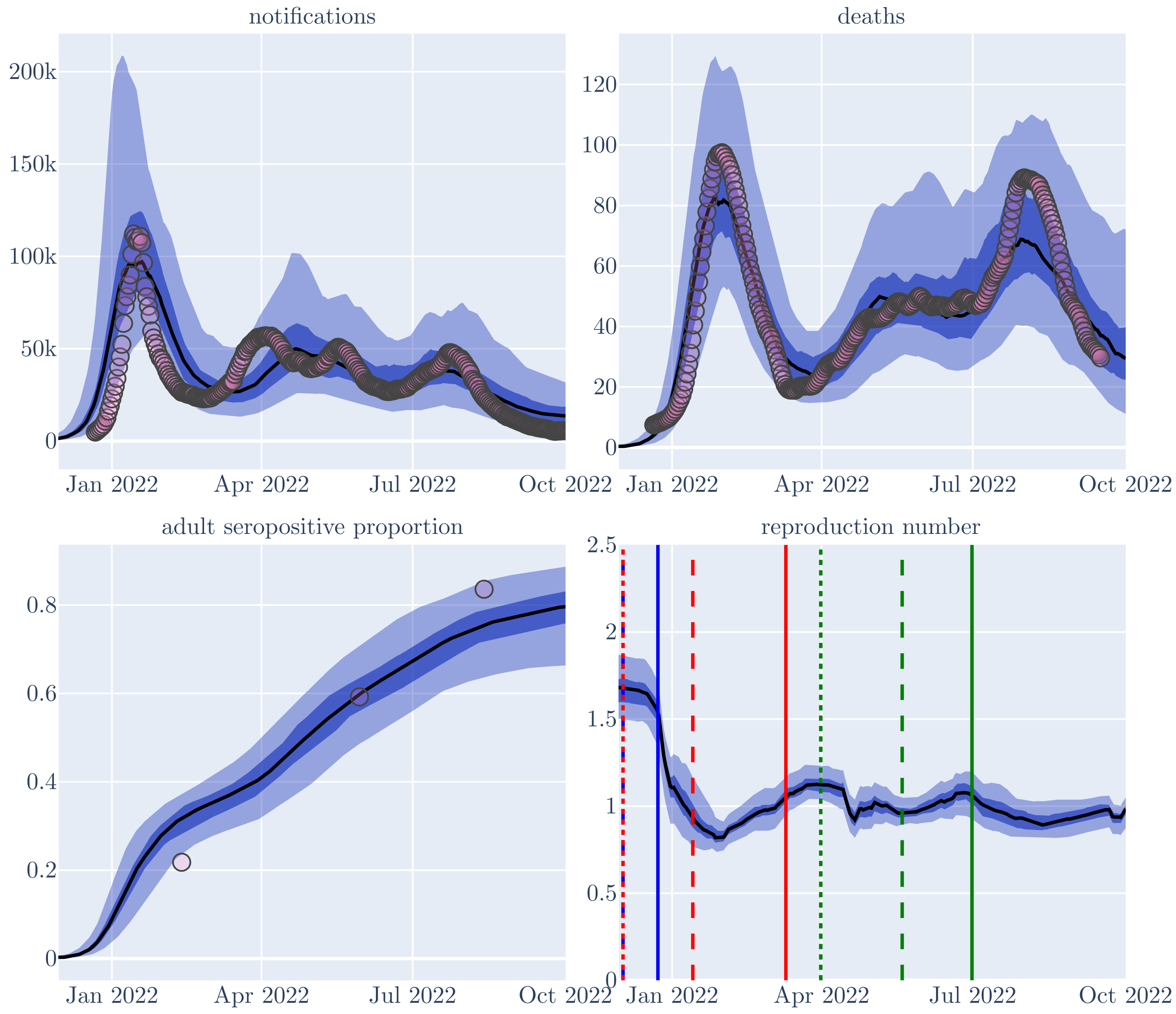

#### 14.1 Calibration performance

| | Mean | Standard deviation | ESS bulk | ESS tail | $\hat{R}$ | High-density interval |
|---|---|---|---|---|---|---|
| Effective contact rate | 0.069 | 0.017 | 281.0 | 470.0 | 1.03 | 0.043 to 0.102 |
| Latent period | 2.58 | 0.32 | 1240.0 | 2260.0 | 1.0 | 1.98 to 3.18 |
| Infectious period | 3.6 | 0.493 | 1190.0 | 2200.0 | 1.01 | 2.71 to 4.52 |
| Natural immunity period | 125.0 | 75.1 | 885.0 | 1390.0 | 1.01 | 30.1 to 257.0 |
| Case detection rate | 0.336 | 0.094 | 902.0 | 913.0 | 1.01 | 0.172 to 0.518 |
| Immunity infection protection | 0.441 | 0.228 | 338.0 | 369.0 | 1.03 | 0.0 to 0.811 |
| Adjuster applied to all age-specific IFRs | 2.39 | 0.566 | 1220.0 | 1920.0 | 1.0 | 1.39 to 3.46 |
| BA.1 seed start time | 610.0 | 10.4 | 916.0 | 983.0 | 1.01 | 591.0 to 625.0 |
| BA.2 seed start time | 642.0 | 9.31 | 957.0 | 963.0 | 1.02 | 625.0 to 657.0 |
| BA.5 seed start time | 694.0 | 17.8 | 817.0 | 1110.0 | 1.01 | 661.0 to 725.0 |
| BA.2 escape BA.1 immunity | 0.448 | 0.085 | 1110.0 | 1650.0 | 1.01 | 0.284 to 0.603 |
| BA.5 escape BA.1 or BA.2 immunity | 0.489 | 0.076 | 972.0 | 2000.0 | 1.01 | 0.341 to 0.628 |

| | | | | | | |
|---|---|---|---|---|---|---|
| Relative IFR for BA.2 compared to BA.1 and BA.5 | 0.703 | 0.139 | 1040.0 | 811.0 | 1.01 | 0.455 to 0.981 |
| Time to WA fully mixing with rest of country | 51.6 | 13.0 | 1160.0 | 1830.0 | 1.01 | 30.3 to 72.1 |
| Gamma distribution mean, infection to notification delay | 4.19 | 0.912 | 1240.0 | 2510.0 | 1.0 | 2.62 to 5.94 |
| Gamma distribution mean, infection to death delay | 16.0 | 1.05 | 1320.0 | 2470.0 | 1.0 | 14.2 to 18.1 |
| Proportion with increased immunity | 0.509 | 0.275 | 179.0 | 576.0 | 1.04 | 0.081 to 0.993 |

***Table 4:*** **Calibration metrics**.

The metrics of the performance of our calibration algorithm are presented in Table 4.

#### 14.2 Parameter inference

Parameter-specific chain traces with parameter and chain-specific posterior densities for the primary 'mob' analysis are presented in Figures 27, 28 and 29. These are used for the epidemiological interpretation of our results in the main manuscript. Overall posterior densitites (pooled over calibration chains) compared against prior distributions are presented in Figures 36 and 37.

#### 14.3 Parameter correlation

Figures 38, 39 and 40 show the bivariate distributions of various sets of pairs of parameters used in the calibration algorithm.

***Figure 27:*** **Parameter posteriors and traces by chain, 1.**

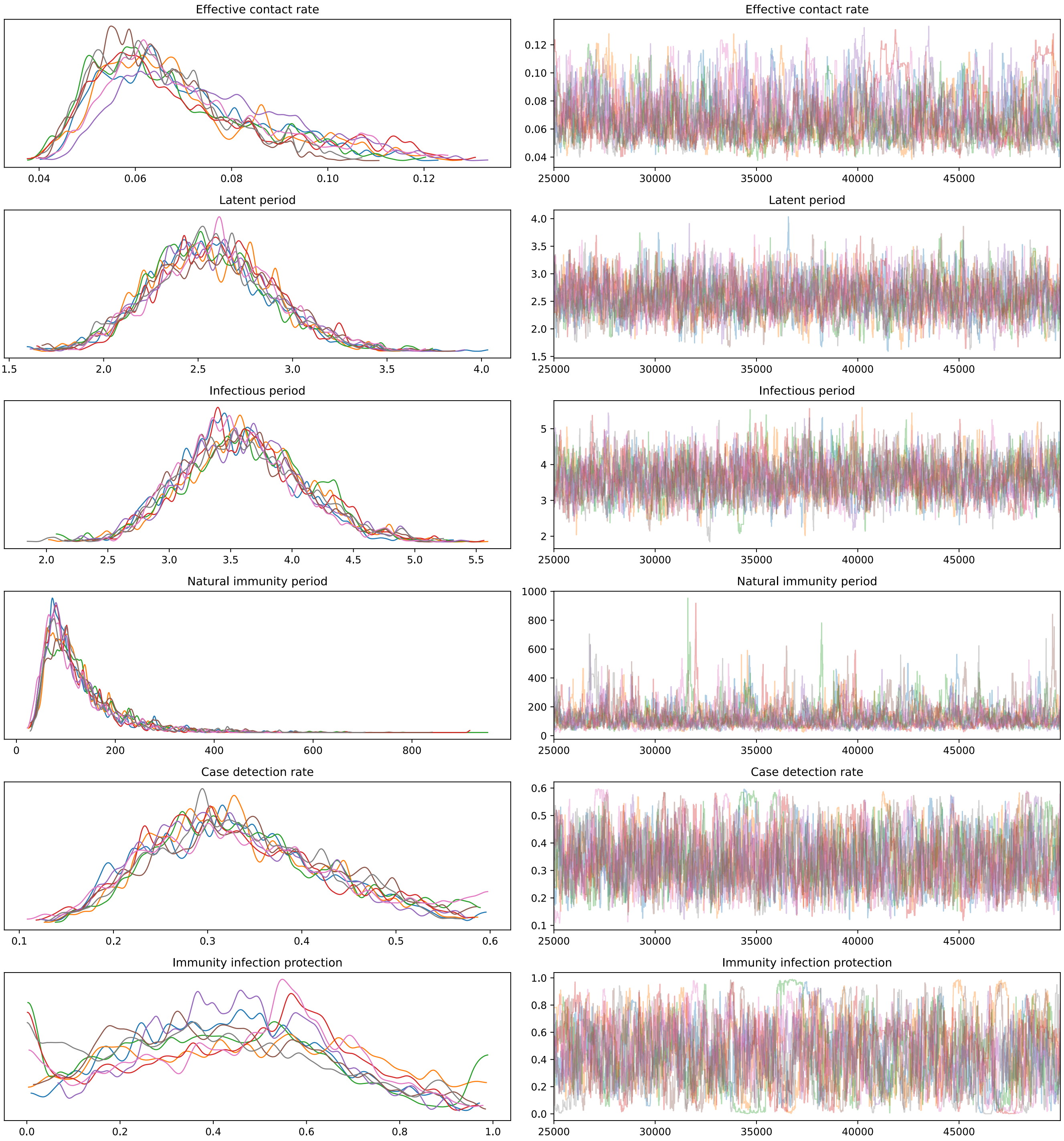

***Figure 28:*** **Parameter posteriors and traces by chain, 2.**

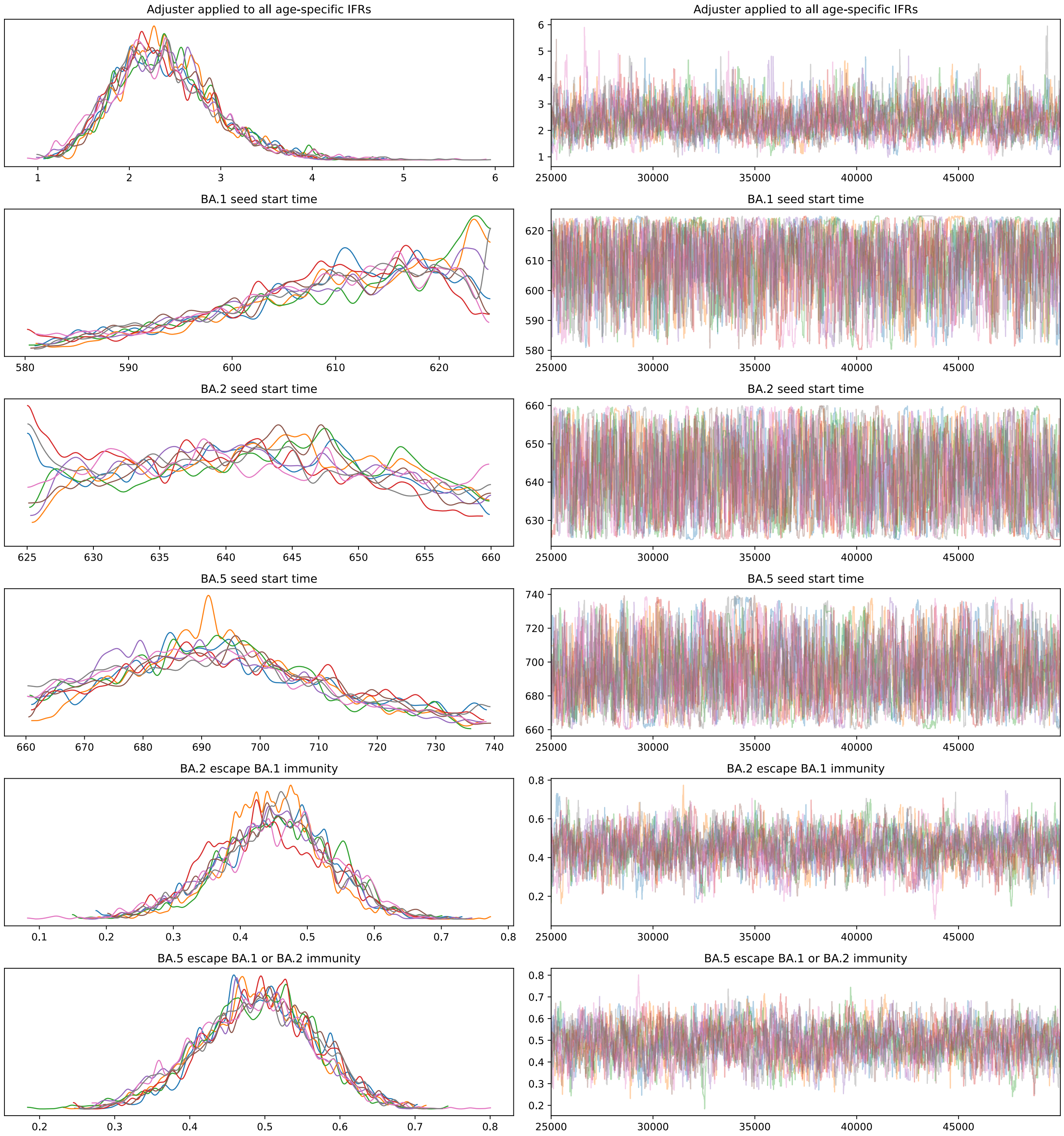

***Figure 29:*** **Parameter posteriors and traces by chain, 3.**

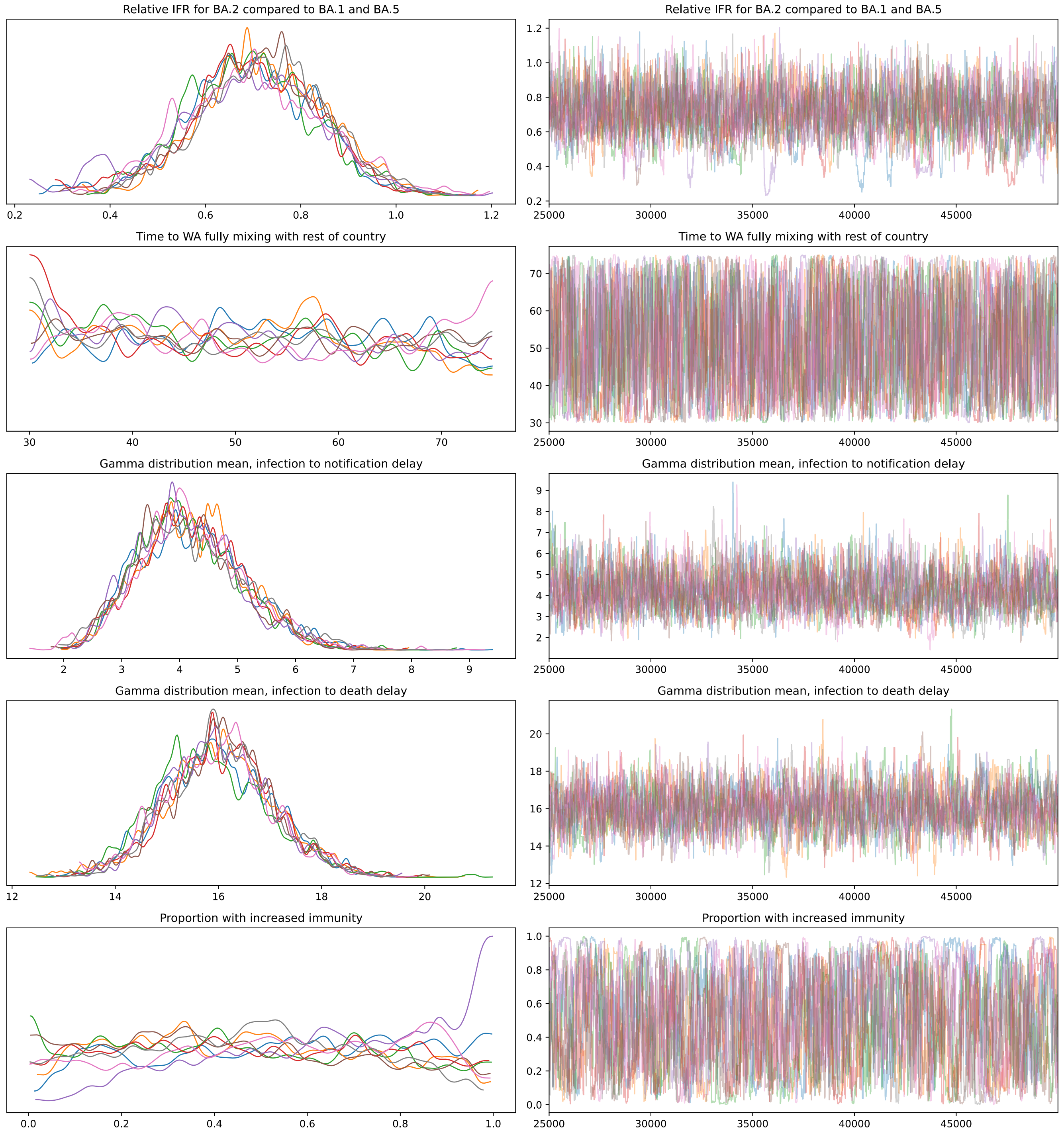

***Figure 30:*** **Posterior densities and prior distributions under the 'none' analysis, 1.** Inferred parameter posterior densities (blue areas) compared against corresponding calibration algorithm prior distributions (grey areas).

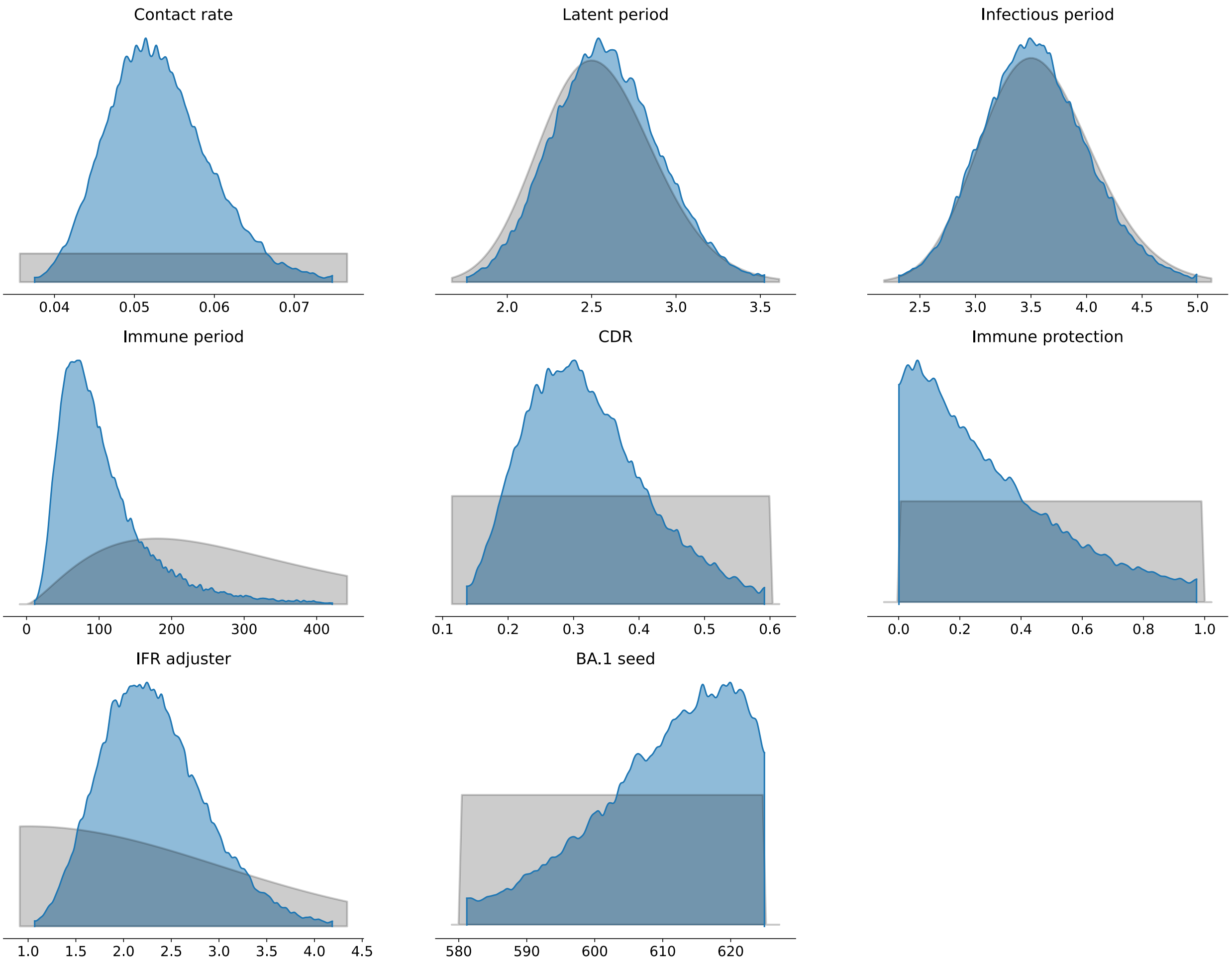

*Figure 31:* **Posterior densities and prior distributions under the 'none' analysis, 2.** Inferred parameter posterior densities (blue areas) compared against corresponding calibration algorithm prior distributions (grey areas).

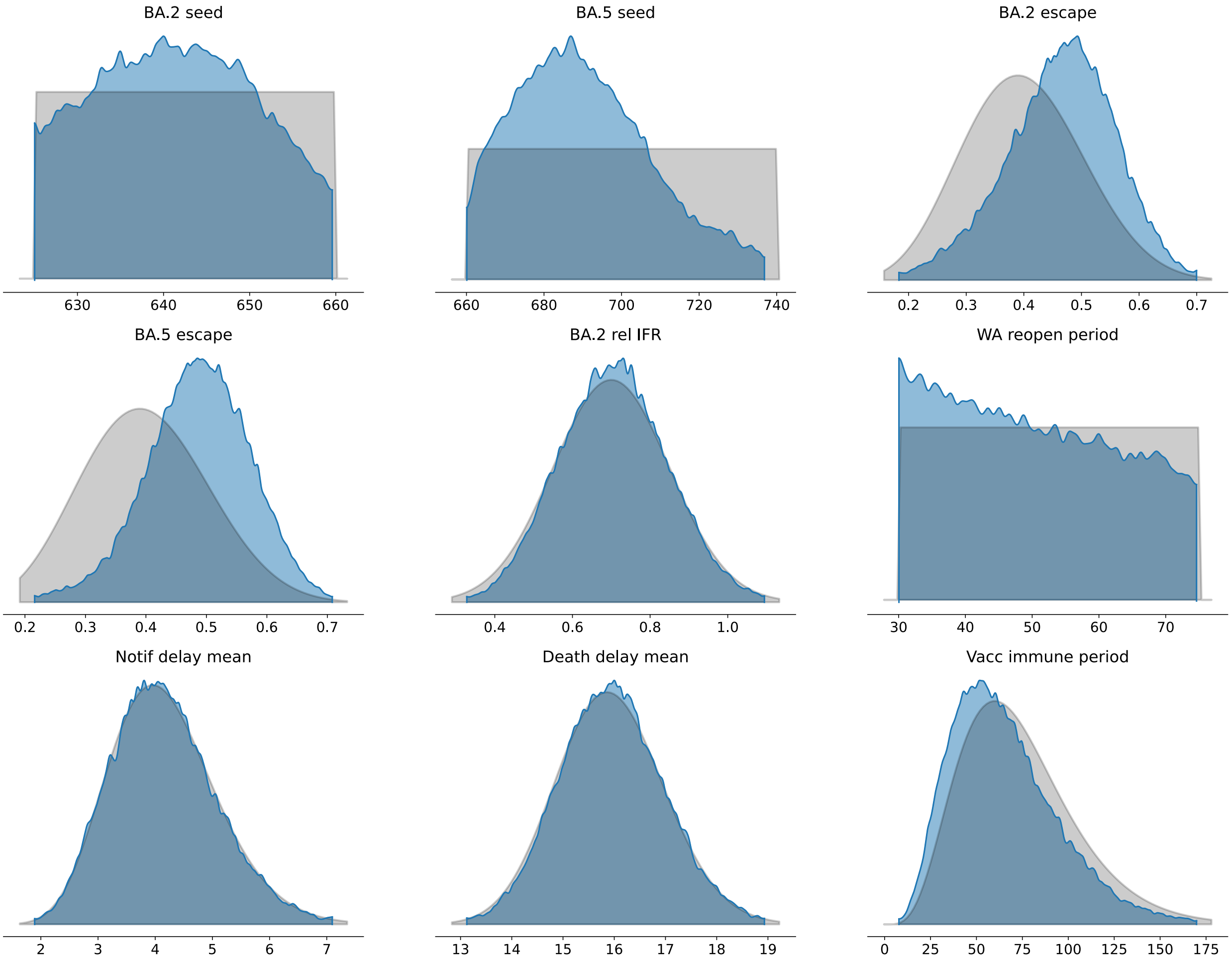

***Figure 32:*** **Posterior densities and prior distributions under the 'mob' analysis, 1.** Inferred parameter posterior densities (blue areas) compared against corresponding calibration algorithm prior distributions (grey areas).

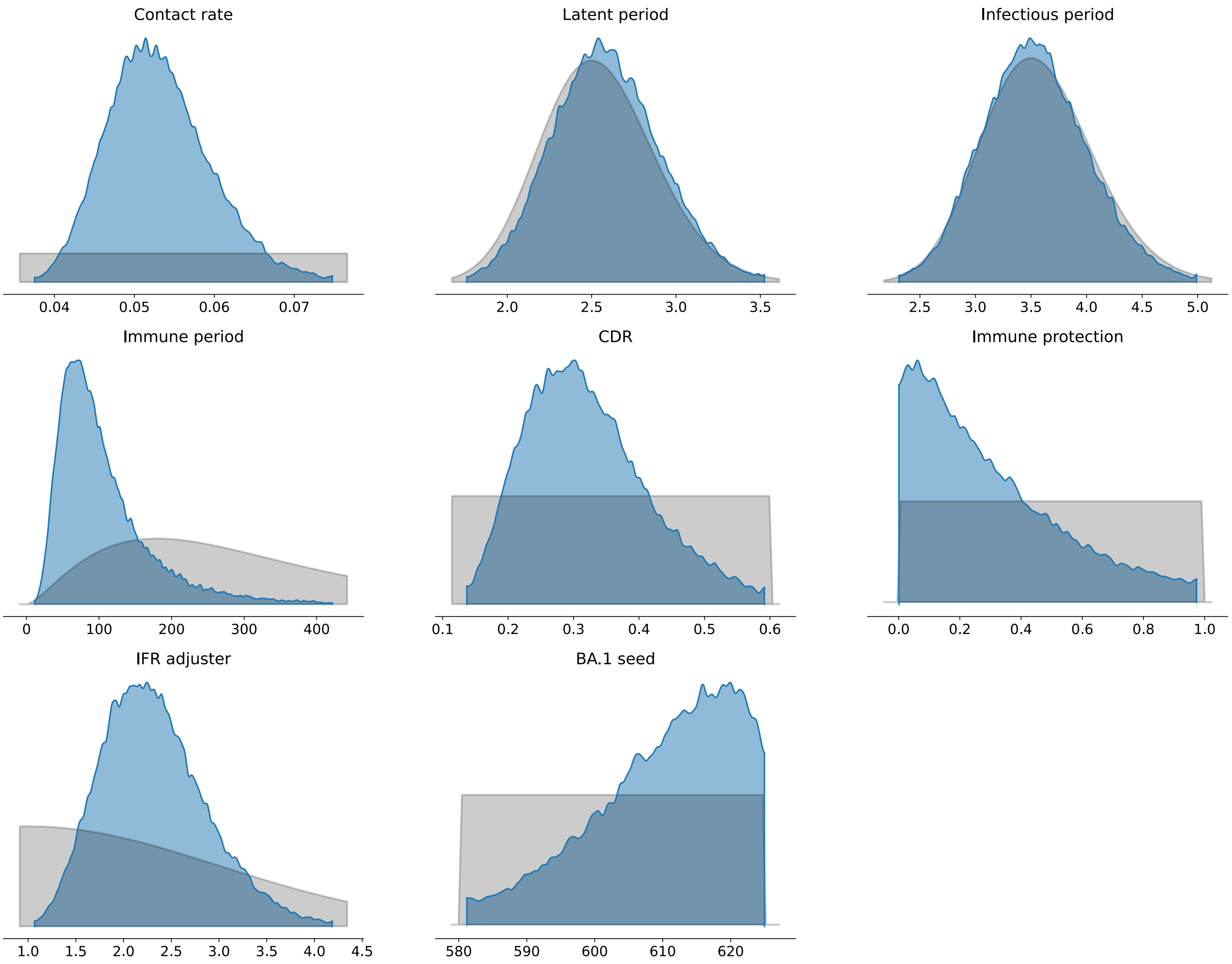

***Figure 33:* Posterior densities and prior distributions under the 'mob' analysis, 2.** Inferred parameter posterior densities (blue areas) compared against corresponding calibration algorithm prior distributions (grey areas).

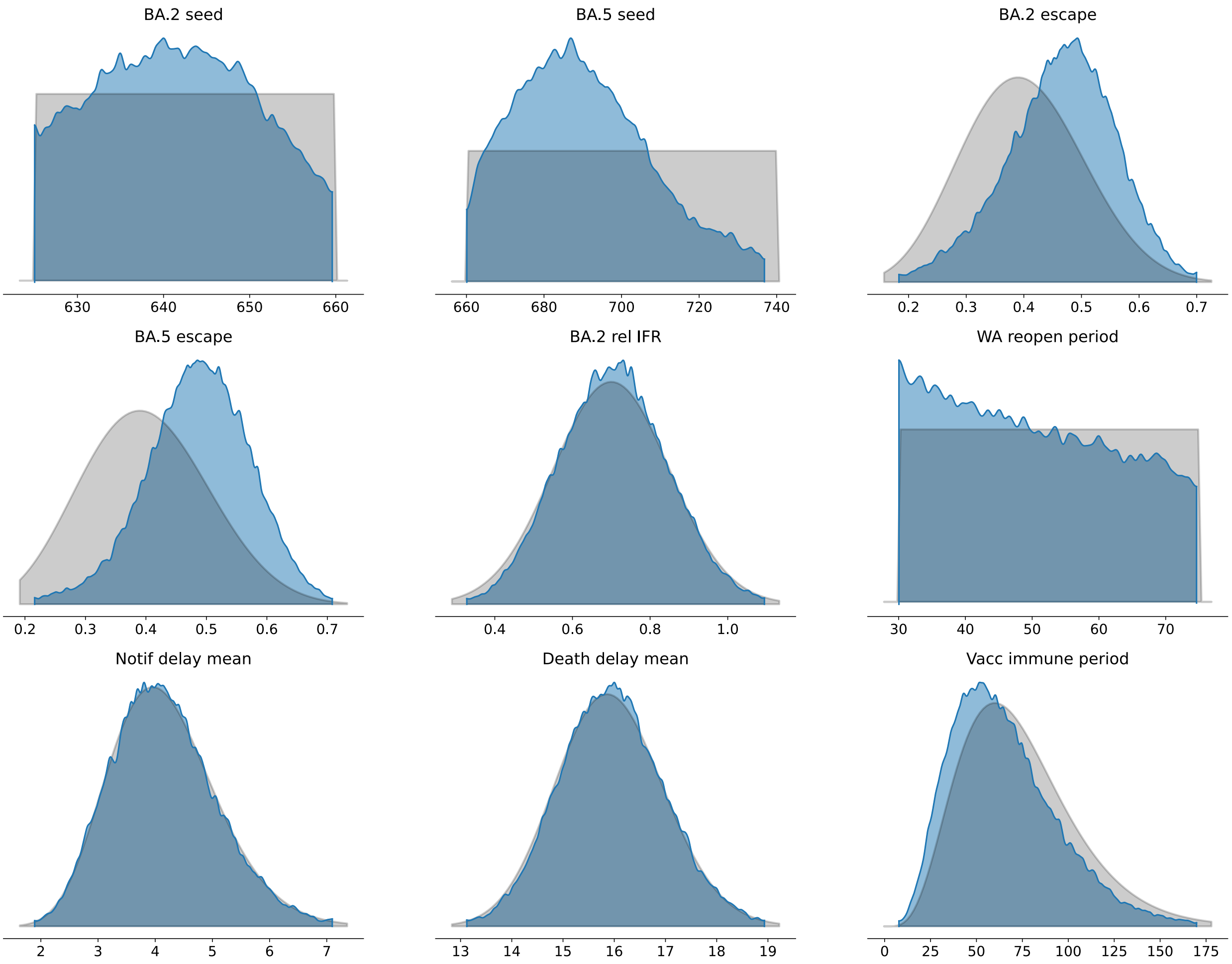

*Figure 34:* **Posterior densities and prior distributions under the 'vacc' analysis, 1.** Inferred parameter posterior densities (blue areas) compared against corresponding calibration algorithm prior distributions (grey areas).

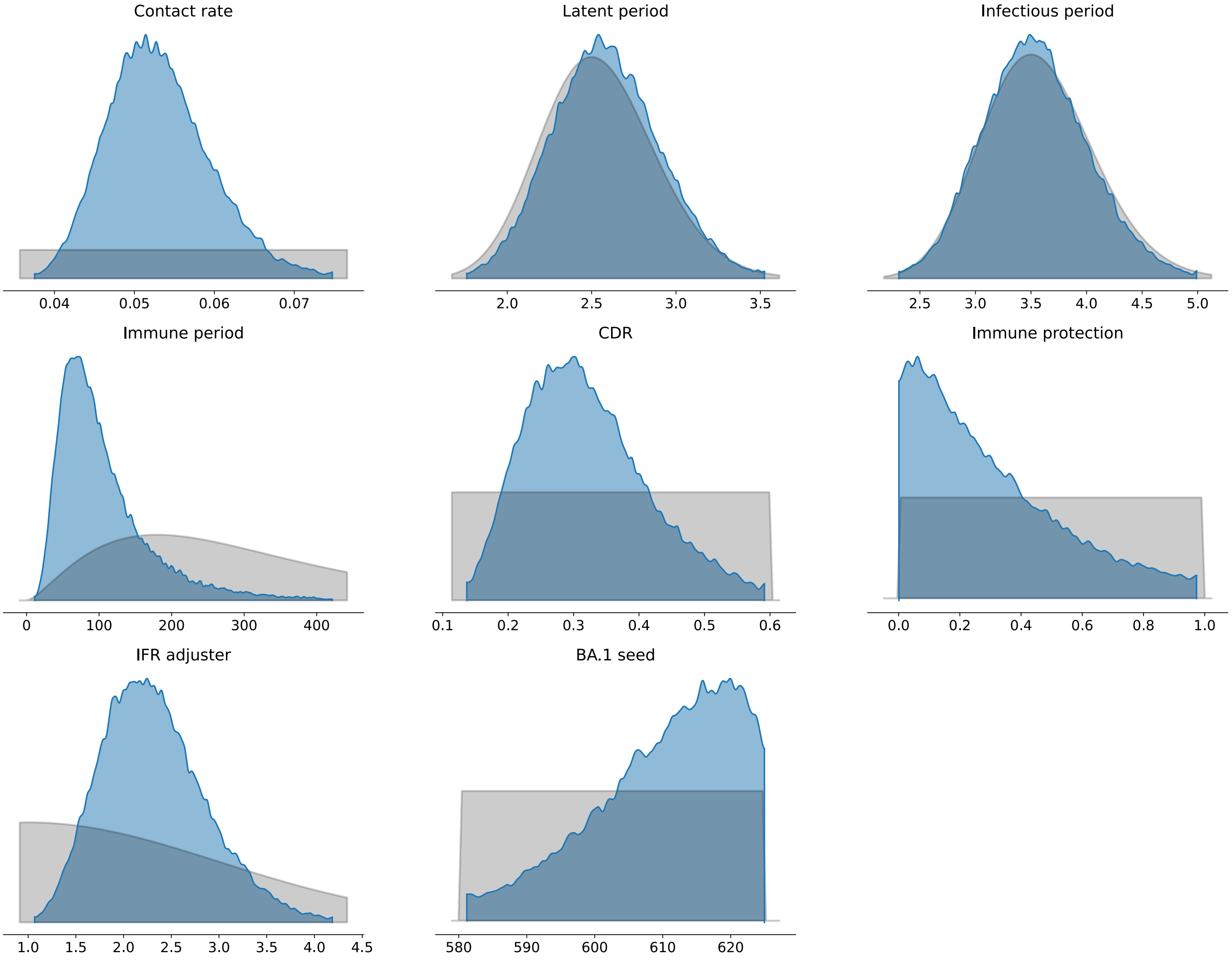

***Figure 35:*** **Posterior densities and prior distributions under the 'vacc' analysis, 2.** Inferred parameter posterior densities (blue areas) compared against corresponding calibration algorithm prior distributions (grey areas).

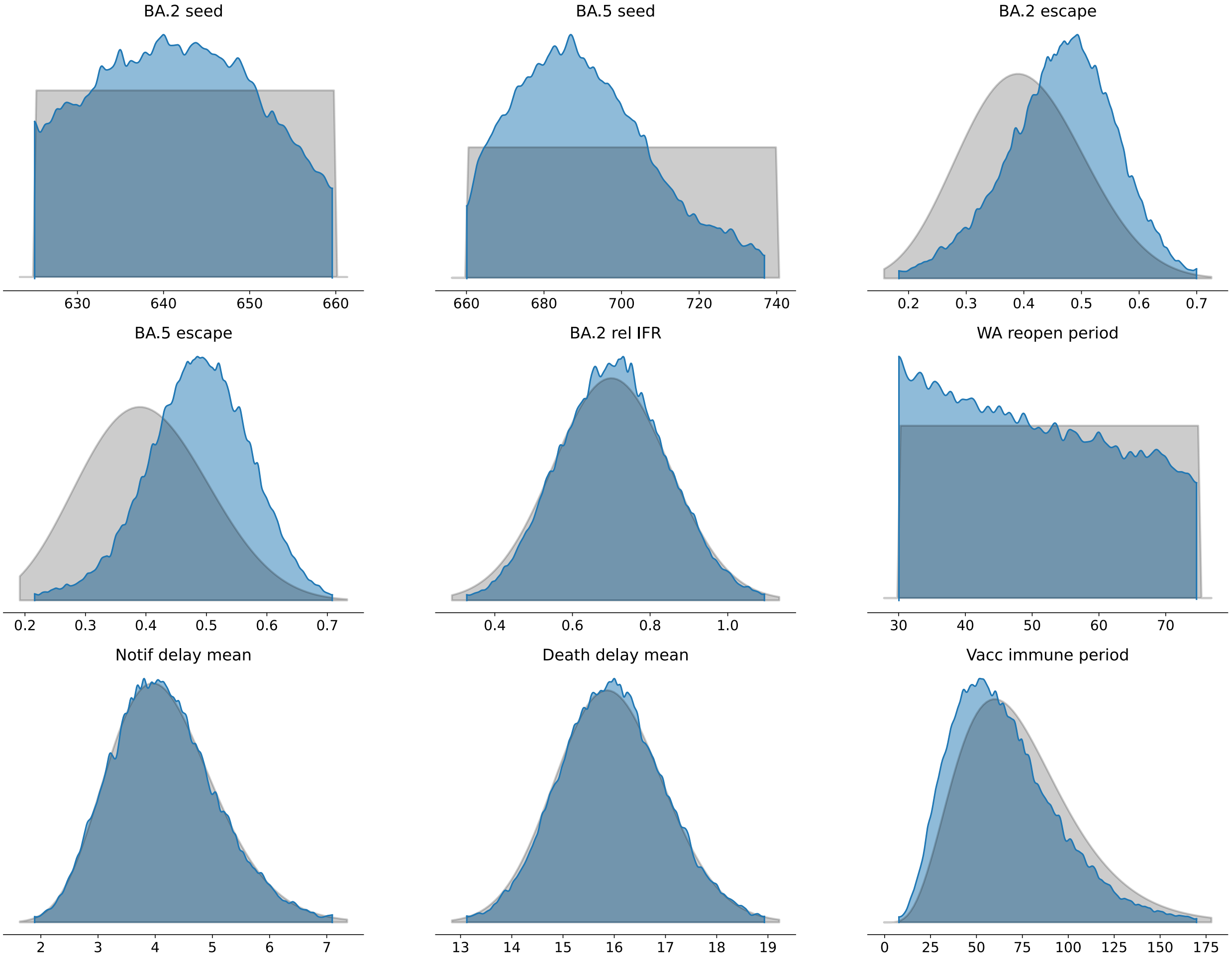

***Figure 36:* Posterior densities and prior distributions under the 'both' analysis, 1.** Inferred parameter posterior densities (blue areas) compared against corresponding calibration algorithm prior distributions (grey areas).

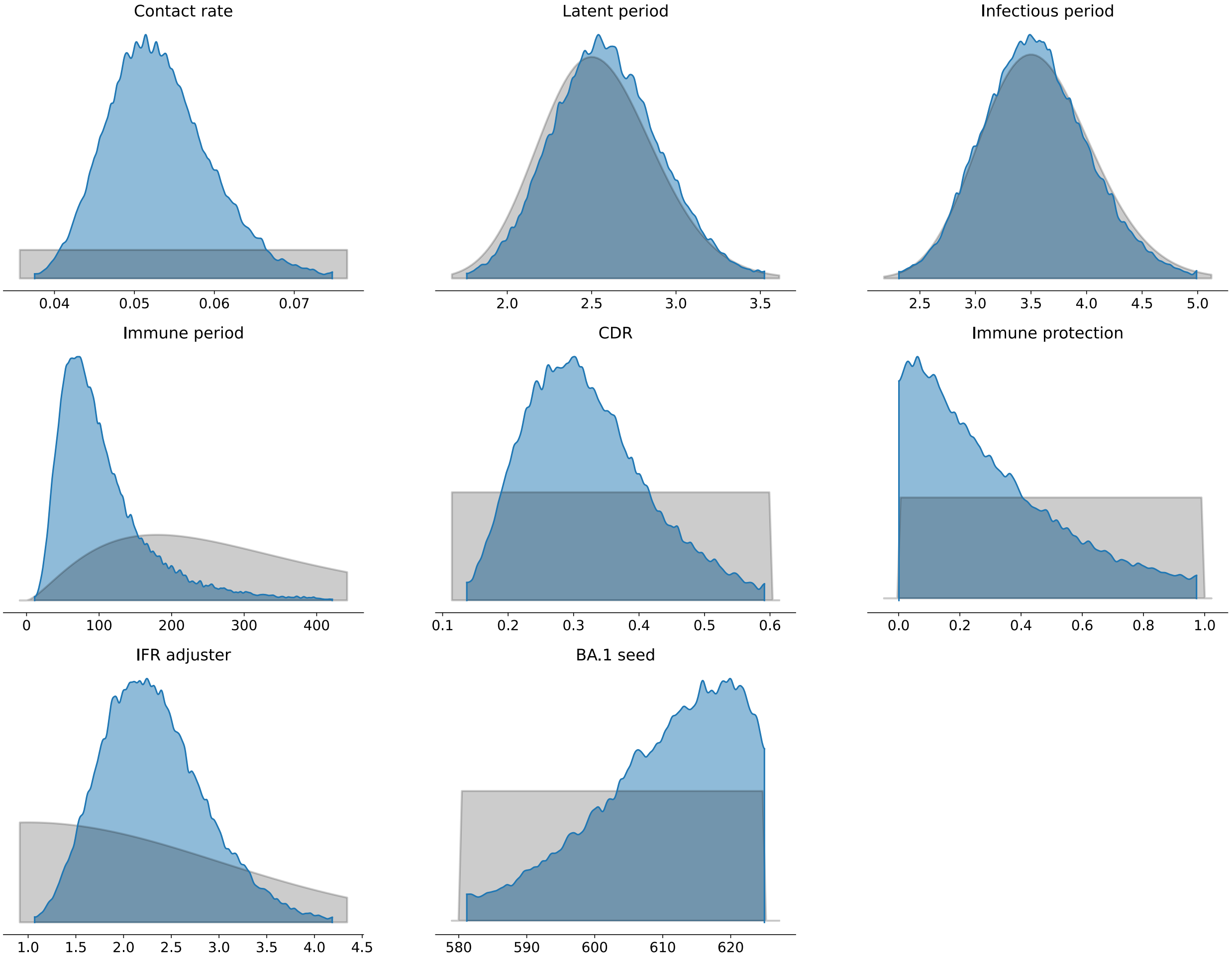

***Figure 37:* Posterior densities and prior distributions under the 'both' analysis, 2.** Inferred parameter posterior densities (blue areas) compared against corresponding calibration algorithm prior distributions (grey areas).

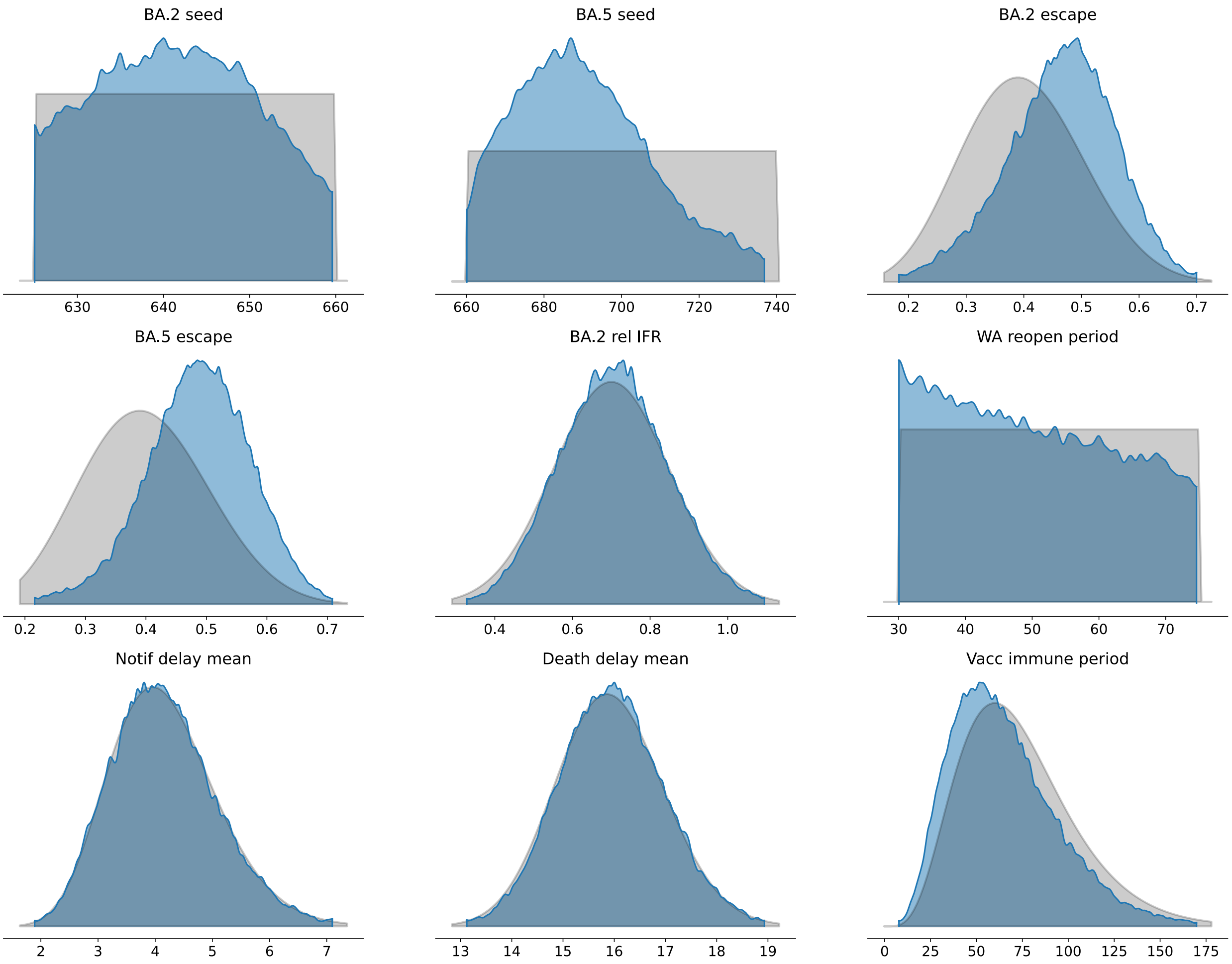

***Figure 38:* All parameter correlation plot matrix.**

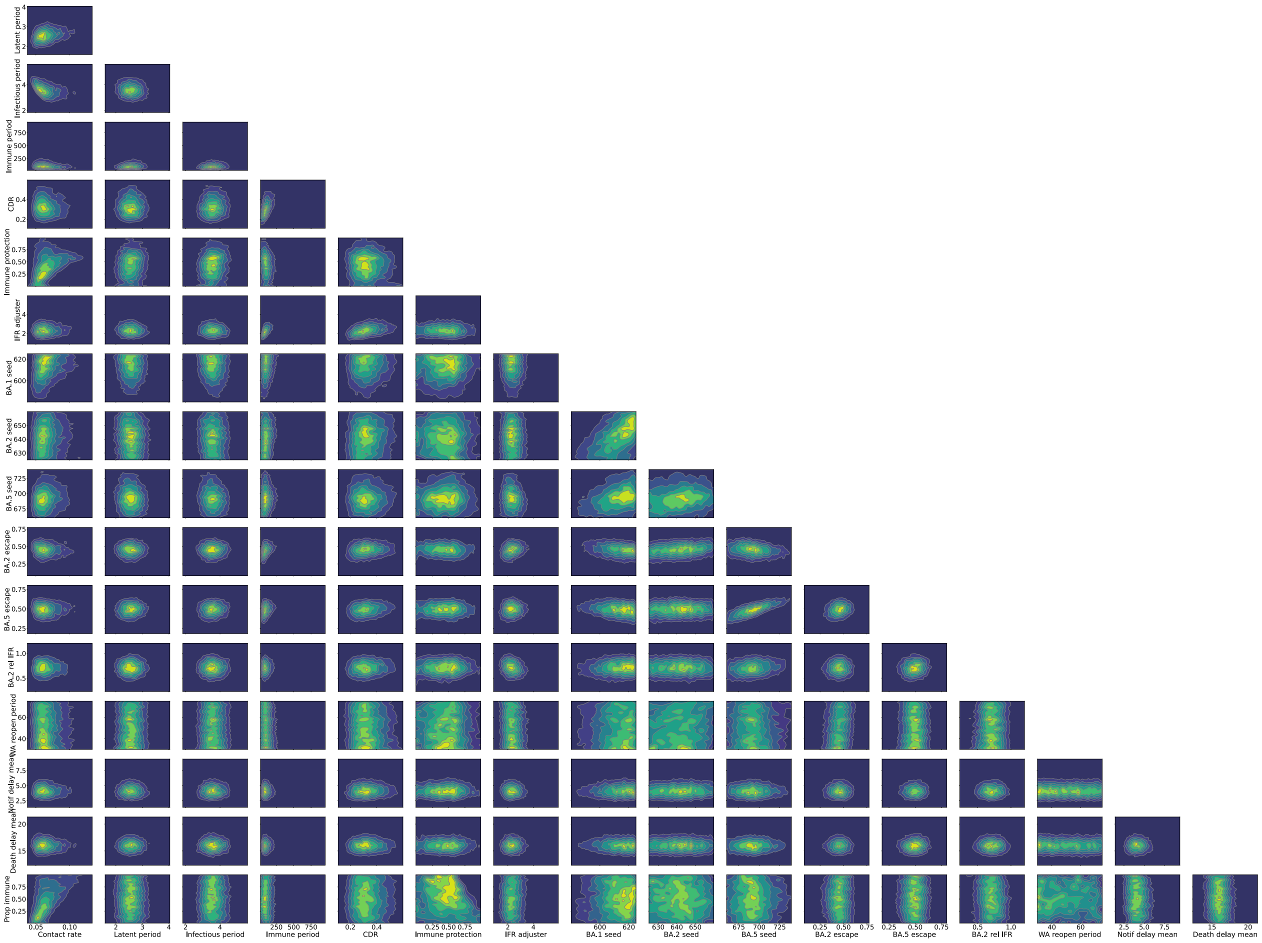

***Figure 39:*** **Selected parameter correlation plot matrix.**

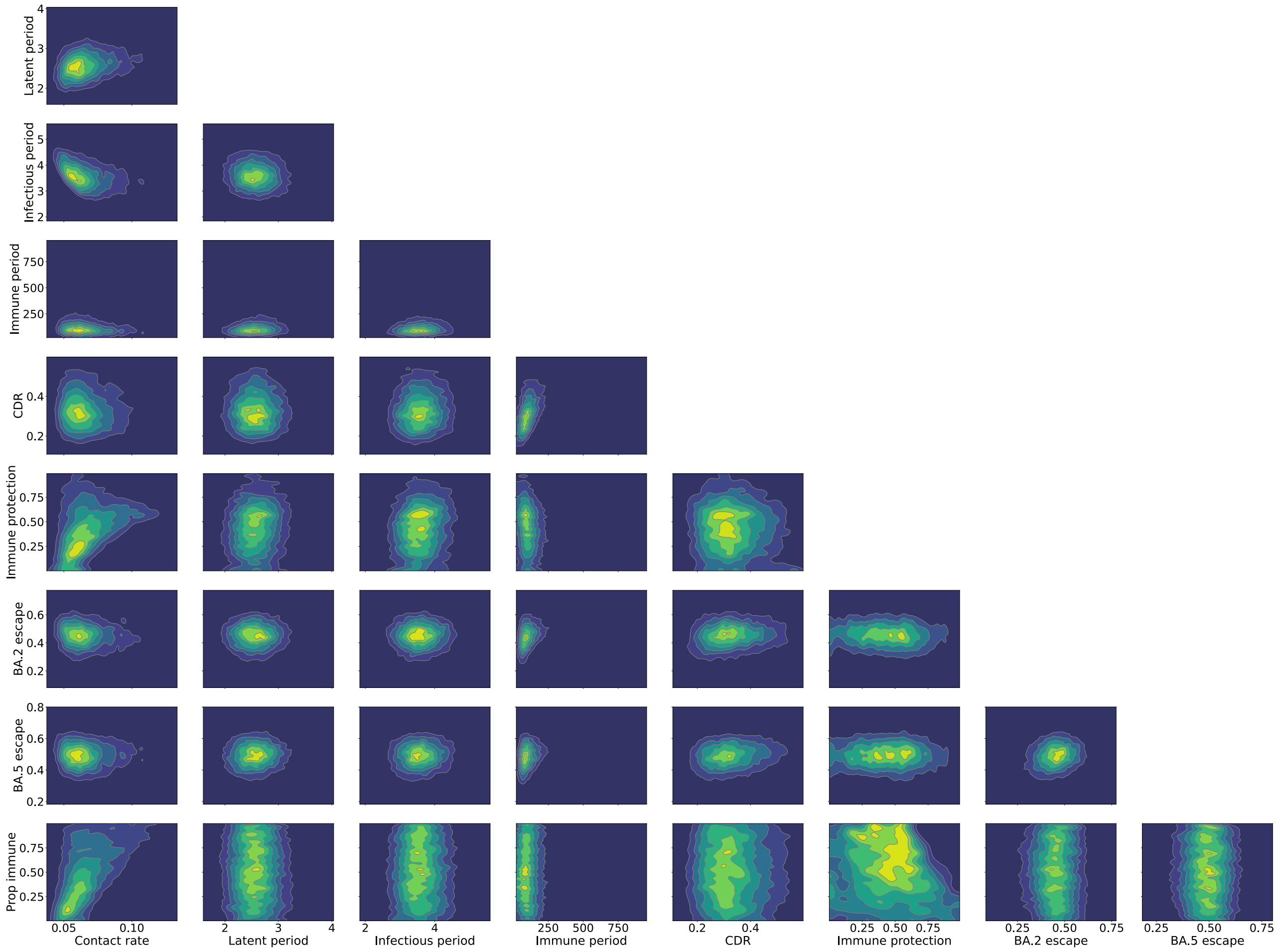

***Figure 40:*** **Immunity-related parameter correlation plot matrix.**

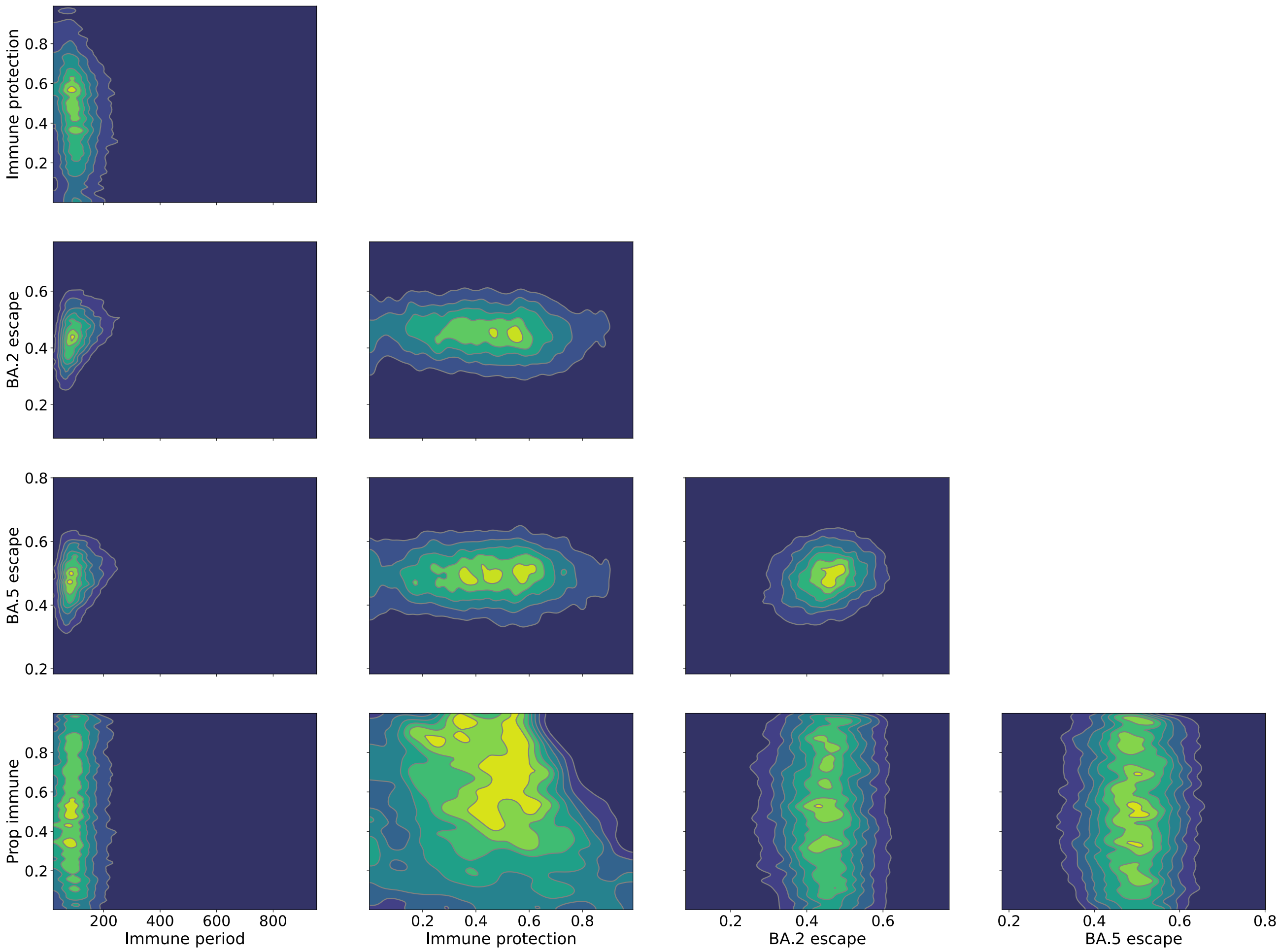
